# Genetics, primary care records and lifestyle factors for short-term dynamic risk prediction of colorectal cancer: prospective study of asymptomatic and symptomatic UK Biobank participants

**DOI:** 10.1101/2023.12.21.23300244

**Authors:** Samantha Ip, Hannah Harrison, Juliet A. Usher-Smith, Matthew Barclay, Jonathan Tyrer, Joe Dennis, Xin Yang, Michael Lush, Cristina Renzi, Nora Pashayan, Spiros Denaxas, Georgios Lyratzopoulos, Antonis C. Antoniou, Angela Wood

## Abstract

**Objectives:** To quantify the contributions of polygenic scores, primary care records (presenting symptoms, medical history and common blood tests) and lifestyle factors, for short-term risk prediction of colorectal cancer (CRC) in both all and symptomatic individuals.

**Design:** Prospective cohort study.

**Setting:** UK Biobank with follow-up until 2018.

**Participants:** All participants with linked primary care records (n=160,507), and a subcohort of participants with a recent (last two years) presentation of a symptom associated with CRC (n=42,782).

**Main outcome measures:** Outcome was the first recorded CRC diagnosis within two years. Dynamic risk models with time-varying predictors were derived in a super-landmark framework. Contributions to model discrimination were quantified using novel inclusion-order-agnostic Shapley values of Harrel’s C-index using cross-validation.

**Results:** C-indices [95% CIs] were 0.73 [0.72-0.73] and 0.69 [0.68-0.70] for the models derived in all and symptomatic participants respectively. The Shapley contributions to model discrimination [95% CIs] differed between the two groups of participants for different predictors: 33% [25%-42%] (34% [9%-75%] in the symptomatic participants) for core predictors (e.g., age, sex, smoking), 16% [8%-26%] (8% [-21%-35%]) for polygenic scores, 32% [19%-43%] (41% [16%-73%]) for primary care blood tests, 11% [4%-17%] (9% [-25%-37%]) for primary care medical history, 6% [0%-11%] (–5% [-32%-13.4%]) for additional lifestyle factors and 3% [-2%-7%] (13% [-19%-41%]) for symptoms.

**Conclusions:** Polygenic scores contribute substantially to short-term risk prediction for CRC in both general and symptomatic populations; however, the contribution of information in primary care records (including presenting symptoms, medical history and common blood tests) is greater. There is, however, only a small contribution by the additional lifestyle risk factors which are not routinely collected in primary care.

## INTRODUCTION

Colorectal cancer (CRC) is the third most common cancer worldwide, with incidence expected to rise (1). Although early stage diagnosis is strongly correlated with improved survival – five-year survival rates for patients in England diagnosed with stage I and IV CRC are 91% and 11% respectively (2) – more than half of incident CRC cases are diagnosed at late stage (57%). Around 90% of CRC cases present with symptoms in primary care prior to diagnosis, however, around one in five of these patients go on to be diagnosed via emergency presentation (2,3). The widespread use of CRC screening has increased both early stage diagnosis and survival rates, however, the programme is constrained by cost-effectiveness considerations and endoscopic capacity (4,5). Models that can predict the likelihood of a CRC diagnosis could support referral decisions for symptomatic patients in primary care (6) or identify high-risk people in the general population to prioritise for screening (e.g. earlier or more frequently) (7).

A range of individual level characteristics (including demographics and lifestyle factors) are associated with CRC risk. Additionally, the use of genetic predictors, such as polygenic scores (PGS) which characterise cancer predisposition, hold substantial potential (8). Healthcare systems have already adopted cancer risk assessments that incorporate genetic predictors (9) for example, the BOADICEA breast cancer model is currently under evaluation for potential implementation in a range of healthcare settings (10,11). Further, electronic health records, which encompass symptoms, blood test results and medical history, are well suited for developing risk assessment tools that can be implemented into healthcare systems. The longitudinal structure of these records supports the dynamic evaluation of risk, enabling periodic updates to predictors and risk as new information becomes available for each individual.

Previous models for predicting the risk of CRC diagnosis have incorporated genetic alongside phenotypic risk factors (12) or utilised electronic health records (13,14). However, no existing CRC model has fully harnessed the longitudinal information contained in electronic health records (7,15) in conjunction with genetics for short-term CRC diagnosis (16). Moreover, when evaluating the role of genetic risk in model performance for early cancer detection, previous studies have typically focused on the incremental improvement achieved by adding a PGS to a pre-existing model (12,17). Comparing this to the incremental improvements in performance made by adding other sets of predictors, and to the overall inclusion order independent contribution of each group should provide further insights into the relative importance of different types of risk factors.

In this study, we derive and internally validate a dynamic prediction model for the diagnosis of CRC using individual-level data from participants in UK Biobank (UKB), incorporating demographics, PGS, primary care data (presenting symptoms, medical history, and common blood tests) and additional lifestyle factors. We aimed to quantify the inclusion-order-agnostic contribution of these six predictor sets to model performance in both a general and symptomatic population.

## METHODS

### Study population

We used data from UKB, a prospective population-based cohort (n=502,371) of UK residents aged 40-69 at enrolment (2006–2010) (18). All participants attended a baseline assessment that collected detailed information about demographics, medical history and lifestyle. Blood samples were taken and genotype data are available for 488,377 participants. Primary care records, including coded information from GP consultations and prescriptions, are available for approximately half of the cohort (n=228,913) with records available up to 2018.

Analyses were restricted to UKB participants with available genetic data and linked primary care data. First degree relatives were excluded by random selection of one of each pair of relatives (so that we could assume an unrelated population when considering genetic risk). Participants with a diagnosis of any cancer, except non-melanoma skin cancer, before baseline assessment were also excluded, ensuring that identified cases of CRC in follow-up were incident primary cancers (Figure S1).

### Super-landmark framework

We structured the cohort into landmark age datasets at landmark ages 40, 41, 42, … up to 74 years. Participants were included in a landmark age dataset if they were alive, had at least six months of continuous primary care records (no gaps >90 days; where multiple continuous periods are available we used the most recent) in the previous two years and had not previously received a cancer diagnosis (except non-melanoma skin cancer) before the landmark age. Participants could be included in multiple landmark age datasets as they aged through the cohort. The date when a participant entered a landmark age dataset (1st of the month in which they reach a landmark age) is referred to as the “index date”. Time-varying predictors and outcomes (defined below) were extracted for each participant at all their index dates. The landmark age datasets were stacked to make a “super-landmark dataset” subsequently referred to as the “study cohort” (19,20). For further details see Supplementary Methods 1.

### Outcomes

CRC outcomes were extracted from the national cancer registries (provided by: Medical Research Information Service, National Cancer Intelligence Network, NHS England, NHS Central Register, Scottish Cancer Registry and Public Health Scotland) – available up to June 2022 – using ICD codes (see GitHub repository) (21,22). CRC diagnosis events (cases) were defined as individuals with a first cancer diagnosis of CRC in the two years following an index date. The super-landmark approach allowed for CRC diagnoses at any time between baseline assessment (2006–2010) to the study end date (2020) to be included in the analysis. Follow-up was censored at the first diagnosis of any incident cancer (excluding non-melanoma skin cancer), death (via linkage to death registry), two years after the end of primary care data availability or two years after the index date.

### Risk predictors

We selected candidate predictors as follows: (i) well-established CRC risk factors (23–25); (ii) systematic literature reviews of CRC risk models (6,7); (iii) analyses of CRC symptoms (26,27), pre-existing conditions (both modifiers of CRC risk and potential misdiagnoses) (28,29) and blood test results (30); and (iv) consultation with clinical and academic experts.

Some candidate predictors, such as family history and smoking behaviour, were collected during the UKB baseline assessment. There is missing data for a small number of variables drawn from baseline assessment (Supplementary Methods 2); missing data is included as a separate category for both smoking status and ethnicity, however, for all other variables with missing data (e.g. alcohol consumption) we used a complete case approach (Figure S1). We used the multi-ancestry PRS-CSx, a PGS developed in a cohort of individuals with European or East Asian ancestry to characterise genetic risk of CRC (31). Candidate predictors drawn from primary care records (medical history, symptoms and blood tests) were extracted respective to each index date. We included candidate predictors for both the occurrence and results of blood tests that could indicate iron deficiency anaemia or inflammation.

We grouped the predictors into six sets for analysis (Table S1): core (8 predictors), polygenic score, symptoms (16 predictors), medical history (12 predictors), common blood tests (4 predictors) and other lifestyle (5 predictors). The groupings were based on both the type of information and the expected data availability from different sources (e.g. primary care records, survey-based questionnaires). See Supplementary Table 1 and Supplementary Methods 2 for more details.

### Defining a Symptomatic Subcohort

We defined a symptomatic super-landmark cohort (subsequently referred to as the “symptomatic subcohort”), by identifying individuals in the study cohort with presenting symptoms relevant to CRC recorded in their primary care records in the two years prior to the landmark age. The relevant symptoms were selected from the 16 predictors in the symptom predictor set using the Akaike information criterion (AIC) based bidirectional stepwise selection with a Cox proportional hazard (Cox PH) model, resulting in the selection of four symptoms: new onset haemorrhoids, new onset constipation, recent record of rectal bleeding and new onset diverticular disease (see Table S1, Fig S3, Supplementary Methods 2).

### Model Development and Validation

We developed dynamic early detection models for CRC diagnoses within two years of an index date. A Cox PH model was developed using the super-landmark dataset with robust standard errors to account for the inclusion of participants multiple times. We selected statistically important predictors using AIC-based bidirectional stepwise selection (Supplementary Methods 3).

We performed bootstrapping by creating 200 bootstrap samples from the entire dataset. For each sample, the data were split into training and testing datasets by person ID, ensuring no individual contributed to both. Model discrimination was quantified by Harrell’s C-index for all possible combinations of the six predictor sets. To quantify and fairly compare the distinct contributions to model discrimination from each of the six predictor sets, we calculated the Shapley values as the average marginal C-index across all possible model combinations. Calibration was assessed using calibration decile plots at 2 years. Confidence intervals (95% CIs) for the C-index, Shapley values, and calibration decile points were estimated using the 200 bootstrap samples.

We present the C-indices and their 95% CIs for all predictor combinations, allowing for the assessment of incremental added discriminative power of each predictor set. We also present the Shapley values as the percentage contribution of each predictor set to the overall C-index; these are independent of the order in which the sets are added to the model (order-agnostic) (28–30). For further details see Supplementary Methods 4.

Model development and validation is reported in line with the TRIPOD guidelines for reporting of clinical prediction models (Supplementary Methods 5)(32).

### Sensitivity Analyses

We conducted the following sensitivity analyses: (i) repeating the predictor selection using the group lasso algorithm and comparing both methods to a model with all candidate predictors (no selection) (Supplementary Methods 3); (ii) repeating the analyses using a PGS (LDPred PGS (33)) developed in a European-ancestry only cohort (we note this may overestimate model performance and the genetic contribution as UKB cohort members were included in LDPred development); (iii) redefining the symptomatic subcohort as participants with any CRC symptom (listed in Table S1), except fatigue (not considered to be sufficiently specific), in the two years prior to each index date, and (iv) repeating the analysis excluding the 14,743 participants with English Vision primary care data (linked primary care records for participants registered English Vision who died pre-2017 are mostly unavailable) (34).

### Patient and public involvement

The Centre for Cancer Genetic Epidemiology has a panel of 6 patients and members of the public (3 with personal cancer experience and 3 without) to advise on all aspects of the cancer risk modelling research incorporating/relating to genetic predisposition to cancer undertaken by members of the centre, under the CanRisk programme of work. They are involved in reviewing the research protocols, the co-design of research and provide feedback on research findings.

## RESULTS

### Cohort Characteristics

We identified 160,507 UKB participants with genetic data and linked primary care records (Figure S1); their characteristics were similar to the whole UKB cohort (Table S7). Within this study cohort we identified 42,782 symptomatic participants (the symptomatic subcohort). The characteristics of participants in the study cohort and symptomatic subcohort had some differences (Table 1); individuals in the symptomatic subcohort were older (59.5 (IQR: 52.0-64.3) compared to 57.9 (IQR: 50.3-63.4) in the study cohort), less likely to be male (43.8% and 47.3%) and more likely to have ever smoked (47.2% and 44.4%). The most commonly recorded CRC symptoms in the study cohort were new-onset constipation (16.8%) and new-onset diarrhoea (15.3%). Most participants (81.0%) had at least one relevant medical history indicator, including: colonoscopy in the last ten years (23.6%) and regular use of non steroidal anti-inflammatory drugs (NSAIDs) (14.8%). Most participants (87%) also had at least one recent blood test that could be used to determine the presence of inflammation (such as C-Reactive Protein) or iron-deficiency anaemia (such as ferritin), with 29% having at least one abnormal result. More symptoms, indicators of medical history and common blood tests were observed for older participants (Figure S2). We identified 1356 CRC diagnosis events (cases) in the study cohort and 237 in the symptomatic subcohort (Table S5).

**Table 1:**
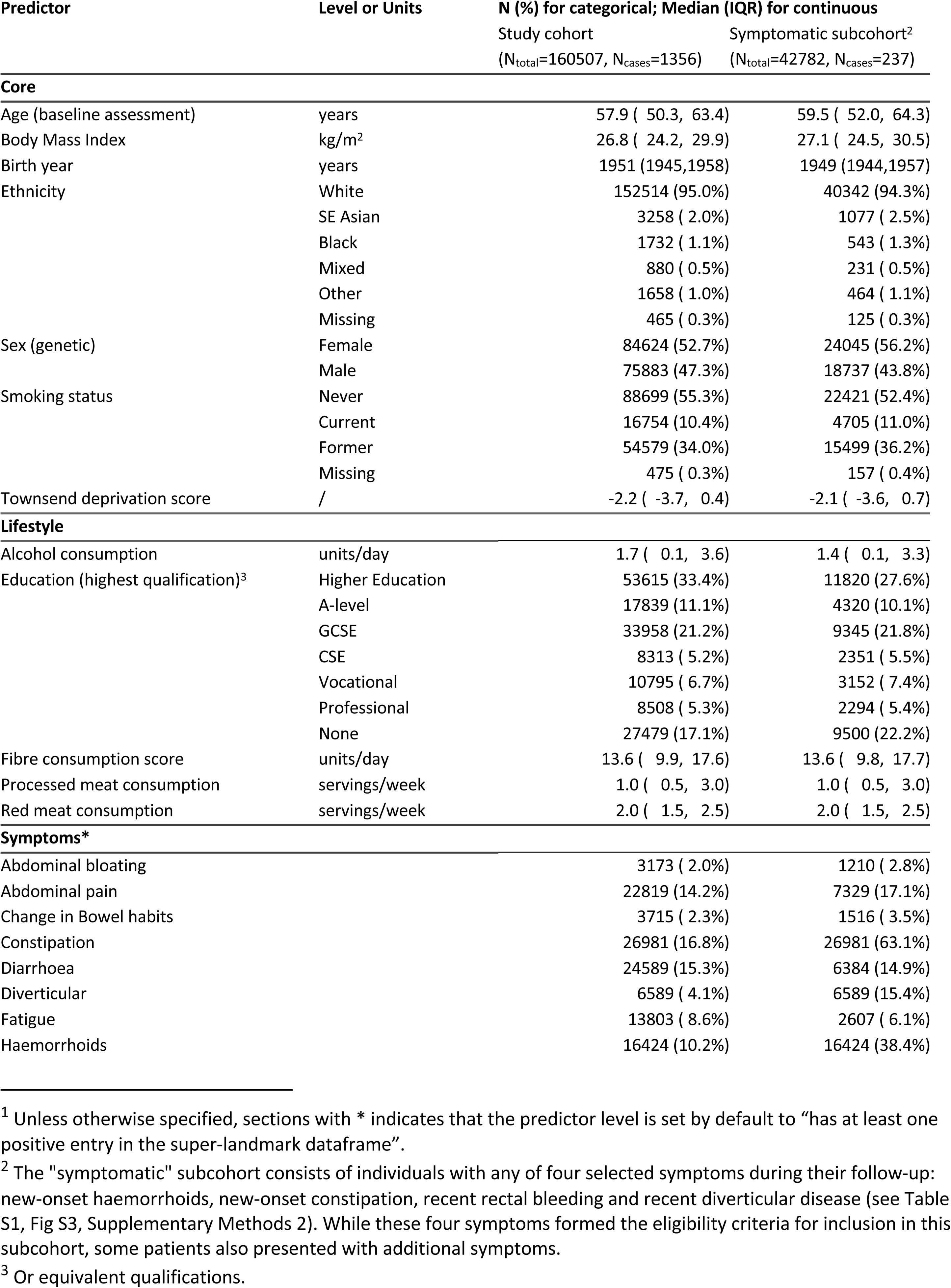
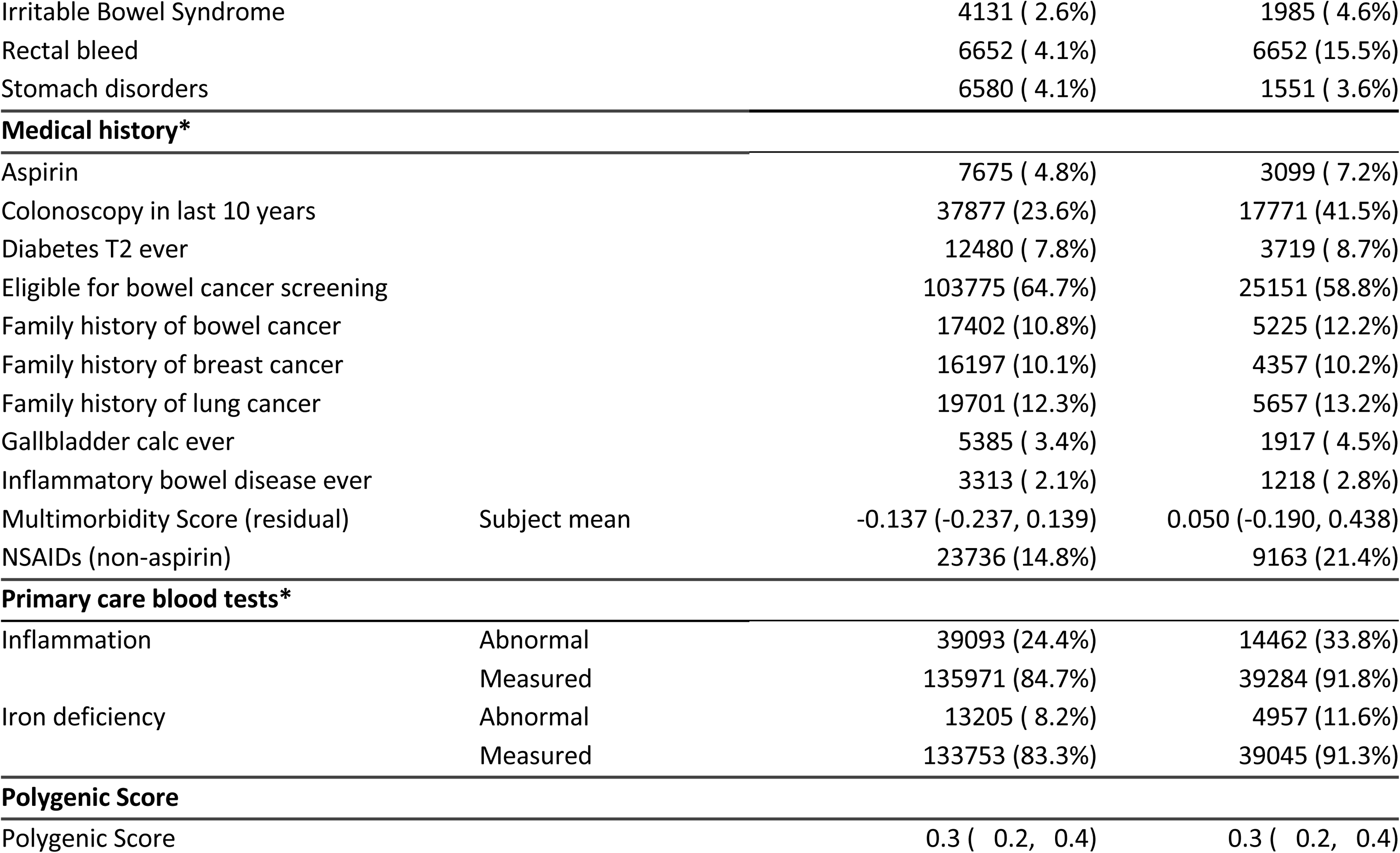
Participant characteristics: Study and symptomatic cohorts^1^.

### Model Development

In the study cohort, 23 (of 39) candidate predictors were selected as important risk predictors of CRC (Figure 1a, Table S2a). This included predictors from each of the six predictor sets. The strongest predictors of CRC (hazard ratio (HR) [95% CI]) other than age were: iron-deficiency anaemia (3.9 [3.4-4.6]) and rectal bleeding (2.7 [2.0-3.7]). The PGS also had a strong association with CRC (1.4 [1.3-1.5] per SD increase).

**Figure 1a.**
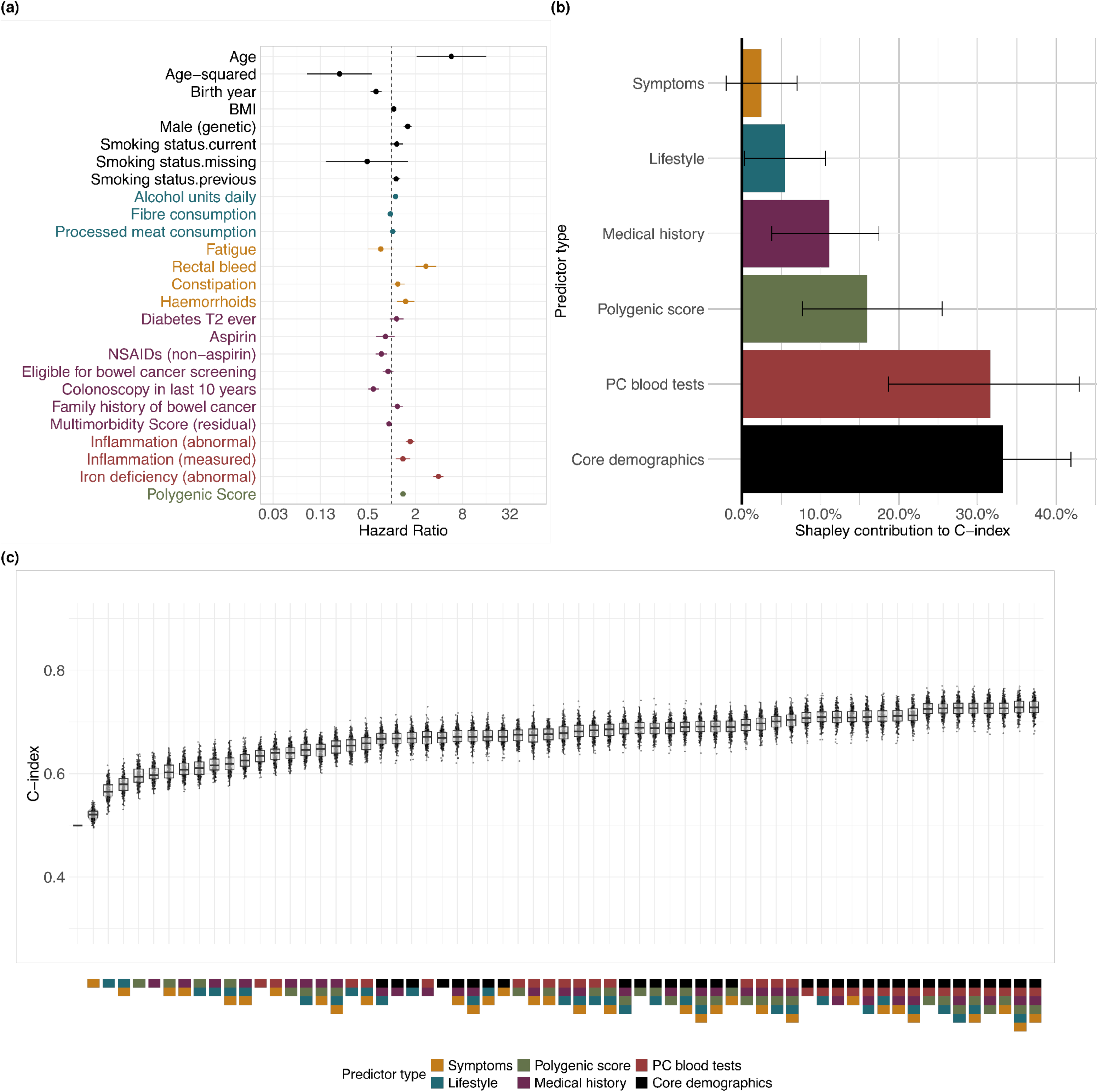
Hazard ratios from bidirectional stepwise Cox regression for the **study cohort**; Figure 1b Discriminative contribution of predictors using Shapley values (C-index > 0.5); Figure 1c C-indices from 200 bootstrap samples for each combination of predictor sets. Colour-coding indicates the predictor set in all figures. Overall model C-index: 0.728 [0.726-0.731].

In the symptomatic subcohort, 18 (of 39) candidate predictors were selected (Figure 2a, Table S2b), 15 of which had also been selected in the study cohort. The 3 distinct predictors were abdominal bloating, diverticular disease and education level. Predictors exhibiting the strongest associations with CRC were: iron-deficiency anaemia (4.0 [2.9-5.5]), a record of a recent blood test for inflammation (2.9 [1.2-6.8]), male sex (2.0, [1.5-2.7]) and rectal bleeding (2.0, [1.4-2.9]). Again, the PGS had a strong association with CRC (1.3 [1.2-1.5] per SD increase).

**Figure 2a.**
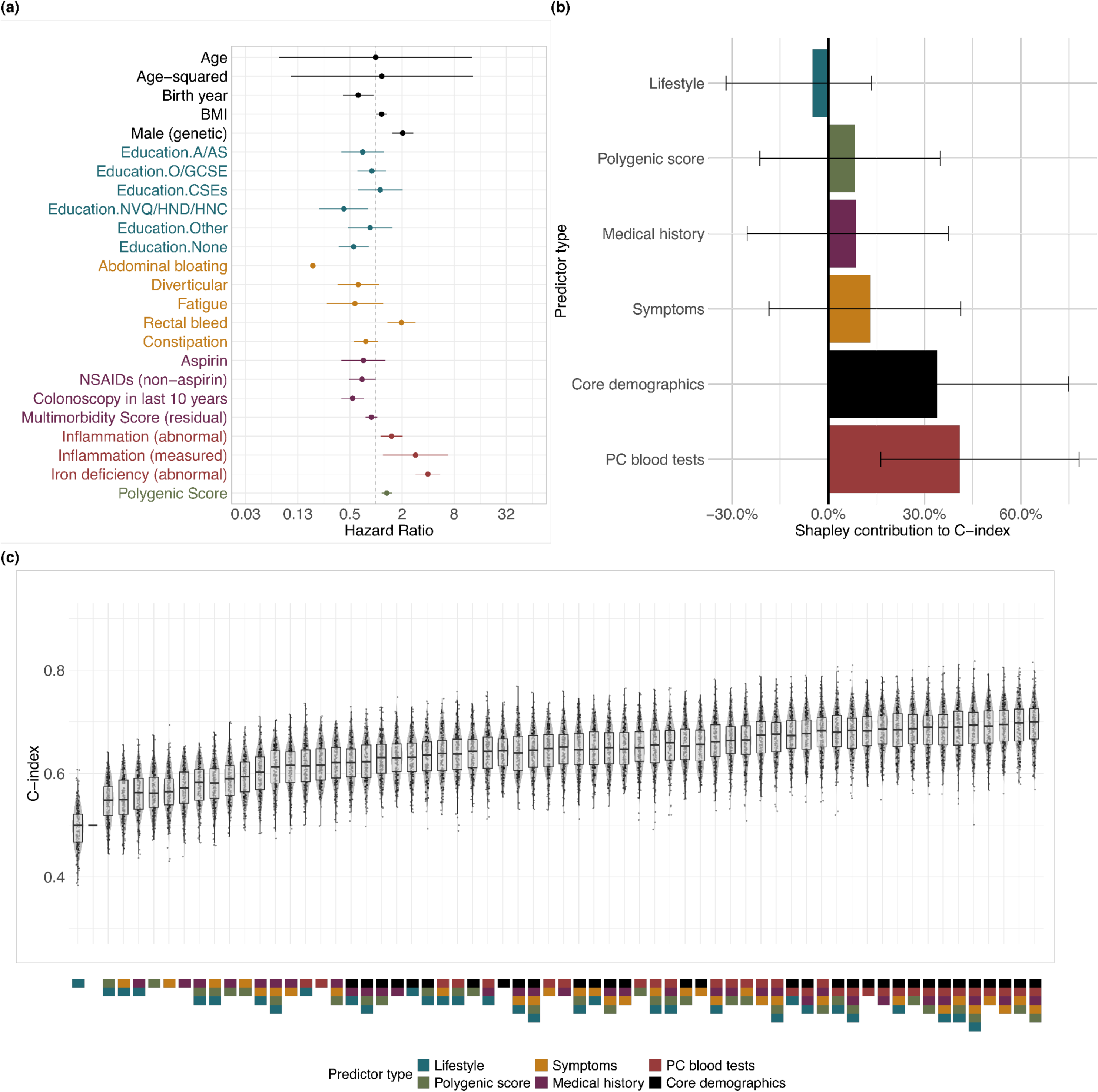
Hazard ratios from bidirectional stepwise Cox regression for the **”symptomatic” cohort**, characterised by haemorrhoids, constipation, rectal bleeding and diverticular disease, as shown in Supplementary Figure 3; Figure 2b Discriminative contribution of predictors using Shapley values (C-index > 0.5); Figure 2c C-indices from 200 bootstrap samples for each combination of predictor sets. Overall model C-index: 0.689 [0.682-0.695].

### Model Performance

The overall discriminative ability (C-index [95% CIs]) of the CRC early detection models developed in the study cohort and symptomatic subcohort was 0.73 [0.72-0.74] and 0.69 [0.68-0.70] respectively. Calibration decile plots at 2 years indicated good calibration for both the study cohort and symptomatic subcohort (Figure S9a-b).

Predictors in the core set (demographics and smoking) made the greatest contribution (33% [25%-42%]) to the discriminative ability of the models in the study cohort (Figure 1, Table S3a), followed by common blood tests (32% [19%-43%]), PGS (16% [8%-26%]) and medical history (11% [4%-17%]). These predictor sets by themselves had discriminatory ability of 0.67 [0.66-0.68], 0.63

[0.63-0.64], 0.60 [0.59-0.60] and 0.60 [0.59-0.61] respectively. Selected predictors in the symptom group made the smallest contribution to overall predictive ability (2.5% [-2.0-7.0%]). The C-indices for each combination of predictor groups were also calculated (Table S3a). The version of the model with only the four highest contributing predictor sets (core, polygenic score, medical history, common blood tests), but without additional lifestyle factors and symptoms, had very similar discrimination to the full model (0.73 [0.72-0.73]).

In the symptomatic subcohort the largest contributions to discriminative ability were made by the core predictors (34% [9%-75%]), primary care blood tests (41% [16%-78%]) and symptoms (13% [– 19%-41%] (Figure 2, Table S3b). The contribution of symptoms was more than five times higher than in the study cohort model and was higher than that made by the PGS (8% [-21%-35%]). Additional lifestyle predictors do not improve the model performance (–5% [-32%-13.4%]; removing this predictor set does not lower overall model discrimination (0.73 [0.72-0.73]).

### Sensitivity Analyses

Lasso selected more predictors than bidirectional stepwise (Figures S5a-b), although the discriminative performance of the models using both selection methods were similar, and comparable to the model with all predictors (no selection) in both the study cohort and symptomatic subcohort (Figures S4a-b).

As expected, replacing the PRS-CSx with LPred PGS resulted in models (in both cohorts) with higher hazard ratios for the PGS (HR per SD increased to 2.3 [2.1-2.4] in the study cohort and 2.1 [1.8-2.4] in the symptomatic subcohort), higher discrimination (C-index was 0.79 [0.78-0.79] in the study cohort and 0.74 [0.73-0.75] in the symptomatic subcohort), and with larger contributions from the PGS to the discriminative ability (46% [35.1%-56%] in study cohort and 43% [11%-69%] in the symptomatic subcohort) (Table S5, Figures S6a-b).

Using any CRC presenting symptoms except fatigue to define the symptomatic individuals resulted in a larger symptomatic subcohort (n=70,231) than in the main analysis. The predictors selected and their performance were almost identical, although with a slightly higher discriminative ability (0.72 [0.71-0.73]) (Table S5, Figure S7).

Excluding 14,743 participants with English Vision primary care records did not change the model discrimination or the ranking of the predictor sets in either the study cohort or symptomatic subcohort (Table S5, Figure S8c), although the symptom set of predictors makes a larger contribution to model performance in the symptomatic subcohort for this sensitivity analysis (15.4% [-19%-50%] compared to 13.2 [-19%-42%] in the main analysis).

## DISCUSSION

### Summary

In this study we have quantified the contributions of polygenic scores, primary care records (including presenting symptoms, medical history and common blood tests) and lifestyle factors for the short-term risk prediction of colorectal cancer (CRC) amongst participants in UK Biobank.

Polygenic scores make substantial contributions to short-term risk prediction for CRC in both a representative sample of UK Biobank participants and a symptomatic sub-cohort (16% [8%-26%] and 8% [-21%-35%] respectively); however, the contribution of information in primary care records (including presenting symptoms, medical history and common blood tests) is greater; the large contribution of common blood tests to the discriminatory ability of the models (32% [19%-43%] and 41% [16%-78%] respectively) is particularly of note. In contrast, the additional lifestyle risk factors make only a small contribution to the model in the study cohort (6% [0%-11%]), and none in the symptomatic subcohort.

This study also presents applications of several cutting-edge methodological approaches to clinical prognostic modelling, including the use of the super-landmark framework (to incorporate longitudinal data from primary care records) and the use of Shapley values to assess the contribution of groups of predictors to model performance (quantifying the contributions of the six predictor sets to overall model discrimination).

### The role of genetics in short-term CRC risk prediction

Several studies have investigated the possibility of adding genetics to other predictors for longer-term CRC risk prediction. Kachuri et al. measure the change in discrimination when a PGS is added to other predictors (demographic, lifestyle and family history) for CRC over a five year follow-up; the C-index [standard error] increases from 0.686 [0.006] to 0.716 [0.006] (35). Briggs et al. showed that adding a PGS to the QCancer-10 model (includes demographic, lifestyle and medical history predictors) increased discrimination for predicted CRC risk, whereas the C-index (in women) increased from 0.65 [0.63-0.66] to 0.69 [0.67-0.70], with authors concluding that there is no clear justification for including a genetic element in risk-stratification for CRC screening (12).

However, in previous studies only the effect of adding a PGS after a phenotypic model had been developed was measured. Evaluation of how genetic information can be used to predict short-term cancer risk has had less attention, although Green et al. found that adding a PGS to age in a symptomatic cohort improved discrimination for prostate cancer from 0.68 [0.65-0.71] to 0.77 [0.74-0.80] for a two year follow-up (17).

In this study, we treat genetic and phenotypic predictors equally, which permits a more objective comparison of their relative contribution. Similar to Briggs et al. (12), a modest improvement in discrimination is seen when a PGS is added to all other included predictors (from 0.71 [0.71-0.72] to 0.73 [0.72-0.74] in the study cohort). However, we demonstrate that genetics makes a substantial contribution to the overall performance of the model (16% [8%-26%] and 8% [-21%-35%] in the study cohort and symptomatic cohort, respectively). In the sensitivity analysis using LDPred, the PGS makes a larger contribution to model performance (46% [35%-56%] in the study cohort) making it the most influential predictor set. LDPred may be better optimised for this cohort (development only included individuals with European ancestry), however, we note that there are risks of overfitting (and hence inflation of the genetic component) due to the use of UK Biobank individuals in LDPred development.

### The role of primary care records in short-term CRC risk prediction

Risk assessment tools using primary care records are well suited for implementation into clinical practice. The C-index of the model that includes the three primary care data predictor sets (presenting symptoms, medical history and common blood tests) was 0.67 [0.66-0.68] in the symptomatic subcohort (or 0.69 [0.68-0.70] when combined with the core predictors).

In particular, we note that the common blood test predictor set makes the second largest contribution after the core predictor set to the discriminatory ability of the models in both the study cohort (32% [19%-43%]) and symptomatic subcohort (41% [16%-78%]). Previous studies have found anaemia (26), raised haemoglobin(36) and inflammatory markers (30,37) to be strongly predictive of diagnosis with colorectal cancer in isolation, with increased levels seen up to 9 months prior to diagnosis (37). Previous models for the early detection of colorectal cancer have included predictors for low haemoglobin levels (13,14), however, they did not consider the wider range of common tests that may indicate the underlying clinical state, for example low ferritin may also indicate iron-deficiency anaemia (details of the derivation of these predictors in Supplementary Methods 2). The large contribution blood tests make to the discriminatory performance of the model demonstrates the value that these common blood tests have for triaging patients for referral in a primary care setting, including as part of a multifactorial risk assessment.

As in other studies (26,36), we find that individual symptoms are strongly predictive of colorectal cancer diagnosis. For example, rectal bleeding has a hazard ratio [95% CI] of 2.7 [2.0-3.7] in the model developed in the study cohort (and 2.0 [1.4-2.9] in the symptomatic subcohort). However, the overall contribution of the symptom predictor set to discriminatory ability is relatively low, especially in the study cohort (3% [-2%-7%]). This may be due to the relatively low incidence of many of these predictors, especially in the study cohort. For example, only 4% of the cohort have a record of rectal bleeding in the two years before any included landmark age. Additionally, the modelling used relatively long-time scales (e.g. two year lookback period and one year intervals between landmark ages); the predictive value of some symptoms over short periods (<1 year), therefore, may not be well captured in this analysis and should be explored further.

### The role of additional lifestyle information in short-term CRC risk prediction

The predictor set “additional lifestyle” included information about patients (collected during UK Biobank baseline assessment) that is not routinely available in primary care records; these include dietary variables (e.g. red meat consumption) and level of education; note that smoking status, which should be routinely collected in primary care, is included in the core predictor set (38). Three (of five) were selected for the model in the study cohort where this predictor set made a modest contribution to model discrimination (6% [0.3%-11%]), however, in the symptomatic subcohort only one was selected and this set made no contribution to model discrimination (-%5 [– 32%-13%]). No increase in discrimination was seen when adding the additional lifestyle predictors to just the core set in the study cohort. Measuring these types of predictors requires data collection via self-reporting (e.g. questionnaire or interview) which is resource-intensive, prone to recall bias (39,40) and would require further development of suitable data collection methods with routine clinical practise(41); given their relatively small contribution to the discriminative ability of the models in this study, this may not be an efficient use of resources when considering implementation of a multifactorial risk assessment for CRC early detection, especially in a symptomatic primary care population.

However, we note that the data for this group of modifiable risk factors were measured during baseline assessment, but were used (within the superlandmark framework) at index dates up to 10 years after initial assessment. Therefore, estimates of the contribution of these predictors may have been affected by our inability to dynamically update them.

### Strengths and Limitations

Within this study, we have implemented a range of cutting-edge methods to make the best use of available data and address the challenges of prognostic modelling for the early detection of cancer. The super-landmark framework allowed us to maximise the number of cases included in the analysis, include information from across the whole study period (from baseline assessment to the end of primary care linkage) in model development and enabled the dynamic updating of predictors from electronic health records. The use of Shapley values to measure the inclusion-order-agnostic contribution of the predictor sets is a novel approach in the context of prognostic modelling for clinical outcomes.

Additionally, the characteristics of the UKB cohort – including its size, extensive data collection (including genotyping) and linkage (to national registries and primary care records) – has made the analysis described in this study possible. The use of linked primary care records has permitted the derivation of predictors describing a range of clinical events (symptoms, medical history and common blood tests). All codelists used to identify predictors of interest from primary care records were harmonised between the four coding frameworks used by the data providers, and newly developed codelists were checked by clinical experts.

However, there are several limitations to these analyses. First, UKB participants are not representative (42); with only a small number reporting non-white ethnicity (5.0% in the study cohort) and lower cancer incidence and fewer symptoms than the UK general population (43). A recent analysis found that cancer symptom reporting was lower in UKB participants than in a more representative primary care population (44). Another study has claimed that some associations between predictors and outcomes may be consistent between UKB and the general population although differences were seen for some outcomes (45), however, this has not yet been explored for cancer incidence. In fact, measured discrimination of the models may be higher in the general population due to greater heterogeneity of some risk predictors (46). External validation in a more representative population would be required to demonstrate generalisability, however, there are currently no similar datasets that include both detailed primary care records and genetic data, although this may change in the future.

Second, primary care records do not give a comprehensive or unbiased view of the health status of cohort members, given the requirement for patients to seek healthcare and for a clinician to code any event of interest (we note that much information is primary care is recorded as free text which is never made available for research). Coding practice in UK primary care is known to vary by data provider (47,48) and over time (48), which may explain the variation seen in the results (symptomatic subcohort only) in the sensitivity analysis excluding participants with English Vision records. Some promising predictors (such as faecal immunochemical tests (49)) were not included in the analysis due to low counts in the available primary care records (linkage ends 2016-2018 before widespread use of this test in clinical practice).

## Conclusions

We have quantified the contributions of six predictor sets to the discrimination of dynamic models predicting the short-term risk of CRC in both a study cohort of UK Biobank participants registered with a primary care provider and a symptomatic subcohort. We included a range of predictors (and data) types – including demographics, genetics, symptoms, medical history, primary care blood tests and lifestyle – and assessed their contributions using an order-agnostic method. We evidence the meaningful independent contribution of genetics and data from primary care records, especially common blood tests, to identify people at high risk of CRC diagnosis in the near future. In comparison, only a small contribution is measured for additional lifestyle risk factors which are not routinely collected in primary care.

We anticipate that the resources developed for this study (available online) – including the novel application of the super-landmark framework (incorporating longitudinal data from primary care records) and Shapley values to clinical prognostic modelling – will be of interest to researchers working with electronic health records.

#### What is already known on this topic

● Associations between colorectal cancer diagnosis and a range of individual level characteristics – including demographics, lifestyle, polygenic scores, symptoms, medical history, and primary care blood tests – are well-established.
● Models using a range of predictor sets offer a way to identify people at high short-term risk of a colorectal cancer diagnosis, but the relative contribution of the different predictor sets to overall model performance is unclear.
● Polygenic scores for colorectal cancer are promising for long-term follow-up (5 or more years), but have not yet been explored for short-term follow-up.

#### What this study adds

● Quantifies the contribution of six predictor sets to the short-term prediction of colorectal cancer using a novel application of Shapley values to health care data in both a general and symptomatic cohort.
● Demonstrates the substantial contribution of polygenic scores and data from primary care records (particularly common blood tests) to models for the early detection of colorectal cancer.
● Uses a super-landmark framework to incorporate longitudinal data from primary care records into a robust analysis pipeline for model development and validation.

## Ethics approval and consent to participate

The UK Biobank study was approved by the North West Multi-Centre Research Ethics Committee (reference number 06/MRE09/ 65), and at recruitment all participants gave informed written consent to participate in UK Biobank and be followed up, using a signature capture device.

Data storage and access for this project was strictly controlled in accordance with the data agreements between the researchers and UK Biobank. Access to this data was approved by UK Biobank under projects 64351 (Real-world Risk-stratified Early Detection and Diagnosis using linked Electronic Health Records (RREDD-EHR)) and 28126 (Validating risk prediction models for common hormonal cancers). Throughout the project the data were stored and accessed only via a secure server at the Centre for Cancer Genetic Epidemiology (DPHPC, University of Cambridge). This work uses data provided by patients and collected by the NHS as part of their care and support.

## Availability of data and materials

This research has been conducted using the UK Biobank Resource under Application Number 64351.

This paper uses data from UK Biobank that the authors do not have permission to distribute. Bona-fide researchers can apply for access to this data, including linked primary care records, from UK Biobank https://www.ukbiobank.ac.uk/

Resources developed for this project including codelists and analysis code can be found at [TBC].

## Data Availability

This paper uses data from UK Biobank that the authors do not have permission to distribute. Bona-fide researchers can apply for access to this data, including linked primary care records, from UK Biobank https://www.ukbiobank.ac.uk/ Resources developed for this project including codelists and analysis code can be found at [TBC GitHub link].

