## Appendix 1 for "Genetics, primary care records and lifestyle factors for short-term dynamic risk prediction of colorectal cancer: prospective study of asymptomatic and symptomatic UK Biobank participants"

Supplementary Material

Supplementary Figure 3: Age and sex-adjusted hazard ratios for symptoms selected by Cox PH bidirectional stepwise selection from the "symptoms" predictor set (Table S1), defining our "symptomatic" subcohort. . 31

Supplementary Figure 6b: Figure 6b-a Hazard ratios from bidirectional stepwise Cox regression for the symptomatic subcohort, using LDPred-generated polygenic scores; Figure 6b-b The inclusion-order-agnostic discriminative contribution (C-index > 0.5) of each predictor set evaluated using Shapley values; Figure 6b-c C-indices from 200 bootstrap samples for each coalition of predictor sets. Colour-coding indicates the predictor set in all figures............................................................................................................................ 35

Supplementary Figure 7: Figure 7a Hazard ratios from bidirectional stepwise Cox regression for the "symptomatic" subcohort (N=70,241) defined as having any symptom in the predictor type "symptoms" sans fatigue; Figure 7b The inclusion-order-agnostic discriminative contribution (C-index > 0.5) of each predictor set evaluated using Shapley values; Figure 7c C-indices from 200 bootstrap samples for each coalition of predictor sets. Colour-coding indicates the predictor set in all figures. ...................................... 36

Supplementary Methods

*Supplementary Methods 1:* ***Super-landmark framework***

We structured the cohort into landmark age datasets at landmark ages 40, 41, 42, … up to 74 years (1,2). Participants were included in a landmark age dataset if they were alive, had at least six months of continuous primary care records (no gaps >90 days; where multiple continuous periods are available we used the most recent) in the previous two years and had not previously received a cancer diagnosis (except non-melanoma skin cancer) before the landmark age. The date when a participant entered a landmark age dataset (1st of the month in which they reach a landmark age) is referred to as the “index date”. Time-varying predictors and outcomes (defined below) were extracted for each participant at all their index dates. We censored follow-up in each landmark dataset at the earliest occurrence of several events, including the occurrence of any incident cancer excluding non-melanoma skin cancer, death, the end of GP records, or two years since the landmark age. Each individual was eligible to contribute to multiple landmark ages during their time at risk, accounted for using robust variance estimation. The ability for participants to contribute to the analysis at multiple time points (in contrast with the conventional single-time-point contribution of participants to a model) optimises the utilisation of the available longitudinal data from primary care records and maximises the number of incident CRC cases[1] included in the analysis.

The survival model derived from this stacked dataset assumed that the baseline hazards at different landmark ages varied only by an adjustment for landmark age and its square:

$$h(t|X(t_*), t_*) = h_0(t)\exp(\beta_0^T X(t_*) + \theta_1 t_* + \theta_2 t_*^2),$$

where $h(t|X(t_*), t_*)$is the hazard function, $h_0(t)$is the baseline hazard, $t_*$is the landmark age, $X(t_*)$is a vector of predictor values at landmark age $t_*$, and $\beta_0, \theta_1, \theta_2$are the log-Hazard Ratios associated with the predictors.

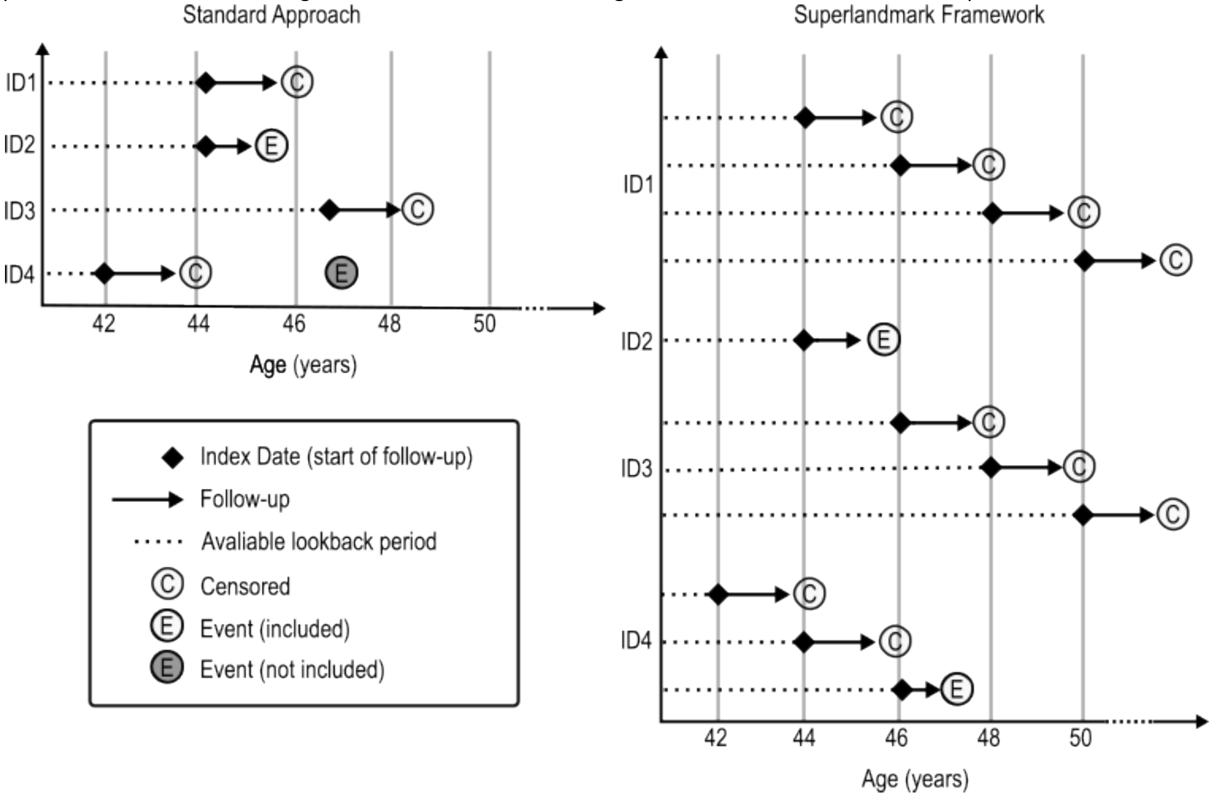

1 Static models underutilised available outcome data (cancer diagnoses), as they allow only one index date per individual, e.g. baseline assessment. Any cancer diagnoses that occur outside the predefined 2-year window would not be included in the model. This is especially problematic in the study of rarer cancers, where we face challenges in statistical power and potential overfitting. It's important to note that the choice of the index date should be independent of the outcome (cancer diagnosis) to avoid introducing biases into the model. On the other hand, dynamic modelling is more flexible, allowing the inclusion of outcomes as they occur throughout the study period.

*Supplementary Methods 2:* ***Dataset and Variable Derivation Details***

For comprehensive information on the dataset used and the methodology for variable derivation in this study, please refer to Appendix 2.

*Supplementary Methods 3:* ***Cox PH model and model selection***

For the survival analysis, we derived Cox Proportional Hazards (Cox PH) models on a superlandmark dataset, using robust standard error estimation. To ensure model parsimony, we applied three model selection methods:

- No selection: Including all defined predictors.
- Bidirectional stepwise selection (3–5): A method that iteratively adds and removes predictors based on Akaike Information Criterion (AIC).
- Group Lasso 1se (6–10): Lasso (Least Absolute Shrinkage and Selection Operator) is a regularisation method that improves model parsimony by selecting a subset of predictors. The "1se" method chooses the regularisation parameter, $\lambda$, as the largest value within one standard error of the minimum cross-validated error, resulting in a simpler model with fewer predictors. Group Lasso extends this by selecting groups of predictors, ensuring either all or none of a categorical variable's levels are included. In our analysis, we accounted for patient IDs during the internal cross-validation for lambda selection to prevent data leakage between internal training and validation sets.

We compared these methods by evaluating their C-indices through 200 bootstrap samples and examining the consistency of the selected predictors across the different methods.

*Supplementary Methods 4:* ***Shapley values***

We calculated the Shapley values (11) of the C-index metric to determine the contribution of each predictor group to the model's discriminative performance, independent of the order of inclusion. We defined six predictor groups: Core, Polygenic score, symptoms, primary care investigations, medical history, and lifestyle; Table S1. Using 200 bootstrap samples, we derived models using only the predictors in each subgroup and presented the C-indices in a violin plot. We calculated the contribution of each combination of predictor groups (including each group alone) to the C-index and presented the contribution of each predictor group above random in a bar chart (Figure 1 and 2) . The Shapley values of the C-index-0.5 for each predictor group were calculated using:

$$\Phi_p = \sum_{S \subseteq P \setminus \{p\}} \frac{|S|!(|P|-|S|-1)!}{|P|!}(\nu(S \cup \{p\}) - \nu(S))$$

,

where
$p \in P = \{\text{Core}, \text{Polygenic score}, \text{Symptoms}, \text{Primary care investigations}, \text{Medical history}, \text{Lifestyle}\}$
is a predictor group in the set of six predictor groups $P$, $S \subseteq P$ is a subset of predictor groups, $\Phi_p$ is the Shapley value of the C-index for predictor group $p$, $\nu(S)$ is the C-index when using only the predictors in $S$.

Comparison with incremental added value approach

This method of value assignment places the predictor groups on equal footing by calculating the average marginal contribution of each group across all possible combinations. This contrasts with the traditional incremental value approach, which evaluates performance gains when adding a predictor group to a model containing all others.

The incremental value approach and the Shapley value approach answer two different questions.

The incremental value approach asks: What is the additional discriminative power gained by adding a novel predictor to existing ones? This approach is highly order-dependent, prioritising existing predictors and may undervalue predictors added later.

Whereas, the Shapley value approach asks: What is each predictor's contribution to overall model discrimination, considering all combinations? This approach calculates average marginal contributions across all possible orders, allowing predictor types to be on equal footing and understanding their individual contributions and interactions with other predictors.

Comparison with SHAP or machine learning interpretability methods

In conventional machine learning interpretability with SHAP (12), the focus is on breaking down a model's prediction to see how individual features contribute to specific predictions. SHAP asks: What is the influence of individual predictors on model predictions? For example, how much does being male contribute to individual A's predicted risk of CRC? These contributions are similar to hazard ratios in our model.

In contrast, our approach using Shapley values applied to C-indices shifts from explaining individual predictions to evaluating the overall model performance attributed to each predictor group. This approach evaluates the discriminative power of predictor groups, asking: What is the contribution of each predictor group to overall model discrimination? For example, how much does a polygenic risk score (PGS) contribute to the overall model's C-index gains?

*Supplementary Methods 5:* ***TRIPOD Checklist: Prediction Model Development and Validation:*** *While internal-validation was included, validation is not the main focus of this study, rather we wanted to quantify the contributions of different predictor sets.*

| **Section/Topic** | | | **Checklist Item** | **Page** |
|---|---|---|---|---|
| **Title and abstract** | | | | |
| Title | 1 | ;V | Identify the study as developing and/or validating a multivariable prediction model, the target population, and the outcome to be predicted. | 1 |
| Abstract | 2 | ;V | Provide a summary of objectives, study design, setting, participants, sample size, predictors, outcome, statistical analysis, results, and conclusions. | 2 |
| **Introduction** | | | | |
| Background and objectives | a | ;V | Explain the medical context (including whether diagnostic or prognostic) and rationale for developing or validating the multivariable prediction model, including references to existing models. | 3 |
| | b | ;V | Specify the objectives, including whether the study describes the development or validation of the model or both. | 3 |
| **Methods** | | | | |
| Source of data | a | ;V | Describe the study design or source of data (e.g., randomized trial, cohort, or registry data), separately for the development and validation data sets, if applicable. | 4 |
| | b | ;V | Specify the key study dates, including start of accrual; end of accrual; and, if applicable, end of follow-up. | 4 |
| Participants | a | ;V | Specify key elements of the study setting (e.g., primary care, secondary care, general population) including number and location of centres. | 4 |
| | b | ;V | Describe eligibility criteria for participants. | 4 |
| | c | ;V | Give details of treatments received, if relevant. | N/A |
| Outcome | a | ;V | Clearly define the outcome that is predicted by the prediction model, including how and when assessed. | 4 |
| | b | ;V | Report any actions to blind assessment of the outcome to be predicted. | N/A |
| Predictors | a | ;V | Clearly define all predictors used in developing or validating the multivariable prediction model, including how and when they were measured. | 5,<br>S-Mthds 2,<br>Appendix 2 |
| | b | ;V | Report any actions to blind assessment of predictors for the outcome and other predictors. | N/A |
| Sample size | 8 | ;V | Explain how the study size was arrived at. | 4,<br>S-Fig 1 |
| Missing data | 9 | ;V | Describe how missing data were handled (e.g., complete-case analysis, single imputation, multiple imputation) with details of any imputation method. | 5 |
| Statistical analysis methods | 0a | D | Describe how predictors were handled in the analyses. | 5 |
| | 0b | D | Specify type of model, all model-building procedures (including any predictor selection), and method for internal validation. | 4-6<br>S-Mthds 1-4 |
| | 0c | V | For validation, describe how the predictions were calculated. | 5-6,<br>S-Mthds 3-4 |
| | 0d | ;V | Specify all measures used to assess model performance and, if relevant, to compare multiple models. | 5-6 |
| | 0e | V | Describe any model updating (e.g., recalibration) arising from the validation, if done. | N/A |

| | | | | |
|---|---|---|---|---|
| Risk groups | 1 | ;V | Provide details on how risk groups were created, if done. | N/A |
| Development vs. validation | 2 | V | For validation, identify any differences from the development data in setting, eligibility criteria, outcome, and predictors. | N/A |
| **Results** | | | | |
| Participants | 3a | ;V | Describe the flow of participants through the study, including the number of participants with and without the outcome and, if applicable, a summary of the follow-up time. A diagram may be helpful. | S-Fig 1 |
| | 3b | ;V | Describe the characteristics of the participants (basic demographics, clinical features, available predictors), including the number of participants with missing data for predictors and outcome. | 14 (Tbl 1), S-Tbls 4,7 Appendix 2 |
| | 3c | V | For validation, show a comparison with the development data of the distribution of important variables (demographics, predictors and outcome). | N/A |
| Model development | 4a | D | Specify the number of participants and outcome events in each analysis. | S-Tbl 5 |
| | 4b | D | If done, report the unadjusted association between each candidate predictor and outcome. | N/A |
| Model specification | 5a | D | Present the full prediction model to allow predictions for individuals (i.e., all regression coefficients, and model intercept or baseline survival at a given time point). | 16, 17, S-Tbls 2a,2b,6 S-Figs 6a,6b,7a,8b,8c |
| | 5b | D | Explain how to use the prediction model. | S-Tables (1-2), S-Mthds /Appendix 2 |
| Model performance | 6 | ;V | Report performance measures (with CIs) for the prediction model. | 8-9,17-18 S-Tbls 3a,3b,5 S-Fig 4 |
| Model-updating | 7 | V | If done, report the results from any model updating (i.e., model specification, model performance). | N/A |
| **Discussion** | | | | |
| Limitations | 8 | ;V | Discuss any limitations of the study (such as nonrepresentative sample, few events per predictor, missing data). | 10-11 |
| Interpretation | 9a | V | For validation, discuss the results with reference to performance in the development data, and any other validation data. | N/A |
| | 9b | ;V | Give an overall interpretation of the results, considering objectives, limitations, results from similar studies, and other relevant evidence. | 9-12 |
| Implications | 0 | ;V | Discuss the potential clinical use of the model and implications for future research. | 9-12 |
| **Other information** | | | | |
| Supplementary information | 1 | ;V | Provide information about the availability of supplementary resources, such as study protocol, Web calculator, and data sets. | 13 |
| Funding | 2 | ;V | Give the source of funding and the role of the funders for the present study. | 13 |

*Supplementary Methods 6:* ***Functional Specification of Age***

We used a quadratic model incorporating age and age-squared terms to capture the nonlinear increase in cancer risk for individuals aged 40-74. This functional form has been selected in previous published work to capture this association (8), avoiding unnecessary complexity and minimising the potential for overfitting within this relatively narrow age range. Additionally, use of this relatively simple form of age enhances the interpretability of the final model.

Supplementary Tables

***Supplementary Table 1:*** *Description of predictors included in each of the six predictor groups. Italic predictors were considered but excluded from modelling due to low counts in the UKBiobank cohort.*[2]

| | Predictors | Data Source | Available in Primary Care? |
|---|---|---|---|
| **Core** | Age, Birth year, Sex, Ethnicity, Body Mass Index, Smoking status, Townsend deprivation score | UKB baseline | Yes |
| **Additional Lifestyle** | Red meat consumption, Processed meat consumption, Fibre consumption score, Education (highest qualification), Alcohol consumption | UKB baseline | No |
| **Symptoms (recorded)*** | Abdominal bloating (recent), Abdominal pain, Rectal bleeding, Change in bowel habit, Stomach disorders, Fatigue (new-onset), Diverticular disease (recent diagnosis), Irritable Bowel Syndrome (recent diagnosis), Constipation (new-onset), Diarrhoea (new-onset), Haemorrhoids (new-onset)<br>Excluded due to low counts: *Abdominal lump (recent), Pelvic pain (recent), Weight loss (new-onset), Jaundice (new-onset), Rectal mass (recent)* | EHR (primary care and prescription) | Yes |
| **Medical history (recorded)** | Family history of bowel cancer, Family history of breast cancer, Family history of lung cancer, Eligibility for bowel cancer screening, Colonoscopy (in last 10 years), Inflammatory bowel disease, Diabetes (Type 2), Gallstones, Multi-morbidity score, Regular use of non-aspirin NSAIDs, Regular use of aspirin<br>Excluded due to low counts: *Type I diabetes* | EHR (primary care and prescription) | Yes |
| **Primary care blood tests (recorded)** | Iron deficiency anaemia (measured or abnormal), inflammation (measured or abnormal) | EHR (primary care) | Yes |
| **Polygenic score** | Polygenic score (PRS-CSx), Principal genetic components (1-10) | UKB genotyping | No |

[2] See appendix 2 for details of symptom derivation

***Supplementary Table 2a:*** *Hazard ratios of risk predictors selected using Cox regression with three different variable selection approaches (no selection, bidirectional selection , and lasso) in the study cohort.*[3]

| Predictor | Predictor level or units[4] | HR (95% CI) |
|---|---|---|
| **Cox PH without selection** | | |
| Age | 8.14 years | 5.649 (2.031, 15.710) |
| Age-squared | 975.00 years$^2$ | 0.222 (0.086, 0.573) |
| Birth year | 7.90 years | 0.631 (0.533, 0.747) |
| BMI | 4.75 kg/m$^2$ | 1.063 (1.005, 1.125) |
| Ethnicity | SE Asian | 1.476 (0.350, 6.228) |
| | Black | 0.760 (0.207, 2.782) |
| | Mixed | 1.946 (0.695, 5.448) |
| | White | 1.016 (0.327, 3.157) |
| | Missing | 1.006 (0.251, 4.033) |
| Sex (genetic) | Male | 1.607 (1.423, 1.815) |
| Smoking status | Current | 1.162 (0.957, 1.412) |
| | Previous | 1.145 (1.017, 1.289) |
| | Missing | 0.499 (0.150, 1.661) |
| Townsend deprivation score | 2.98 | 1.014 (0.956, 1.076) |
| Alcohol units daily | 2.89 units/day | 1.119 (1.085, 1.154) |
| Education | A-level | 0.855 (0.694, 1.054) |
| | GCSE | 0.992 (0.848, 1.161) |
| | CSE | 0.953 (0.706, 1.285) |
| | Vocational | 0.980 (0.787, 1.221) |
| | Professional | 0.876 (0.679, 1.130) |
| | None | 0.898 (0.761, 1.060) |
| Fibre consumption | 6.37 units/day | 0.966 (0.912, 1.023) |
| Processed meat consumption | 1.39 servings/week | 1.029 (0.973, 1.088) |
| Red meat consumption | 1.45 servings/week | 1.027 (0.972, 1.086) |
| Abdominal bloating | True | 1.027 (0.577, 1.829) |
| Abdominal pain | True | 0.965 (0.764, 1.219) |
| Change in Bowel habits | True | 1.414 (0.841, 2.377) |
| Diverticular | True | 0.852 (0.529, 1.371) |
| Fatigue | True | 0.734 (0.503, 1.072) |
| IBS | True | 1.228 (0.667, 2.261) |
| Rectal bleed | True | 2.749 (2.038, 3.709) |
| Stomach disorders | True | 0.851 (0.514, 1.408) |
| Constipation | True | 1.204 (0.986, 1.471) |
| Diarrhoea | True | 0.865 (0.673, 1.110) |
| Haemorrhoids | True | 1.517 (1.169, 1.969) |
| Diabetes T2 ever | True | 1.154 (0.940, 1.416) |
| Gallbladder calc ever | True | 1.098 (0.818, 1.474) |
| IBD ever | True | 0.935 (0.618, 1.413) |
| Aspirin | True | 0.840 (0.640, 1.102) |
| NSAIDs (non-aspirin) | True | 0.744 (0.629, 0.880) |
| Eligible for bowel cancer screening | True | 0.901 (0.774, 1.050) |
| Colonoscopy in last 10 years | True | 0.593 (0.504, 0.699) |
| Family history of bowel cancer | True | 1.183 (1.009, 1.388) |
| Family history of breast cancer | True | 1.030 (0.861, 1.233) |
| Family history of lung cancer | True | 1.074 (0.918, 1.256) |
| Multimorbidity Score (residual) | 0.39 | 0.931 (0.877, 0.989) |

[3] * indicates no CRC cases with that risk predictor level.

[4] Continuous variables are standardised, with their standard deviations presented here. Hazard Ratios (HRs) are interpreted per standard deviation increase.

| | | |
|---|---|---|
| Inflammation | Abnormal | 1.731 (1.530, 1.958) |
| | Measured | 1.311 (0.990, 1.735) |
| Iron deficiency | Abnormal | 3.945 (3.404, 4.571) |
| | Measured | 1.091 (0.838, 1.420) |
| Polygenic Score | 0.09 | 1.403 (1.325, 1.486) |
| **Cox PH with backwards/forwards stepwise selection** | | |
| Age | 8.14 years | 5.750 (2.071, 15.964) |
| Age-squared | 975.00 years² | 0.218 (0.085, 0.563) |
| Birth year | 7.90 years | 0.637 (0.539, 0.753) |
| BMI | 4.75 kg/m² | 1.064 (1.006, 1.126) |
| Sex (genetic) | Male | 1.608 (1.426, 1.813) |
| Smoking status | Current | 1.163 (0.962, 1.407) |
| | Previous | 1.145 (1.017, 1.288) |
| | Missing | 0.489 (0.147, 1.625) |
| Alcohol units daily | 2.89 units/day | 1.122 (1.089, 1.156) |
| Fibre consumption | 6.37 units/day | 0.967 (0.913, 1.024) |
| Processed meat consumption | 1.39 servings/week | 1.034 (0.980, 1.091) |
| Fatigue | True | 0.731 (0.501, 1.067) |
| Rectal bleed | True | 2.731 (2.023, 3.688) |
| Constipation | True | 1.202 (0.985, 1.466) |
| Haemorrhoids | True | 1.514 (1.167, 1.963) |
| Diabetes T2 ever | True | 1.158 (0.944, 1.421) |
| Aspirin | True | 0.837 (0.638, 1.098) |
| NSAIDs (non-aspirin) | True | 0.743 (0.629, 0.878) |
| Eligible for bowel cancer screening | True | 0.902 (0.775, 1.051) |
| Colonoscopy in last 10 years | True | 0.590 (0.502, 0.694) |
| Family history of bowel cancer | True | 1.188 (1.013, 1.394) |
| Multimorbidity Score (residual) | 0.39 | 0.927 (0.873, 0.983) |
| Inflammation | Abnormal | 1.730 (1.530, 1.956) |
| | Measured | 1.399 (1.129, 1.733) |
| Iron deficiency | Abnormal | 3.948 (3.409, 4.572) |
| Polygenic Score | 0.09 | 1.403 (1.325, 1.485) |
| **LASSO Cox regression** | | |
| Age | 8.14 years | 5.658 (2.034, 15.740) |
| Age-squared | 975.00 years² | 0.222 (0.086, 0.573) |
| Birth year | 7.90 years | 0.631 (0.533, 0.748) |
| BMI | 4.75 kg/m² | 1.064 (1.006, 1.126) |
| Ethnicity | SE Asian | 1.477 (0.350, 6.236) |
| | Black | 0.759 (0.207, 2.781) |
| | Mixed | 1.946 (0.695, 5.448) |
| | White | 1.015 (0.327, 3.152) |
| | Missing | 1.008 (0.251, 4.039) |
| Sex (genetic) | Male | 1.602 (1.419, 1.809) |
| Smoking status | Current | 1.162 (0.956, 1.412) |
| | Previous | 1.145 (1.017, 1.289) |
| | Missing | 0.499 (0.150, 1.658) |
| Townsend deprivation score | 2.98 | 1.014 (0.956, 1.076) |
| Alcohol units daily | 2.89 units/day | 1.119 (1.085, 1.154) |
| Education | A-level | 0.855 (0.694, 1.055) |
| | GCSE | 0.993 (0.848, 1.162) |
| | CSE | 0.953 (0.707, 1.285) |
| | Vocational | 0.980 (0.787, 1.221) |
| | Professional | 0.876 (0.680, 1.130) |
| | None | 0.898 (0.761, 1.060) |
| Fibre consumption | 6.37 units/day | 0.966 (0.912, 1.023) |

| | | |
|---|---|---|
| Processed meat consumption | 1.39 servings/week | 1.029 (0.973, 1.088) |
| Red meat consumption | 1.45 servings/week | 1.027 (0.972, 1.085) |
| Abdominal bloating | True | 1.029 (0.578, 1.831) |
| Abdominal pain | True | 0.969 (0.767, 1.224) |
| Change in Bowel habits | True | 1.416 (0.842, 2.380) |
| Diverticular | True | 0.853 (0.530, 1.372) |
| Fatigue | True | 0.735 (0.504, 1.073) |
| IBS | True | 1.230 (0.668, 2.267) |
| Rectal bleed | True | 2.750 (2.038, 3.711) |
| Stomach disorders | True | 0.852 (0.515, 1.410) |
| Constipation | True | 1.206 (0.987, 1.473) |
| Diarrhoea | True | 0.864 (0.673, 1.110) |
| Haemorrhoids | True | 1.518 (1.169, 1.971) |
| Diabetes T2 ever | True | 1.156 (0.942, 1.418) |
| Aspirin | True | 0.838 (0.639, 1.099) |
| NSAIDs (non-aspirin) | True | 0.745 (0.630, 0.880) |
| Eligible for bowel cancer screening | True | 0.902 (0.774, 1.050) |
| Colonoscopy in last 10 years | True | 0.593 (0.503, 0.699) |
| Family history of bowel cancer | True | 1.184 (1.009, 1.389) |
| Family history of breast cancer | True | 1.030 (0.861, 1.233) |
| Family history of lung cancer | True | 1.073 (0.917, 1.256) |
| Multimorbidity Score (residual) | 0.39 | 0.931 (0.877, 0.989) |
| Inflammation | Abnormal | 1.737 (1.536, 1.964) |
| | Measured | 1.402 (1.131, 1.737) |
| Iron deficiency | Abnormal | 3.960 (3.417, 4.588) |
| Polygenic Score | 0.09 | 1.403 (1.325, 1.486) |

***Supplementary Table 2b:*** *Hazard ratios of risk predictors selected using Cox regression with three different variable selection approaches (no selection, bidirectional selection , and lasso) in the symptomatic subcohort.*[5]

| Predictor | Predictor level or units[6] | HR (95% CI) |
|---|---|---|
| **Cox PH without selection** | | |
| Age | 8.02 years | 0.826 (0.063, 10.748) |
| Age-squared | 968.90 years² | 1.277 (0.116, 14.073) |
| Birth year | 7.78 years | 0.603 (0.388, 0.935) |
| BMI | 5.03 kg/m² | 1.160 (1.012, 1.331) |
| Ethnicity | SE Asian | 0.439 (0.035, 5.491) |
| | Black | 0.623 (0.073, 5.302) |
| | Mixed | 2.183 (0.199, 23.954) |
| | White | 0.745 (0.050, 11.196) |
| | * Missing | 0.000 (0.000, 0.000) |
| Sex (genetic) | Male | 1.989 (1.459, 2.711) |
| Smoking status | Current | 1.025 (0.617, 1.704) |
| | * Missing | 0.000 (0.000, 0.000) |
| | Previous | 1.198 (0.893, 1.608) |
| Townsend deprivation score | 3.13 | 1.079 (0.929, 1.253) |
| Alcohol units daily | 2.87 units/day | 1.006 (0.888, 1.140) |
| Education | A-level | 0.697 (0.395, 1.230) |
| Education | GCSE | 0.883 (0.601, 1.295) |
| Education | CSE | 1.088 (0.596, 1.984) |
| Education | Vocational | 0.418 (0.217, 0.808) |
| Education | Professional | 0.844 (0.461, 1.546) |
| Education | None | 0.539 (0.354, 0.819) |
| Fibre consumption | 6.64 units/day | 0.959 (0.843, 1.091) |
| Processed meat consumption | 1.40 servings/week | 0.981 (0.849, 1.134) |
| Red meat consumption | 1.45 servings/week | 1.004 (0.868, 1.162) |
| Abdominal bloating | True | 0.185 (0.026, 1.304) |
| Abdominal pain | True | 0.924 (0.601, 1.421) |
| Change in Bowel habits | True | 0.624 (0.182, 2.137) |
| Diverticular | True | 0.630 (0.348, 1.139) |
| Fatigue | True | 0.569 (0.268, 1.208) |
| IBS | True | 1.025 (0.452, 2.327) |
| Rectal bleed | True | 1.978 (1.341, 2.917) |
| Stomach disorders | True | 1.276 (0.562, 2.900) |
| Constipation | True | 0.764 (0.514, 1.137) |
| Diarrhoea | True | 1.147 (0.751, 1.750) |
| Haemorrhoids | True | 1.005 (0.711, 1.420) |
| Diabetes T2 ever | True | 0.857 (0.520, 1.413) |
| Gallbladder calc ever | True | 1.028 (0.510, 2.074) |
| IBD ever | True | 0.678 (0.260, 1.769) |
| Aspirin | True | 0.719 (0.399, 1.293) |
| NSAIDs (non-aspirin) | True | 0.677 (0.473, 0.968) |
| Eligible for bowel cancer screening | True | 1.163 (0.758, 1.782) |
| Colonoscopy in last 10 years | True | 0.539 (0.398, 0.730) |
| Family history of bowel cancer | True | 1.198 (0.805, 1.783) |
| Family history of breast cancer | True | 0.935 (0.606, 1.441) |
| Family history of lung cancer | True | 0.877 (0.572, 1.345) |
| Multimorbidity Score (residual) | 0.50 | 0.889 (0.755, 1.047) |

[5] * indicates no CRC cases with that risk predictor level.

[6] Continuous variables are standardized, with their standard deviations presented here. Hazard Ratios (HRs) are interpreted per standard deviation increase.

| | | |
|---|---|---|
| Inflammation | Abnormal | 1.523 (1.133, 2.047) |
| | Measured | 3.903 (1.306, 11.664) |
| Iron deficiency | Abnormal | 4.016 (2.881, 5.599) |
| | Measured | 0.703 (0.298, 1.662) |
| Polygenic Score | 0.09 | 1.326 (1.157, 1.520) |
| **Cox PH with backwards/forwards stepwise selection** | | |
| Age | 8.02 years | 0.989 (0.076, 12.870) |
| Age-squared | 968.90 years² | 1.165 (0.103, 13.159) |
| Birth year | 7.78 years | 0.623 (0.416, 0.932) |
| BMI | 5.03 kg/m² | 1.160 (1.012, 1.328) |
| Sex (genetic) | Male | 2.031 (1.535, 2.688) |
| Education | A-level | 0.700 (0.398, 1.234) |
| | GCSE | 0.893 (0.612, 1.304) |
| | CSE | 1.117 (0.617, 2.025) |
| | Vocational | 0.425 (0.222, 0.813) |
| | Professional | 0.854 (0.470, 1.553) |
| | None | 0.552 (0.370, 0.822) |
| Abdominal bloating | True | 0.186 (0.026, 1.312) |
| Diverticular | True | 0.624 (0.361, 1.079) |
| Fatigue | True | 0.570 (0.269, 1.208) |
| Rectal bleed | True | 1.962 (1.350, 2.852) |
| Constipation | True | 0.762 (0.556, 1.045) |
| Aspirin | True | 0.715 (0.399, 1.283) |
| NSAIDs (non-aspirin) | True | 0.690 (0.484, 0.984) |
| Colonoscopy in last 10 years | True | 0.534 (0.396, 0.722) |
| Multimorbidity Score (residual) | 0.50 | 0.885 (0.759, 1.032) |
| Inflammation | Abnormal | 1.512 (1.125, 2.032) |
| | Measured | 2.858 (1.203, 6.792) |
| Iron deficiency | Abnormal | 3.985 (2.867, 5.539) |
| Polygenic Score | 0.09 | 1.331 (1.160, 1.527) |
| **LASSO Cox regression** | | |
| Age | 8.02 years | 0.828 (0.064, 10.762) |
| Age-squared | 968.90 years² | 1.275 (0.116, 14.043) |
| Birth year | 7.78 years | 0.603 (0.389, 0.935) |
| BMI | 5.03 kg/m² | 1.161 (1.012, 1.331) |
| Ethnicity | SE Asian | 0.438 (0.035, 5.496) |
| | Black | 0.622 (0.073, 5.305) |
| | Mixed | 2.183 (0.199, 23.916) |
| | White | 0.744 (0.050, 11.159) |
| | * Missing | 0.000 (0.000, 0.000) |
| Sex (genetic) | Male | 1.987 (1.458, 2.707) |
| Smoking status | Current | 1.025 (0.617, 1.704) |
| | Previous | 1.198 (0.893, 1.608) |
| | * Missing | 0.000 (0.000, 0.000) |
| Townsend deprivation score | 3.13 | 1.079 (0.928, 1.253) |
| Alcohol units daily | 2.87 units/day | 1.006 (0.888, 1.140) |
| Education | A-level | 0.697 (0.394, 1.231) |
| | GCSE | 0.883 (0.601, 1.295) |
| | CSE | 1.087 (0.596, 1.983) |
| | Vocational | 0.418 (0.217, 0.808) |
| | Professional | 0.844 (0.461, 1.545) |
| | None | 0.539 (0.354, 0.819) |
| Fibre consumption | 6.64 units/day | 0.959 (0.842, 1.091) |
| Processed meat consumption | 1.40 servings/week | 0.981 (0.849, 1.134) |

| | | |
|---|---|---|
| Red meat consumption | 1.45 servings/week | 1.004 (0.868, 1.161) |
| Abdominal bloating | True | 0.185 (0.026, 1.302) |
| Abdominal pain | True | 0.925 (0.604, 1.417) |
| Change in Bowel habits | True | 0.624 (0.183, 2.136) |
| Diverticular | True | 0.630 (0.348, 1.139) |
| Fatigue | True | 0.569 (0.268, 1.208) |
| Rectal bleed | True | 1.977 (1.340, 2.918) |
| Stomach disorders | True | 1.276 (0.562, 2.900) |
| Constipation | True | 0.765 (0.514, 1.137) |
| Diarrhoea | True | 1.147 (0.752, 1.751) |
| Haemorrhoids | True | 1.004 (0.710, 1.420) |
| Diabetes T2 ever | True | 0.857 (0.520, 1.412) |
| Gallbladder calc ever | True | 0.678 (0.260, 1.769) |
| IBD ever | True | 0.718 (0.399, 1.292) |
| Aspirin | True | 0.677 (0.473, 0.967) |
| NSAIDs (non-aspirin) | True | 1.163 (0.758, 1.782) |
| Eligible for bowel cancer screening | True | 0.539 (0.398, 0.730) |
| Colonoscopy in last 10 years | True | 1.198 (0.805, 1.783) |
| Family history of bowel cancer | True | 0.934 (0.606, 1.439) |
| Family history of breast cancer | True | 0.877 (0.572, 1.345) |
| Family history of lung cancer | True | 0.890 (0.755, 1.048) |
| Multimorbidity Score (residual) | 0.50 | 1.523 (1.133, 2.046) |
| Inflammation | Abnormal | 3.901 (1.306, 11.655) |
| | Measured | 4.016 (2.879, 5.603) |
| Iron deficiency | Abnormal | 0.704 (0.298, 1.663) |
| | Measured | 1.326 (1.157, 1.520) |
| Polygenic Score | 0.09 | 1.977 (1.340, 2.918) |

***Supplementary Table 3a:**.Mean C-indices for all distinct coalitions of the predictor sets, as defined in Table S1, in the **study cohort**, estimated from the Cox PH model with bidirectional stepwise selection and 200 bootstrap samples.*[7]

| **Predictor sets** | **Mean C-index (95%CI)** |
|---|---|
| Core demographics | 0.670 (0.668,0.672) |
| Lifestyle | 0.566 (0.564,0.569) |
| Medical history | 0.600 (0.598,0.602) |
| PC blood tests | 0.634 (0.631,0.636) |
| Polygenic score | 0.595 (0.593,0.598) |
| Symptoms | 0.521 (0.519,0.522) |
| Core demographics, Lifestyle | 0.669 (0.667,0.671) |
| Core demographics, Medical history | 0.668 (0.666,0.670) |
| Core demographics, PC blood tests | 0.709 (0.706,0.711) |
| Core demographics, Polygenic score | 0.688 (0.686,0.690) |
| Core demographics, Symptoms | 0.673 (0.671,0.675) |
| Lifestyle, Medical history | 0.618 (0.615,0.620) |
| Lifestyle, PC blood tests | 0.655 (0.652,0.657) |
| Lifestyle, Polygenic score | 0.611 (0.609,0.614) |
| Lifestyle, Symptoms | 0.579 (0.576,0.581) |
| Medical history, PC blood tests | 0.670 (0.668,0.673) |
| Medical history, Polygenic score | 0.640 (0.638,0.642) |
| Medical history, Symptoms | 0.609 (0.607,0.612) |
| PC blood tests, Polygenic score | 0.674 (0.672,0.676) |
| PC blood tests, Symptoms | 0.638 (0.636,0.640) |
| Polygenic score, Symptoms | 0.604 (0.601,0.606) |
| Core demographics, Lifestyle, Medical history | 0.668 (0.666,0.670) |
| Core demographics, Lifestyle, PC blood tests | 0.710 (0.708,0.712) |
| Core demographics, Lifestyle, Polygenic score | 0.688 (0.685,0.690) |
| Core demographics, Lifestyle, Symptoms | 0.673 (0.671,0.675) |
| Core demographics, Medical history, PC blood tests | 0.710 (0.707,0.712) |
| Core demographics, Medical history, Polygenic score | 0.688 (0.685,0.690) |
| Core demographics, Medical history, Symptoms | 0.672 (0.670,0.674) |
| Core demographics, PC blood tests, Polygenic score | 0.725 (0.723,0.727) |
| Core demographics, PC blood tests, Symptoms | 0.710 (0.708,0.712) |
| Core demographics, Polygenic score, Symptoms | 0.691 (0.688,0.693) |
| Lifestyle, Medical history, PC blood tests | 0.679 (0.677,0.681) |
| Lifestyle, Medical history, Polygenic score | 0.646 (0.644,0.648) |
| Lifestyle, Medical history, Symptoms | 0.626 (0.623,0.628) |
| Lifestyle, PC blood tests, Polygenic score | 0.683 (0.681,0.685) |
| Lifestyle, PC blood tests, Symptoms | 0.658 (0.656,0.660) |
| Lifestyle, Polygenic score, Symptoms | 0.619 (0.617,0.621) |
| Medical history, PC blood tests, Polygenic score | 0.694 (0.692,0.696) |
| Medical history, PC blood tests, Symptoms | 0.674 (0.671,0.676) |
| Medical history, Polygenic score, Symptoms | 0.646 (0.643,0.648) |
| PC blood tests, Polygenic score, Symptoms | 0.675 (0.673,0.678) |

[7] Predictor set definitions: 'Core' variables include Age, Birth Year, Sex, Ethnicity, Body Mass Index, Smoking Status, and Townsend Deprivation Score. 'Lifestyle' variables cover Red Meat Consumption, Processed Meat Consumption, Fibre Consumption, Education (highest qualification achieved), and Alcohol consumption. 'Symptoms (recorded)' include Abdominal Bloating, Abdominal Pain, Rectal Bleeding, Change in Bowel Habit, Stomach Disorders, New-onset Fatigue, Recent Diagnosis of Diverticular Disease, Recent Diagnosis of Irritable Bowel Syndrome, New-onset Constipation, New-onset Diarrhoea, New-onset Haemorrhoids. 'Medical history (recorded)' encompasses Family History of Bowel Cancer, Family History of Breast Cancer, Family History of Lung Cancer, Eligibility for Bowel Cancer Screening, Colonoscopy in Last 10 Years, Previous Diagnosis of Inflammatory Bowel Disease, Previous Diagnosis of Type 2 Diabetes, Previous Diagnosis of Gallstones, Multi-morbidity Score, Regular Use of Non-aspirin NSAIDs, and Regular Use of Aspirin. 'Primary care blood tests (recorded)' involves tests for Iron Deficiency Anaemia and Inflammation, including both the measurement and abnormal result records. 'Polygenic score' includes Polygenic Score (PRS-CSx) and Principal Genetic Components (1-10).

| | |
|---|---|
| Core demographics, Lifestyle, Medical history, PC blood tests | 0.711 (0.709,0.713) |
| Core demographics, Lifestyle, Medical history, Polygenic score | 0.687 (0.685,0.689) |
| Core demographics, Lifestyle, Medical history, Symptoms | 0.673 (0.670,0.675) |
| Core demographics, Lifestyle, PC blood tests, Polygenic score | 0.726 (0.724,0.728) |
| Core demographics, Lifestyle, PC blood tests, Symptoms | 0.711 (0.709,0.713) |
| Core demographics, Lifestyle, Polygenic score, Symptoms | 0.690 (0.688,0.692) |
| Core demographics, Medical history, PC blood tests, Polygenic score | 0.726 (0.724,0.728) |
| Core demographics, Medical history, PC blood tests, Symptoms | 0.712 (0.710,0.714) |
| Core demographics, Medical history, Polygenic score, Symptoms | 0.691 (0.689,0.693) |
| Core demographics, PC blood tests, Polygenic score, Symptoms | 0.726 (0.724,0.728) |
| Lifestyle, Medical history, PC blood tests, Polygenic score | 0.701 (0.699,0.703) |
| Lifestyle, Medical history, PC blood tests, Symptoms | 0.682 (0.680,0.685) |
| Lifestyle, Medical history, Polygenic score, Symptoms | 0.652 (0.650,0.654) |
| Lifestyle, PC blood tests, Polygenic score, Symptoms | 0.684 (0.682,0.687) |
| Medical history, PC blood tests, Polygenic score, Symptoms | 0.696 (0.694,0.698) |
| Core demographics, Lifestyle, Medical history, PC blood tests, Polygenic score | 0.726 (0.724,0.729) |
| Core demographics, Lifestyle, Medical history, PC blood tests, Symptoms | 0.713 (0.711,0.716) |
| Core demographics, Lifestyle, Medical history, Polygenic score, Symptoms | 0.691 (0.689,0.693) |
| Core demographics, Lifestyle, PC blood tests, Polygenic score, Symptoms | 0.726 (0.724,0.729) |
| Core demographics, Medical history, PC blood tests, Polygenic score, Symptoms | 0.728 (0.725,0.730) |
| Lifestyle, Medical history, PC blood tests, Polygenic score, Symptoms | 0.703 (0.701,0.705) |
| Core demographics, Lifestyle, Medical history, PC blood tests, Polygenic score, Symptoms | 0.728 (0.726,0.731) |

***Supplementary Table 3b:*** Mean C-indices for all distinct coalitions of the predictor sets, as defined in Table S1, in the **symptomatic subcohort**, estimated from the Cox PH model with bidirectional stepwise selection and 200 bootstrap samples.[8]

| **Predictor types** | **Mean C-index (95%CI)** |
|---|---|
| Core demographics | 0.641 (0.636,0.646) |
| Lifestyle | 0.495 (0.489,0.500) |
| Medical history | 0.571 (0.565,0.577) |
| PC blood tests | 0.619 (0.614,0.625) |
| Polygenic score | 0.563 (0.558,0.569) |
| Symptoms | 0.564 (0.558,0.570) |
| Core demographics, Lifestyle | 0.631 (0.625,0.636) |
| Core demographics, Medical history | 0.630 (0.625,0.635) |
| Core demographics, PC blood tests | 0.681 (0.675,0.686) |
| Core demographics, Polygenic score | 0.640 (0.634,0.645) |
| Core demographics, Symptoms | 0.657 (0.651,0.663) |
| Lifestyle, Medical history | 0.561 (0.555,0.567) |
| Lifestyle, PC blood tests | 0.616 (0.609,0.622) |
| Lifestyle, Polygenic score | 0.546 (0.541,0.552) |
| Lifestyle, Symptoms | 0.551 (0.545,0.557) |
| Medical history, PC blood tests | 0.647 (0.642,0.653) |
| Medical history, Polygenic score | 0.588 (0.582,0.593) |
| Medical history, Symptoms | 0.611 (0.605,0.617) |
| PC blood tests, Polygenic score | 0.651 (0.645,0.657) |
| PC blood tests, Symptoms | 0.644 (0.638,0.650) |
| Polygenic score, Symptoms | 0.592 (0.586,0.598) |
| Core demographics, Lifestyle, Medical history | 0.621 (0.616,0.627) |
| Core demographics, Lifestyle, PC blood tests | 0.673 (0.667,0.679) |
| Core demographics, Lifestyle, Polygenic score | 0.632 (0.626,0.638) |
| Core demographics, Lifestyle, Symptoms | 0.648 (0.642,0.654) |
| Core demographics, Medical history, PC blood tests | 0.682 (0.677,0.688) |
| Core demographics, Medical history, Polygenic score | 0.629 (0.624,0.635) |
| Core demographics, Medical history, Symptoms | 0.649 (0.643,0.654) |
| Core demographics, PC blood tests, Polygenic score | 0.685 (0.679,0.691) |
| Core demographics, PC blood tests, Symptoms | 0.689 (0.683,0.695) |
| Core demographics, Polygenic score, Symptoms | 0.656 (0.650,0.662) |
| Lifestyle, Medical history, PC blood tests | 0.641 (0.635,0.647) |
| Lifestyle, Medical history, Polygenic score | 0.578 (0.572,0.583) |
| Lifestyle, Medical history, Symptoms | 0.600 (0.594,0.606) |
| Lifestyle, PC blood tests, Polygenic score | 0.639 (0.633,0.645) |
| Lifestyle, PC blood tests, Symptoms | 0.636 (0.630,0.642) |
| Lifestyle, Polygenic score, Symptoms | 0.579 (0.573,0.585) |
| Medical history, PC blood tests, Polygenic score | 0.663 (0.657,0.669) |
| Medical history, PC blood tests, Symptoms | 0.670 (0.663,0.676) |
| Medical history, Polygenic score, Symptoms | 0.619 (0.613,0.625) |
| PC blood tests, Polygenic score, Symptoms | 0.663 (0.657,0.669) |

[8] Predictor set definitions: 'Core' variables include Age, Birth Year, Sex, Ethnicity, Body Mass Index, Smoking Status, and Townsend Deprivation Score. 'Lifestyle' variables cover Red Meat Consumption, Processed Meat Consumption, Fibre Consumption, Education (highest qualification achieved), and Alcohol consumption. 'Symptoms (recorded)' include Abdominal Bloating, Abdominal Pain, Rectal Bleeding, Change in Bowel Habit, Stomach Disorders, New-onset Fatigue, Recent Diagnosis of Diverticular Disease, Recent Diagnosis of Irritable Bowel Syndrome, New-onset Constipation, New-onset Diarrhoea, New-onset Haemorrhoids. 'Medical history (recorded)' encompasses Family History of Bowel Cancer, Family History of Breast Cancer, Family History of Lung Cancer, Eligibility for Bowel Cancer Screening, Colonoscopy in Last 10 Years, Previous Diagnosis of Inflammatory Bowel Disease, Previous Diagnosis of Type 2 Diabetes, Previous Diagnosis of Gallstones, Multi-morbidity Score, Regular Use of Non-aspirin NSAIDs, and Regular Use of Aspirin. 'Primary care blood tests (recorded)' involves tests for Iron Deficiency Anaemia and Inflammation, including both the measurement and abnormal result records. 'Polygenic score' includes Polygenic Score (PRS-CSx) and Principal Genetic Components (1-10).

| | |
|---|---|
| Core demographics, Lifestyle, Medical history, PC blood tests | 0.675 (0.669,0.681) |
| Core demographics, Lifestyle, Medical history, Polygenic score | 0.623 (0.617,0.629) |
| Core demographics, Lifestyle, Medical history, Symptoms | 0.641 (0.635,0.647) |
| Core demographics, Lifestyle, PC blood tests, Polygenic score | 0.679 (0.673,0.685) |
| Core demographics, Lifestyle, PC blood tests, Symptoms | 0.683 (0.677,0.689) |
| Core demographics, Lifestyle, Polygenic score, Symptoms | 0.648 (0.641,0.654) |
| Core demographics, Medical history, PC blood tests, Polygenic score | 0.685 (0.679,0.691) |
| Core demographics, Medical history, PC blood tests, Symptoms | 0.692 (0.686,0.698) |
| Core demographics, Medical history, Polygenic score, Symptoms | 0.649 (0.643,0.654) |
| Core demographics, PC blood tests, Polygenic score, Symptoms | 0.694 (0.687,0.700) |
| Lifestyle, Medical history, PC blood tests, Polygenic score | 0.655 (0.649,0.661) |
| Lifestyle, Medical history, PC blood tests, Symptoms | 0.661 (0.655,0.667) |
| Lifestyle, Medical history, Polygenic score, Symptoms | 0.609 (0.603,0.615) |
| Lifestyle, PC blood tests, Polygenic score, Symptoms | 0.653 (0.646,0.659) |
| Medical history, PC blood tests, Polygenic score, Symptoms | 0.678 (0.672,0.685) |
| Core demographics, Lifestyle, Medical history, PC blood tests, Polygenic score | 0.679 (0.673,0.686) |
| Core demographics, Lifestyle, Medical history, PC blood tests, Symptoms | 0.686 (0.680,0.692) |
| Core demographics, Lifestyle, Medical history, Polygenic score, Symptoms | 0.642 (0.636,0.648) |
| Core demographics, Lifestyle, PC blood tests, Polygenic score, Symptoms | 0.687 (0.681,0.694) |
| Core demographics, Medical history, PC blood tests, Polygenic score, Symptoms | 0.695 (0.688,0.701) |
| Lifestyle, Medical history, PC blood tests, Polygenic score, Symptoms | 0.671 (0.664,0.677) |
| Core demographics, Lifestyle, Medical history, PC blood tests, Polygenic score, Symptoms | 0.689 (0.682,0.695) |

***Supplementary Table 4:*** Participant characteristics (excluding participants with Vision as their primary care data provider): Study cohort and symptomatic subcohort.[9]

| **Predictor** | **Level** | **N (%) for categorical; Median (IQR) for continuous** | |
|---|---|---|---|
| | | Study cohort ($N_{total}$=145764, $N_{cases}$=1209) | Symptomatic subcohort ($N_{total}$=38553, $N_{cases}$=206) |
| **Core** | | | |
| Age (baseline assessment) | years | 57.9 ( 50.3, 63.4) | 59.5 ( 52.0, 64.3) |
| Birth year | years | 1951.0 (1945.0,1958.0) | 1949.0 (1944.0,1957.0) |
| Body Mass Index | kg/m² | 26.8 ( 24.2, 29.9) | 27.1 ( 24.5, 30.5) |
| Ethnicity | White | 138762 (95.2%) | 36431 (94.5%) |
| | SE Asian | 2911 ( 2.0%) | 961 ( 2.5%) |
| | Black | 1466 ( 1.0%) | 457 ( 1.2%) |
| | Mixed | 765 ( 0.5%) | 204 ( 0.5%) |
| | Other | 1442 ( 1.0%) | 387 ( 1.0%) |
| | Missing | 418 ( 0.3%) | 113 ( 0.3%) |
| Sex (genetic) | Female | 76712 (52.6%) | 21655 (56.2%) |
| | Male | 69052 (47.4%) | 16898 (43.8%) |
| Smoking status | Never | 80574 (55.3%) | 20199 (52.4%) |
| | Current | 15286 (10.5%) | 4259 (11.0%) |
| | Former | 49479 (33.9%) | 13958 (36.2%) |
| | Missing | 425 ( 0.3%) | 137 ( 0.4%) |
| Townsend deprivation score | / | -2.2 ( -3.7, 0.4) | -2.1 ( -3.6, 0.7) |
| **Lifestyle** | | | |
| Alcohol | units/day | 1.7 ( 0.1, 3.6) | 1.4 ( 0.1, 3.3) |
| Education (highest qualification)[10] | Higher Education | 48439 (33.2%) | 10591 (27.5%) |
| | A-level | 16126 (11.1%) | 3870 (10.0%) |
| | GCSE | 30863 (21.2%) | 8440 (21.9%) |
| | CSE | 7503 ( 5.1%) | 2095 ( 5.4%) |
| | Vocational | 9895 ( 6.8%) | 2876 ( 7.5%) |
| | Professional | 7706 ( 5.3%) | 2062 ( 5.3%) |
| | None | 25232 (17.3%) | 8619 (22.4%) |
| Fibre consumption score | units/day | 13.6 ( 9.9, 17.5) | 13.6 ( 9.8, 17.7) |
| Processed meat consumption | servings/week | 1.0 ( 0.5, 3.0) | 1.0 ( 0.5, 3.0) |
| Red meat consumption | servings/week | 2.0 ( 1.5, 2.5) | 2.0 ( 1.5, 2.5) |
| **Symptoms*** | | | |
| Abdominal bloating | | 2600 ( 1.8%) | 1064 ( 2.8%) |
| Abdominal pain | | 19474 (13.4%) | 6354 (16.5%) |
| Change in Bowel habits | | 3098 ( 2.1%) | 1273 ( 3.3%) |
| Constipation | | 24527 (16.8%) | 24527 (63.6%) |
| Diarrhoea | | 22757 (15.6%) | 5891 (15.3%) |
| Diverticular | | 5933 ( 4.1%) | 5933 (15.4%) |
| Fatigue | | 11585 ( 7.9%) | 2184 ( 5.7%) |
| Haemorrhoids | | 14650 (10.1%) | 14650 (38.0%) |
| Irritable Bowel Syndrome | | 3670 ( 2.5%) | 1818 ( 4.7%) |
| Rectal bleed | | 5609 ( 3.8%) | 5609 (14.5%) |
| Stomach disorders | | 5294 ( 3.6%) | 1255 ( 3.3%) |
| **Medical history*** | | | |
| Aspirin | | 5578 ( 3.8%) | 2288 ( 5.9%) |
| Colonoscopy in last 10 years | | 34346 (23.6%) | 15955 (41.4%) |
| Diabetes T2 ever | | 11321 ( 7.8%) | 3343 ( 8.7%) |

[9] Unless otherwise specifed, sections with * indicates that the predictor level is by default “has at least one positive entry in the super-landmark dataframe”.

[10] Or equivalent qualifications.

| | | | |
|---|---|---|---|
| Eligible for bowel cancer screening | | 94050 (64.5%) | 22605 (58.6%) |
| Family history of bowel cancer | | 15801 (10.8%) | 4689 (12.2%) |
| Family history of breast cancer | | 14666 (10.1%) | 3906 (10.1%) |
| Family history of lung cancer | | 17947 (12.3%) | 5131 (13.3%) |
| Gallbladder calc ever | | 4891 ( 3.4%) | 1748 ( 4.5%) |
| Inflammatory bowel disease ever | | 2590 ( 1.8%) | 939 ( 2.4%) |
| Multimorbidity Score (residual) | Subject mean | -0.138 (-0.237, 0.136) | 0.050 (-0.192, 0.434) |
| NSAIDs (non-aspirin) | | 20963 (14.4%) | 8098 (21.0%) |
| **Primary care blood tests*** | | | |
| Inflammation | Abnormal | 34970 (24.0%) | 12881 (33.4%) |
| | Measured | 121963 (83.7%) | 35121 (91.1%) |
| Iron deficiency | Abnormal | 11735 ( 8.1%) | 4385 (11.4%) |
| | Measured | 119870 (82.2%) | 34895 (90.5%) |
| **Polygenic Score** | | | |
| Polygenic Score | / | 0.3 ( 0.2, 0.4) | 0.3 ( 0.2, 0.4) |

***Supplementary Table 5:*** *C-indices from 200 bootstrap samples for models derived using all predictor types (maximal coalition).*

| Cohort | PGS[11] | Mean C-index (95%CI) | #Individuals | #Cases | Total followup (person-years) | Mean followup per individual (SD; years) |
|---|---|---|---|---|---|---|
| Study | PRS-CSx | 0.728 (0.726,0.731) | 160507 | 1356 | 1374482.63 | 8.56 (2.52) |
| Symptomatic | PRS-CSx | 0.689 (0.682,0.695) | 42782 | 237 | 153073.95 | 3.58 (1.86) |
| Symptomatic (extended)[12] | PRS-CSx | 0.716 (0.711,0.721) | 70231 | 363 | 300173.68 | 4.27 (2.35) |
| Study | LDPred[13] | 0.784 (0.782,0.786) | 160507 | 1356 | 1374482.63 | 8.56 (2.52) |
| Symptomatic | LDPred | 0.740 (0.734,0.746) | 42782 | 237 | 153073.95 | 3.58 (1.86) |
| Study (sans Vision)[14] | PRS-CSx | 0.727 (0.725,0.730) | 145764 | 1209 | 1248803.95 | 8.57 (2.52) |
| Symptomatic (sans Vision) | PRS-CSx | 0.690 (0.683,0.696) | 38553 | 206 | 137251.38 | 3.56 (1.84) |

***Supplementary Table 6:*** *Hazard ratios of risk predictors selected using bidirectional (backwards/forwards) stepwise selection for all sensitivity analyses.[15]*

| Predictor | Predictor level or units[16] | HR (95% CI) |
|---|---|---|
| **Symptomatic subcohort (extended); PGS: PRS-CSx** | | |
| Age | 8.07 years | 1.254 (0.154, 10.246) |
| Age-squared | 971.06 years² | 0.704 (0.100, 4.940) |
| Birth year | 7.83 years | 0.535 (0.375, 0.763) |
| BMI | 5.04 kg/m² | 1.085 (0.972, 1.211) |
| Sex (genetic) | Male | 1.929 (1.523, 2.444) |
| Smoking status | Current | 1.199 (0.815, 1.762) |
| | * Missing | 0.000 (0.000, 0.000) |
| | Previous | 1.184 (0.936, 1.497) |
| Alcohol units daily | 2.90 units/day | 1.082 (1.016, 1.152) |
| Education | A-level | 0.628 (0.391, 1.008) |
| | GCSE | 0.861 (0.631, 1.174) |
| | CSE | 0.844 (0.489, 1.455) |
| | Vocational | 0.713 (0.454, 1.120) |
| | Professional | 0.739 (0.441, 1.241) |
| | None | 0.658 (0.478, 0.904) |
| Change in Bowel habits | True | 1.430 (0.849, 2.407) |
| Fatigue | True | 0.599 (0.338, 1.064) |
| Rectal bleed | True | 2.758 (2.041, 3.727) |
| Constipation | True | 1.188 (0.948, 1.489) |
| Diarrhoea | True | 0.844 (0.637, 1.117) |

---

[11] Polygenic Score (PGS)

[12] The extended symptomatic subcohort consists of participants with any CRC symptom (listed in Table S1), except fatigue (not considered to be sufficiently specific).

[13] The study and symptomatic subcohorts using an alternative polygenic score (PGS), namely the LDPred-derived PGS by Thomas et al. (13). This PGS was developed in a European-ancestry only cohort that includes some UKB cohort members.

[14] The study and symptomatic subcohorts excluding participants with English Vision primary care data (as primary care data for those who died pre-2017 are mostly unavailable) (14).

[15] * indicates no CRC cases with that risk predictor level.

[16] Continuous variables are standardized, with their standard deviations presented here. Hazard Ratios (HRs) are interpreted per standard deviation increase.

| | | |
|---|---|---|
| Haemorrhoids | True | 1.482 (1.131, 1.942) |
| Gallbladder calc ever | True | 1.387 (0.886, 2.170) |
| NSAIDs (non-aspirin) | True | 0.709 (0.531, 0.947) |
| Eligible for bowel cancer screening | True | 1.261 (0.908, 1.751) |
| Colonoscopy in last 10 years | True | 0.472 (0.364, 0.612) |
| Multimorbidity Score (residual) | 0.49 | 0.880 (0.780, 0.992) |
| Inflammation | Abnormal | 1.851 (1.462, 2.344) |
| | Measured | 1.779 (0.969, 3.268) |
| Iron deficiency | Abnormal | 3.738 (2.852, 4.901) |
| Polygenic Score | 0.09 | 1.306 (1.172, 1.456) |
| **Study cohort; PGS: LDPred** | | |
| Age | 8.14 years | 5.617 (2.025, 15.582) |
| Age-squared | 975.00 years² | 0.228 (0.088, 0.588) |
| Birth year | 7.90 years | 0.643 (0.545, 0.760) |
| BMI | 4.75 kg/m² | 1.063 (1.005, 1.125) |
| Sex (genetic) | Male | 1.595 (1.414, 1.799) |
| Smoking status | Current | 1.166 (0.964, 1.410) |
| | Missing | 0.499 (0.153, 1.630) |
| | Previous | 1.148 (1.020, 1.292) |
| Alcohol units daily | 2.89 units/day | 1.123 (1.086, 1.161) |
| Fibre consumption | 6.37 units/day | 0.970 (0.916, 1.027) |
| Processed meat consumption | 1.39 servings/week | 1.037 (0.984, 1.094) |
| Change in Bowel habits | True | 1.409 (0.844, 2.352) |
| Fatigue | True | 0.732 (0.502, 1.068) |
| Rectal bleed | True | 2.690 (1.992, 3.633) |
| Constipation | True | 1.194 (0.979, 1.457) |
| Haemorrhoids | True | 1.494 (1.154, 1.935) |
| Diabetes T2 ever | True | 1.120 (0.911, 1.376) |
| Aspirin | True | 0.828 (0.632, 1.085) |
| NSAIDs (non-aspirin) | True | 0.747 (0.632, 0.882) |
| Eligible for bowel cancer screening | True | 0.904 (0.776, 1.053) |
| Colonoscopy in last 10 years | True | 0.581 (0.493, 0.683) |
| Family history of bowel cancer | True | 1.094 (0.932, 1.284) |
| Multimorbidity Score (residual) | 0.39 | 0.930 (0.876, 0.987) |
| Inflammation | Abnormal | 1.717 (1.518, 1.941) |
| | Measured | 1.399 (1.129, 1.735) |
| Iron deficiency | Abnormal | 3.855 (3.333, 4.458) |
| Polygenic Score | 0.24 | 2.250 (2.127, 2.380) |
| **Symptomatic subcohort; PGS: LDPred** | | |
| Age | 8.02 years | 0.919 (0.070, 11.975) |
| Age-squared | 968.90 years² | 1.265 (0.112, 14.304) |
| Birth year | 7.78 years | 0.621 (0.415, 0.931) |
| BMI | 5.03 kg/m² | 1.165 (1.015, 1.336) |
| Sex (genetic) | Male | 2.033 (1.535, 2.693) |
| Education | A-level | 0.706 (0.402, 1.238) |
| | GCSE | 0.895 (0.611, 1.312) |
| | CSE | 1.087 (0.593, 1.991) |
| | Vocational | 0.420 (0.220, 0.803) |
| | Professional | 0.866 (0.476, 1.574) |
| | None | 0.545 (0.364, 0.815) |
| Abdominal bloating | True | 0.190 (0.027, 1.346) |
| Diverticular | True | 0.626 (0.363, 1.078) |
| Fatigue | True | 0.563 (0.265, 1.196) |
| Rectal bleed | True | 1.932 (1.327, 2.810) |

| | | |
|---|---|---|
| Constipation | True | 0.771 (0.563, 1.057) |
| Aspirin | True | 0.711 (0.398, 1.271) |
| NSAIDs (non-aspirin) | True | 0.709 (0.497, 1.012) |
| Colonoscopy in last 10 years | True | 0.532 (0.395, 0.719) |
| Multimorbidity Score (residual) | 0.5 | 0.884 (0.757, 1.032) |
| Inflammation | Abnormal | 1.501 (1.115, 2.022) |
| | Measured | 2.893 (1.216, 6.881) |
| Iron deficiency | Abnormal | 3.890 (2.800, 5.404) |
| Polygenic Score | 0.24 | 2.104 (1.822, 2.431) |
| **Study cohort (sans Vision); PGS: PRS-CSx** | | |
| Age | 8.14 years | 6.225 (2.153, 17.996) |
| Age-squared | 974.94 years² | 0.174 (0.064, 0.469) |
| Birth year | 7.90 years | 0.576 (0.487, 0.682) |
| BMI | 4.75 kg/m² | 1.067 (1.006, 1.131) |
| Sex (genetic) | Male | 1.624 (1.429, 1.845) |
| Smoking status | Current | 1.172 (0.959, 1.434) |
| | Missing | 0.575 (0.173, 1.907) |
| | Previous | 1.136 (1.002, 1.287) |
| Alcohol units daily | 2.90 units/day | 1.116 (1.081, 1.152) |
| Fibre consumption | 6.36 units/day | 0.968 (0.910, 1.029) |
| Processed meat consumption | 1.39 servings/week | 1.035 (0.978, 1.095) |
| Fatigue | True | 0.615 (0.389, 0.973) |
| Rectal bleed | True | 2.829 (2.046, 3.910) |
| Constipation | True | 1.152 (0.932, 1.426) |
| Haemorrhoids | True | 1.541 (1.171, 2.027) |
| Diabetes T2 ever | True | 1.216 (0.983, 1.505) |
| Aspirin | True | 0.767 (0.547, 1.076) |
| NSAIDs (non-aspirin) | True | 0.733 (0.613, 0.877) |
| Colonoscopy in last 10 years | True | 0.599 (0.505, 0.711) |
| Family history of bowel cancer | True | 1.154 (0.973, 1.369) |
| Multimorbidity Score (residual) | 0.39 | 0.935 (0.878, 0.996) |
| Inflammation | Abnormal | 1.790 (1.571, 2.040) |
| | Measured | 1.380 (1.108, 1.718) |
| Iron deficiency | Abnormal | 3.888 (3.323, 4.549) |
| Polygenic Score | 0.09 | 1.392 (1.310, 1.478) |
| **Symptomatic subcohort (sans Vision); PGS: PRS-CSx** | | |
| Age | 8.03 years | 3.153 (0.206, 48.383) |
| Age-squared | 969.09 years² | 0.339 (0.026, 4.469) |
| Birth year | 7.79 years | 0.561 (0.369, 0.852) |
| BMI | 5.04 kg/m² | 1.154 (0.999, 1.332) |
| Sex (genetic) | Male | 2.150 (1.586, 2.913) |
| Townsend | 3.15 | 1.098 (0.938, 1.285) |
| Education | A-level | 0.610 (0.326, 1.143) |
| | GCSE | 0.841 (0.560, 1.262) |
| | CSE | 0.731 (0.359, 1.490) |
| | Vocational | 0.410 (0.208, 0.811) |
| | Professional | 0.769 (0.403, 1.467) |
| | None | 0.497 (0.322, 0.767) |
| Abdominal bloating | * True | 0.000 (0.000, 0.000) |
| Diverticular | True | 0.628 (0.351, 1.125) |
| Fatigue | True | 0.563 (0.250, 1.269) |
| Rectal bleed | True | 2.039 (1.368, 3.041) |
| Constipation | True | 0.711 (0.508, 0.994) |
| IBD ever | True | 0.292 (0.073, 1.167) |

| | | |
|---|---|---|
| Aspirin | True | 0.596 (0.292, 1.217) |
| NSAIDs (non-aspirin) | True | 0.697 (0.479, 1.014) |
| Colonoscopy in last 10 years | True | 0.534 (0.387, 0.736) |
| Inflammation | Abnormal | 1.531 (1.120, 2.092) |
| | Measured | 2.674 (1.115, 6.414) |
| Iron deficiency | Abnormal | 3.777 (2.653, 5.376) |
| Polygenic Score | 0.09 | 1.324 (1.143, 1.534) |

***Supplementary Table 7:*** *Participant characteristics: Full UKBiobank, study cohort and symptomatic subcohort.*[17]

| Predictor | Level | N (%) for categorical; Median (IQR) for continuous | | |
|---|---|---|---|---|
| | | Full UKBiobank cohort (N=502413) | Study cohort (N=160507) | Symptomatic subcohort[18] (N=42782) |
| **Core** | | | | |
| Age (baseline assessment) | Years | 58.3 ( 50.6, 63.7) | 57.9 ( 50.3, 63.4) | 59.5 ( 52.0, 64.3) |
| Birth year | Years | 1950.0 (1945.0,1958.0) | 26.8 ( 24.2, 29.9) | 27.1 ( 24.5, 30.5) |
| Body Mass Index | kg/m² | 26.7 ( 24.2, 29.8) | 1951 (1945,1958) | 1949 (1944,1957) |
| Ethnicity | White | 472616 (94.1%) | 152514 (95.0%) | 40342 (94.3%) |
| | SE Asian | 9879 ( 2.0%) | 3258 ( 2.0%) | 1077 ( 2.5%) |
| | Black | 8058 ( 1.6%) | 1732 ( 1.1%) | 543 ( 1.3%) |
| | Mixed | 2954 ( 0.6%) | 880 ( 0.5%) | 231 ( 0.5%) |
| | Other | 6130 ( 1.2%) | 1658 ( 1.0%) | 464 ( 1.1%) |
| | Missing | 2776 ( 0.6%) | 465 ( 0.3%) | 125 ( 0.3%) |
| Sex (genetic) | Female | 264745 (54.2%) | 84624 (52.7%) | 24045 (56.2%) |
| | Male | 223430 (45.8%) | 75883 (47.3%) | 18737 (43.8%) |
| Smoking status | Never | 273478 (54.4%) | 88699 (55.3%) | 22421 (52.4%) |
| | Current | 52962 (10.5%) | 16754 (10.4%) | 4705 (11.0%) |
| | Previous | 173025 (34.4%) | 54579 (34.0%) | 15499 (36.2%) |
| | Missing | 2948 ( 0.6%) | 475 ( 0.3%) | 157 ( 0.4%) |
| Townsend deprivation score | / | -2.1 ( -3.6, 0.5) | -2.2 ( -3.7, 0.4) | -2.1 ( -3.6, 0.7) |
| **Lifestyle** | | | | |
| Alcohol consumption | Units/day | 1.6 ( 0.1, 3.5) | 1.7 ( 0.1, 3.6) | 1.4 ( 0.1, 3.3) |
| Education | Higher Education | 161130 (32.7%) | 53615 (33.4%) | 11820 (27.6%) |
| (highest qualification)[19] | A-level | 55311 (11.2%) | 17839 (11.1%) | 4320 (10.1%) |
| | GCSE | 105176 (21.4%) | 33958 (21.2%) | 9345 (21.8%) |
| | CSE | 26885 ( 5.5%) | 8313 ( 5.2%) | 2351 ( 5.5%) |
| | Vocational | 32724 ( 6.6%) | 10795 ( 6.7%) | 3152 ( 7.4%) |
| | Professional | 25799 ( 5.2%) | 8508 ( 5.3%) | 2294 ( 5.4%) |
| | None | 85259 (17.3%) | 27479 (17.1%) | 9500 (22.2%) |
| Fibre consumption score | Units/day | 13.6 ( 9.9, 17.6) | 13.6 ( 9.9, 17.6) | 13.6 ( 9.8, 17.7) |
| Processed meat consumption | Servings/week | 1.0 ( 0.5, 3.0) | 1.0 ( 0.5, 3.0) | 1.0 ( 0.5, 3.0) |
| Red meat consumption | Servings/week | 2.0 ( 1.5, 2.5) | 2.0 ( 1.5, 2.5) | 2.0 ( 1.5, 2.5) |
| **Medical history*** | | | | |
| Family history of bowel cancer | True | 54619 (10.9%) | 17402 (10.8%) | 5225 (12.2%) |
| Family history of breast cancer | True | 52495 (10.4%) | 16197 (10.1%) | 4357 (10.2%) |
| Family history of lung cancer | True | 62205 (12.4%) | 19701 (12.3%) | 5657 (13.2%) |

17 Percentages for full UKBiobank cohort exclude NAs unless explicitly labeled as "Missing".

18 The "symptomatic" subcohort consists of individuals with any of four selected symptoms during their follow-up: new-onset haemorrhoids, new-onset constipation, recent rectal bleeding and recent diverticular disease (see Table S1, Fig S3, Supplementary Methods 2). While these four symptoms formed the eligibility criteria for inclusion in this subcohort, some patients also presented with additional symptoms.

19 Or equivalent qualifications.

***Supplementary Table 8:*** *Discriminative contribution of predictors using Shapley values (C-index > 0.5) from 200 bootstrap samples.*

| Cohort | Predictor type | Mean Discriminative Contribution (95% CI) |
|---|---|---|
| Study | Core demographics | 33.3% (25.1%,41.9%) |
| | Lifestyle | 5.5% (0.3%,10.6%) |
| | Medical history | 11.1% (3.8%,17.4%) |
| | PC blood tests | 31.6% (18.6%,42.9%) |
| | Polygenic score | 16.0% (7.7%,25.5%) |
| | Symptoms | 2.5% (-2.0%,7.0%) |
| Symptomatic | Core demographics | 34.0% (8.7%,74.9%) |
| | Lifestyle | -4.9% (-31.9%,13.4%) |
| | Medical history | 8.6% (-25.3%,37.4%) |
| | PC blood tests | 40.9% (16.3%,78.2%) |
| | Polygenic score | 8.3% (-21.4%,34.8%) |
| | Symptoms | 13.2% (-18.5%,41.2%) |
| Symptomatic (extended) | Core demographics | 31.6% (10.8%,50.7%) |
| | Lifestyle | -3.1% (-26.7%,8.0%) |
| | Medical history | 12.1% (-13.8%,28.9%) |
| | PC blood tests | 38.0% (16.0%,61.7%) |
| | Polygenic score | 9.9% (-12.6%,29.5%) |
| | Symptoms | 11.6% (-9.7%,32.6%) |
| Study (sans Vision) | Core demographics | 34.2% (25.8%,44.2%) |
| | Lifestyle | 5.2% (-0.1%,11.0%) |
| | Medical history | 10.7% (2.6%,18.4%) |
| | PC blood tests | 32.0% (20.8%,42.1%) |
| | Polygenic score | 15.5% (7.8%,25.3%) |
| | Symptoms | 2.4% (-3.1%,7.1%) |
| Symptomatic (sans Vision) | Core demographics | 38.0% (8.5%,82.4%) |
| | Lifestyle | -6.5% (-43.0%,15.7%) |
| | Medical history | 7.7% (-38.3%,43.2%) |
| | PC blood tests | 41.2% (9.9%,77.5%) |
| | Polygenic score | 4.1% (-33.7%,35.2%) |
| | Symptoms | 15.4% (-19.4%,49.7%) |
| Study (LDpred) | Core demographics | 21.6% (15.9%,27.2%) |
| | Lifestyle | 3.8% (0.2%,7.1%) |
| | Medical history | 7.6% (2.8%,12.0%) |
| | PC blood tests | 19.6% (12.4%,27.4%) |
| | Polygenic score | 45.7% (35.1%,55.7%) |
| | Symptoms | 1.7% (-1.2%,4.8%) |
| Symptomatic (LDpred) | Core demographics | 21.5% (-0.1%,42.2%) |
| | Lifestyle | -3.9% (-24.8%,8.7%) |
| | Medical history | 6.2% (-14.1%,25.1%) |
| | PC blood tests | 25.9% (8.4%,45.4%) |
| | Polygenic score | 43.0% (11.1%,68.7%) |
| | Symptoms | 7.4% (-14.7%,24.7%) |

***Supplementary Table 9:*** *Using only age or age² as the age variable to investigate the wide CIs in Figure 2a. Hazard ratios of risk predictors selected using Cox regression with three different variable selection approaches (no selection, bidirectional selection , and lasso) in the symptomatic subcohort.*

| **Predictor** | **Predictor level or units[20]** | **HR (95% CI)** | |
|---|---|---|---|
| | | **Only age** | **Only age²** |
| Age-variable | 8.02 years (age); 968.90 years² (age²) | 1.163 (0.785, 1.722) | 1.154 (0.795, 1.675) |
| Birth year | 7.78 years | 0.626 (0.419, 0.934) | 0.623 (0.420, 0.924) |
| BMI | 5.03 kg/m² | 1.159 (1.012, 1.328) | 1.160 (1.012, 1.328) |
| Sex (genetic) | Male (genetic) | 2.032 (1.535, 2.688) | 2.031 (1.535, 2.688) |
| Education | A-level | 0.701 (0.398, 1.235) | 0.700 (0.398, 1.234) |
| | GCSE | 0.893 (0.612, 1.304) | 0.893 (0.612, 1.303) |
| | CSE | 1.118 (0.616, 2.030) | 1.117 (0.616, 2.028) |
| | Vocational | 0.425 (0.222, 0.813) | 0.425 (0.222, 0.813) |
| | Professional | 0.855 (0.470, 1.554) | 0.854 (0.470, 1.553) |
| | None | 0.552 (0.371, 0.822) | 0.552 (0.371, 0.821) |
| Abdominal bloating | True | 0.186 (0.026, 1.309) | 0.186 (0.026, 1.312) |
| Diverticular | True | 0.624 (0.361, 1.079) | 0.624 (0.361, 1.079) |
| Fatigue | True | 0.570 (0.269, 1.209) | 0.570 (0.269, 1.209) |
| Rectal bleed | True | 1.961 (1.350, 2.850) | 1.962 (1.350, 2.852) |
| Constipation | True | 0.762 (0.556, 1.046) | 0.762 (0.555, 1.046) |
| Aspirin | True | 0.716 (0.398, 1.285) | 0.715 (0.398, 1.284) |
| NSAIDs (non-aspirin) | True | 0.690 (0.483, 0.984) | 0.690 (0.484, 0.984) |
| Colonoscopy in last 10 years | True | 0.534 (0.396, 0.721) | 0.534 (0.396, 0.721) |
| Multimorbidity Score (residual) | 0.50 | 0.885 (0.759, 1.032) | 0.885 (0.759, 1.032) |
| Inflammation | Abnormal | 1.511 (1.125, 2.030) | 1.512 (1.125, 2.031) |
| | Measured | 2.855 (1.201, 6.788) | 2.858 (1.202, 6.796) |
| Iron deficiency | Abnormal | 3.989 (2.874, 5.537) | 3.985 (2.871, 5.534) |
| Polygenic Score | 0.09 | 1.330 (1.160, 1.526) | 1.331 (1.160, 1.527) |

[20] Continuous variables are standardised, with their standard deviations presented here. Hazard Ratios (HRs) are interpreted per standard deviation increase.

Supplementary Figures - Relating to main results

***Supplementary Figure 1:*** *Study cohort selection flowchart*

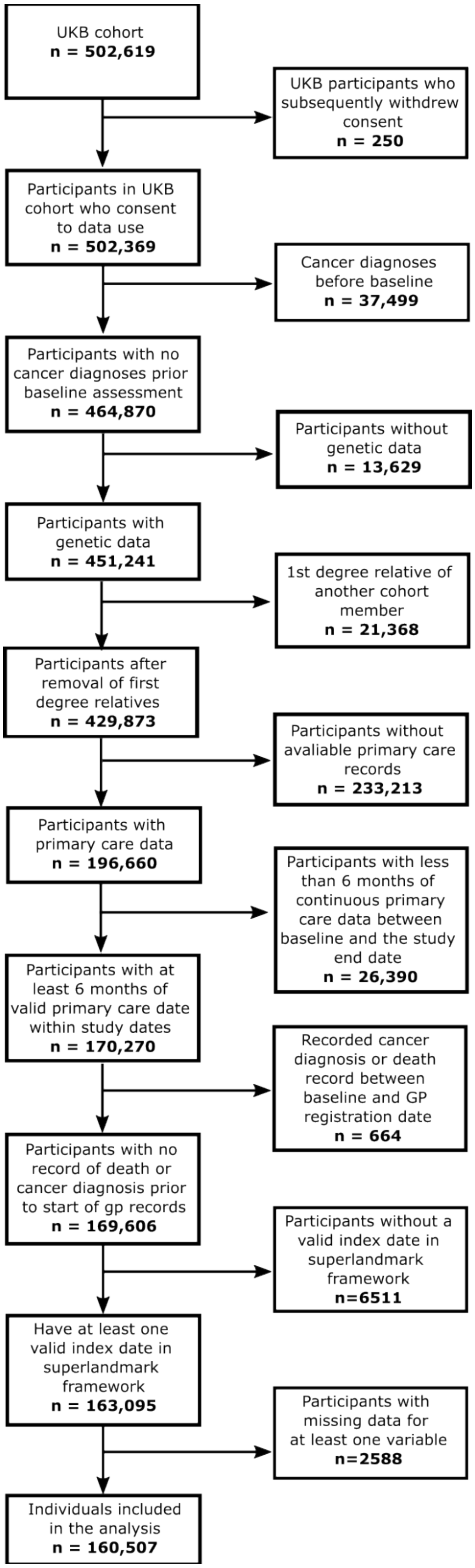

[illegible]

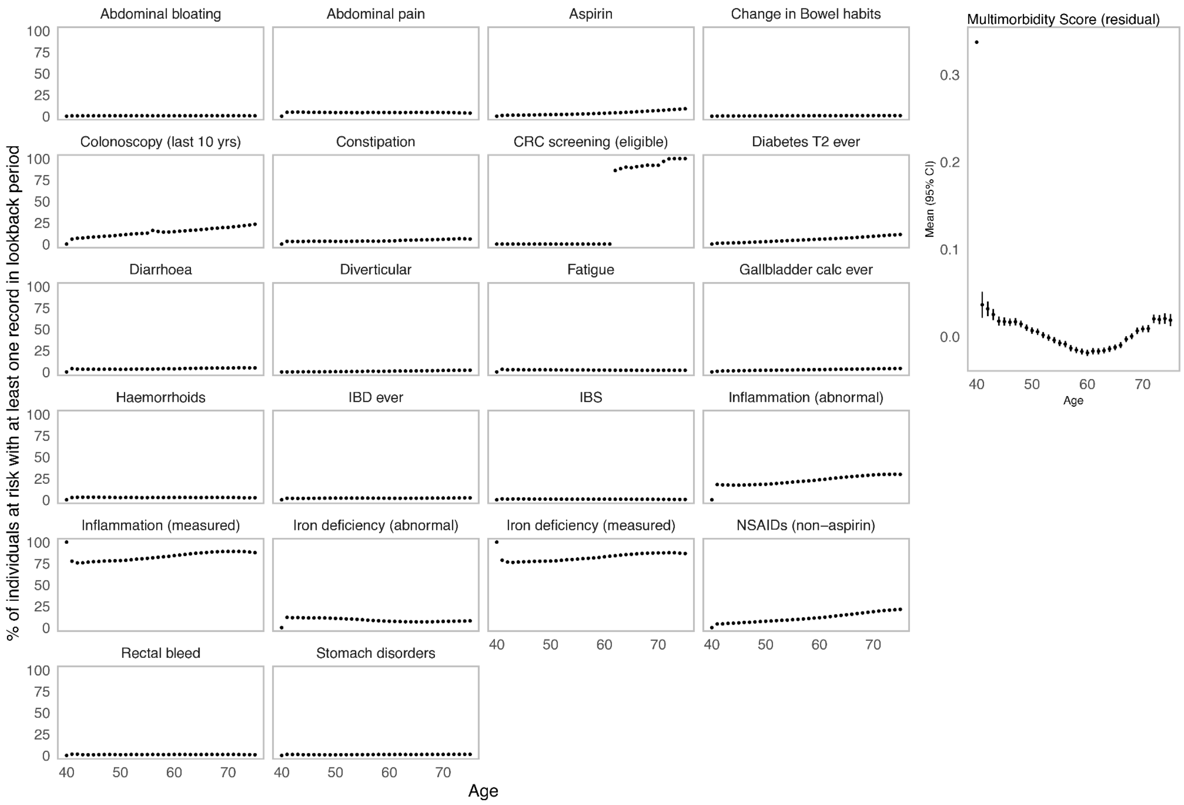

Abdominal bloating
Abdominal pain
Aspirin
Change in Bowel habits
Colonoscopy (last 10 yrs)
Constipation
CRC screening (eligible)
Diabetes T2 ever
Diarrhoea
Diverticular
Fatigue
Gallbladder calc ever
Haemorrhoids
IBD ever
IBS
Inflammation (abnormal)
Inflammation (measured)
Iron deficiency (abnormal)
Iron deficiency (measured)
NSAIDs (non–aspirin)
Rectal bleed
Stomach disorders
% of individuals at risk with at least one record in lookback period
100
75
50
25
0
40
50
60
70
Age
Multimorbidity Score (residual)
Mean (95% CI)
0.3
0.2
0.1
0.0

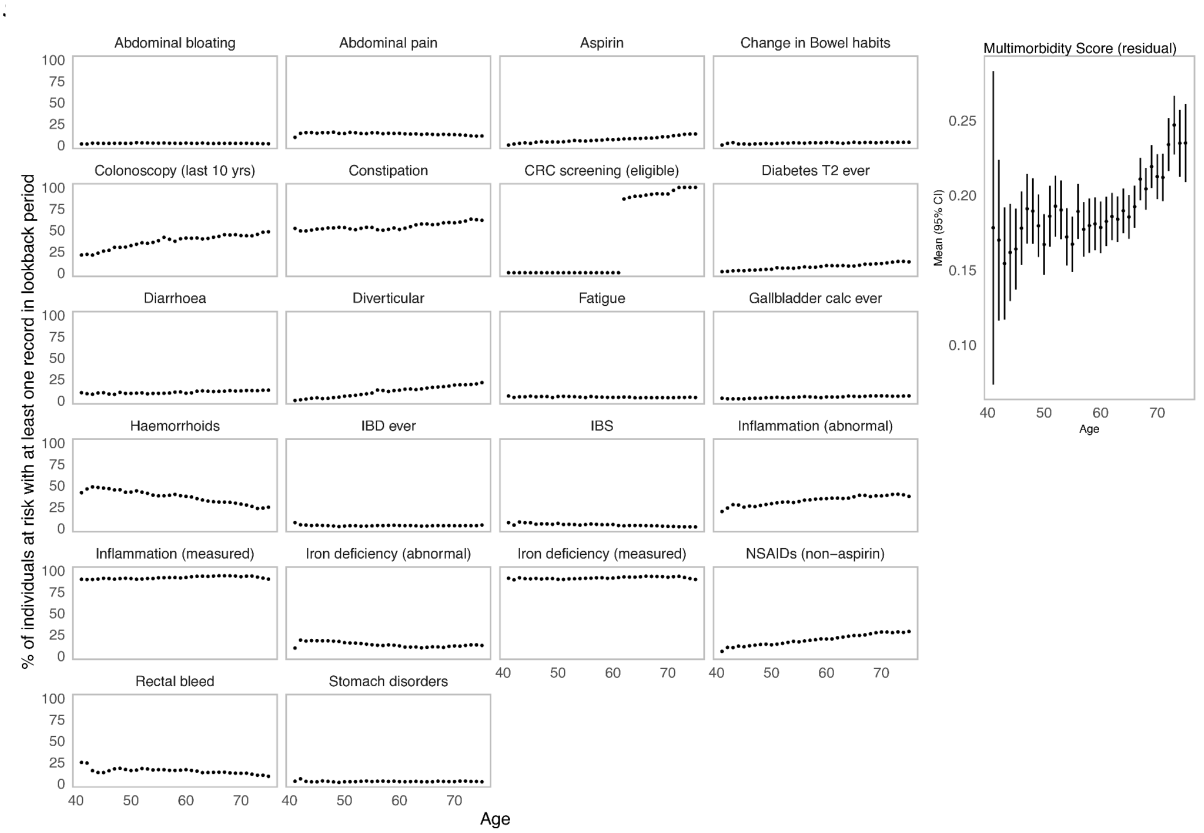

Abdominal bloating
Abdominal pain
Aspirin
Change in Bowel habits
Colonoscopy (last 10 yrs)
Constipation
CRC screening (eligible)
Diabetes T2 ever
Diarrhoea
Diverticular
Fatigue
Gallbladder calc ever
Haemorrhoids
IBD ever
IBS
Inflammation (abnormal)
Inflammation (measured)
Iron deficiency (abnormal)
Iron deficiency (measured)
NSAIDs (non−aspirin)
Rectal bleed
Stomach disorders
% of individuals at risk with at least one record in lookback period
Age
Multimorbidity Score (residual)
Mean (95% CI)
Age

***Supplementary Figure 3:*** *Age and sex-adjusted hazard ratios for symptoms selected by Cox PH bidirectional stepwise selection from the "symptoms" predictor set (Table S1), defining our "symptomatic" subcohort*

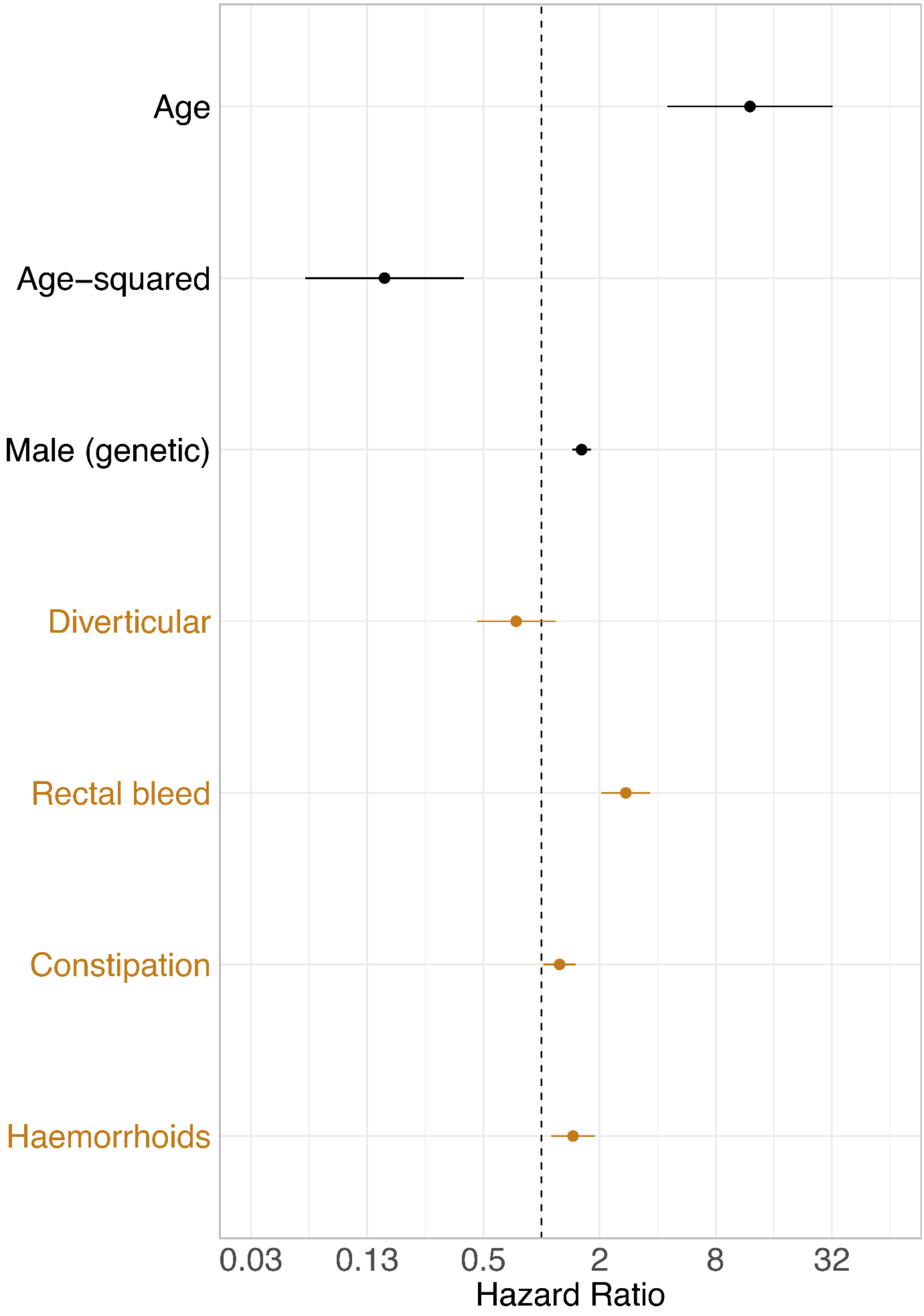

***Supplementary Figure 4:*** *C-indices using 3 different predictor selection approaches for CRC risk prediction in the (a) study cohort and (b) symptomatic subcohort, respectively.*

(a) Study cohort

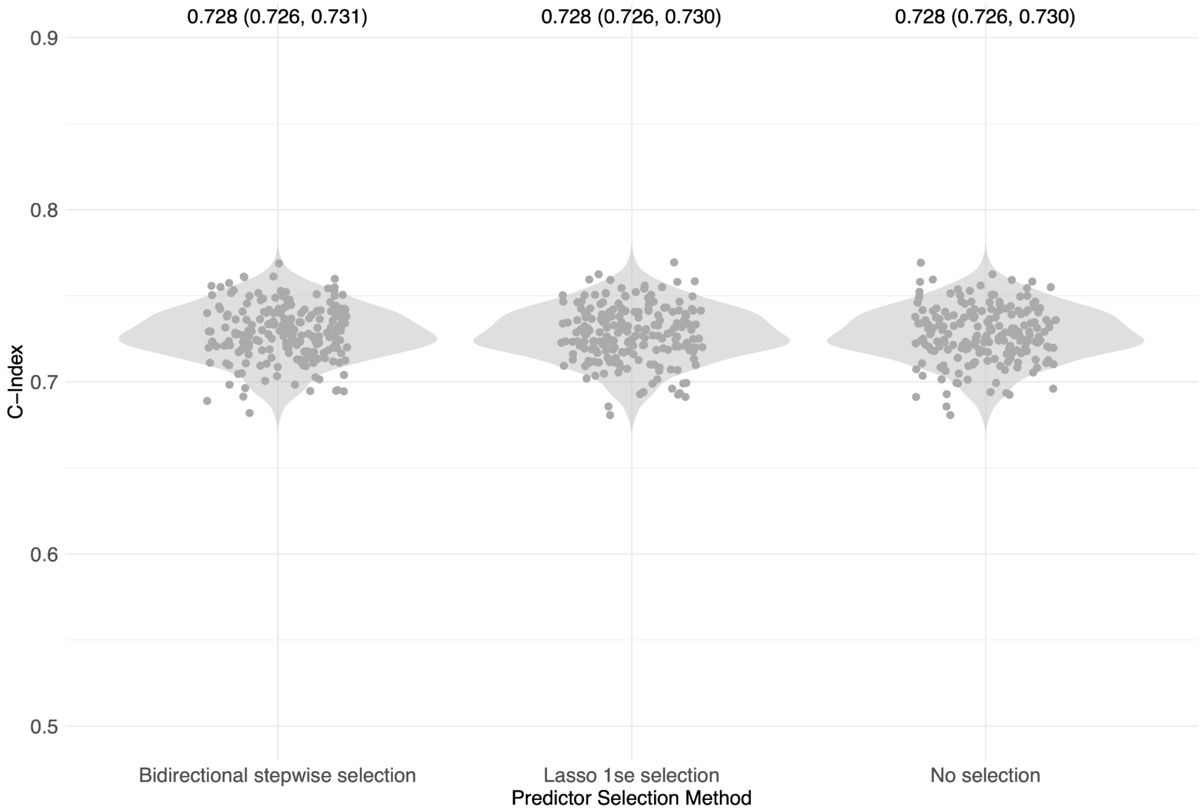

(b) Symptomatic subcohort

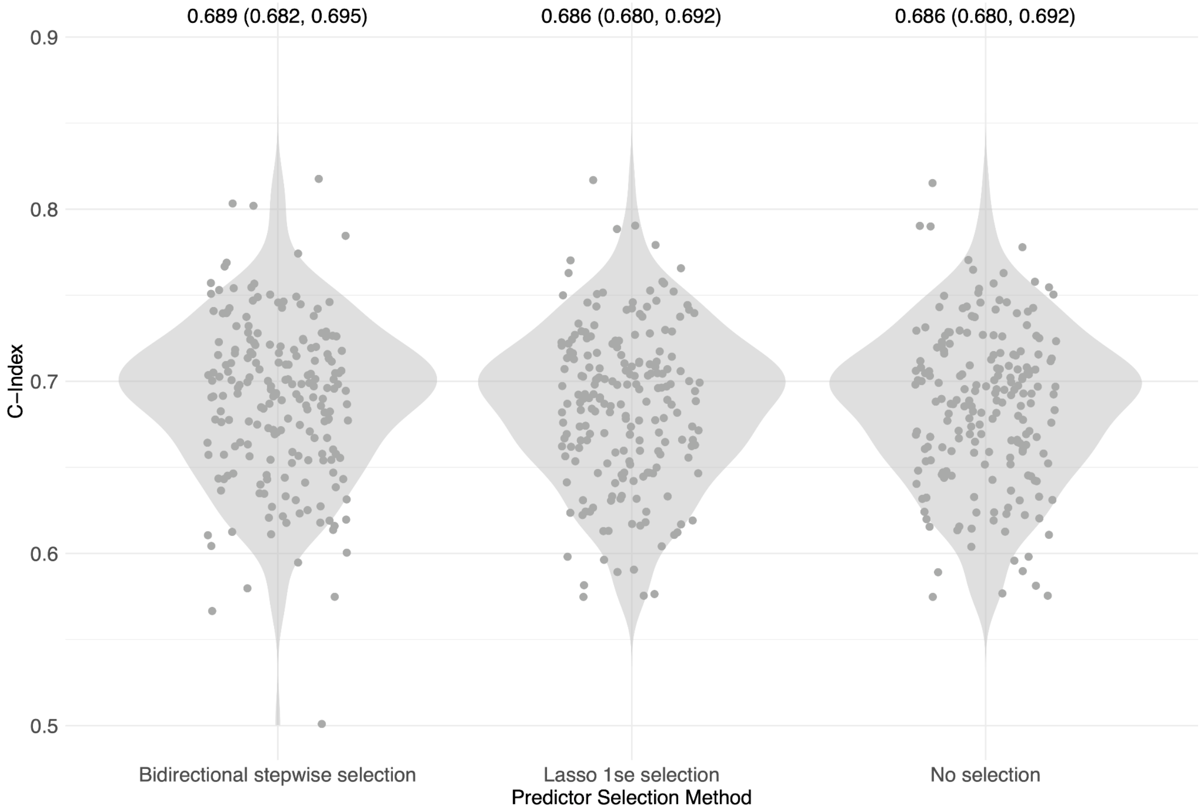

***Supplementary Figure 5:*** *Venn diagram to show which risk predictors were selected from each predictor selection approach for the (a) study cohort and (b) symptomatic subcohort, respectively.*

(a) Study cohort

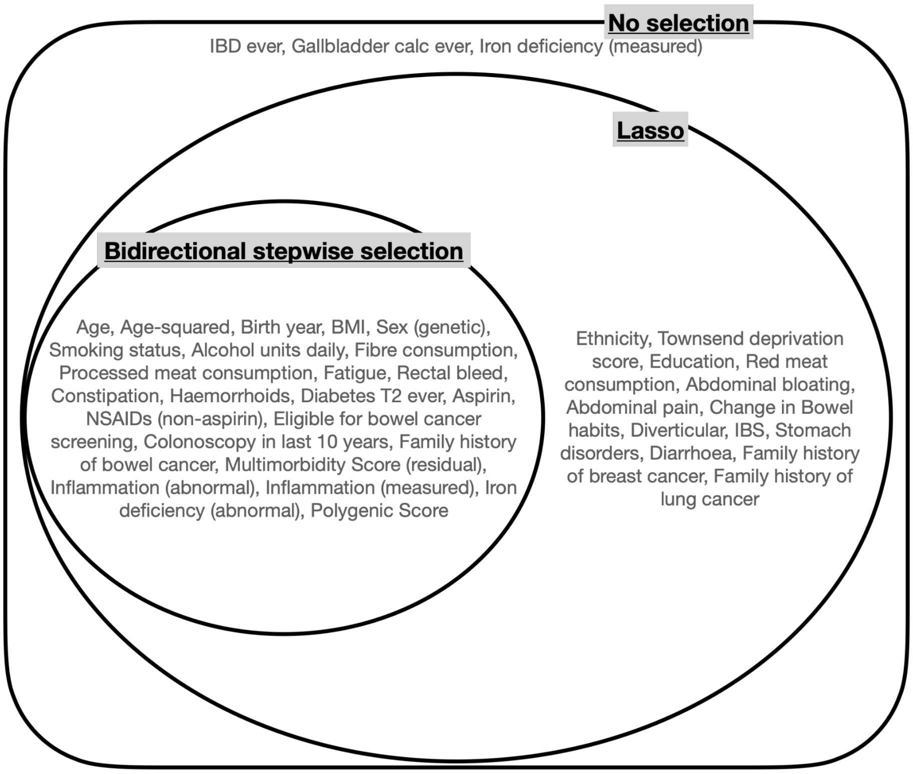

(b) Symptomatic subcohort

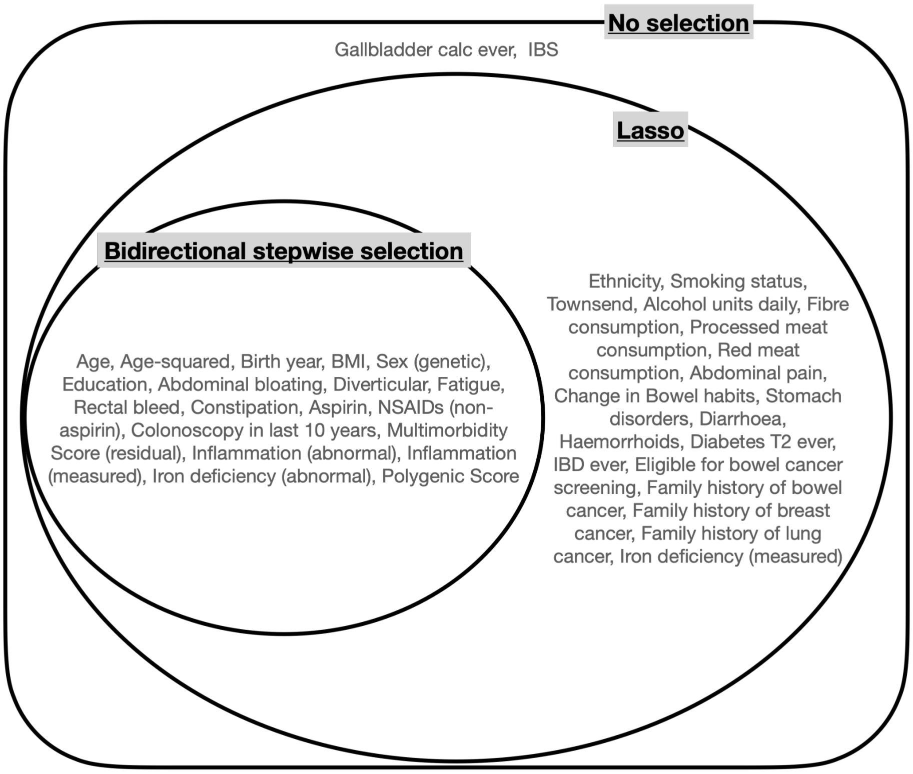

***Supplementary Figure 6a:  Figure 6a-a*** *Hazard ratios from bidirectional stepwise Cox regression for the* ***study cohort****, using* ***LDPred-generated polygenic scores; Figure 6a-b*** *The inclusion-order-agnostic discriminative contribution (C-index > 0.5) of each predictor set evaluated using Shapley values;* ***Figure 6a-c*** *C-indices from 200 bootstrap samples for each coalition of predictor sets. Colour-coding indicates the predictor set in all figures.*

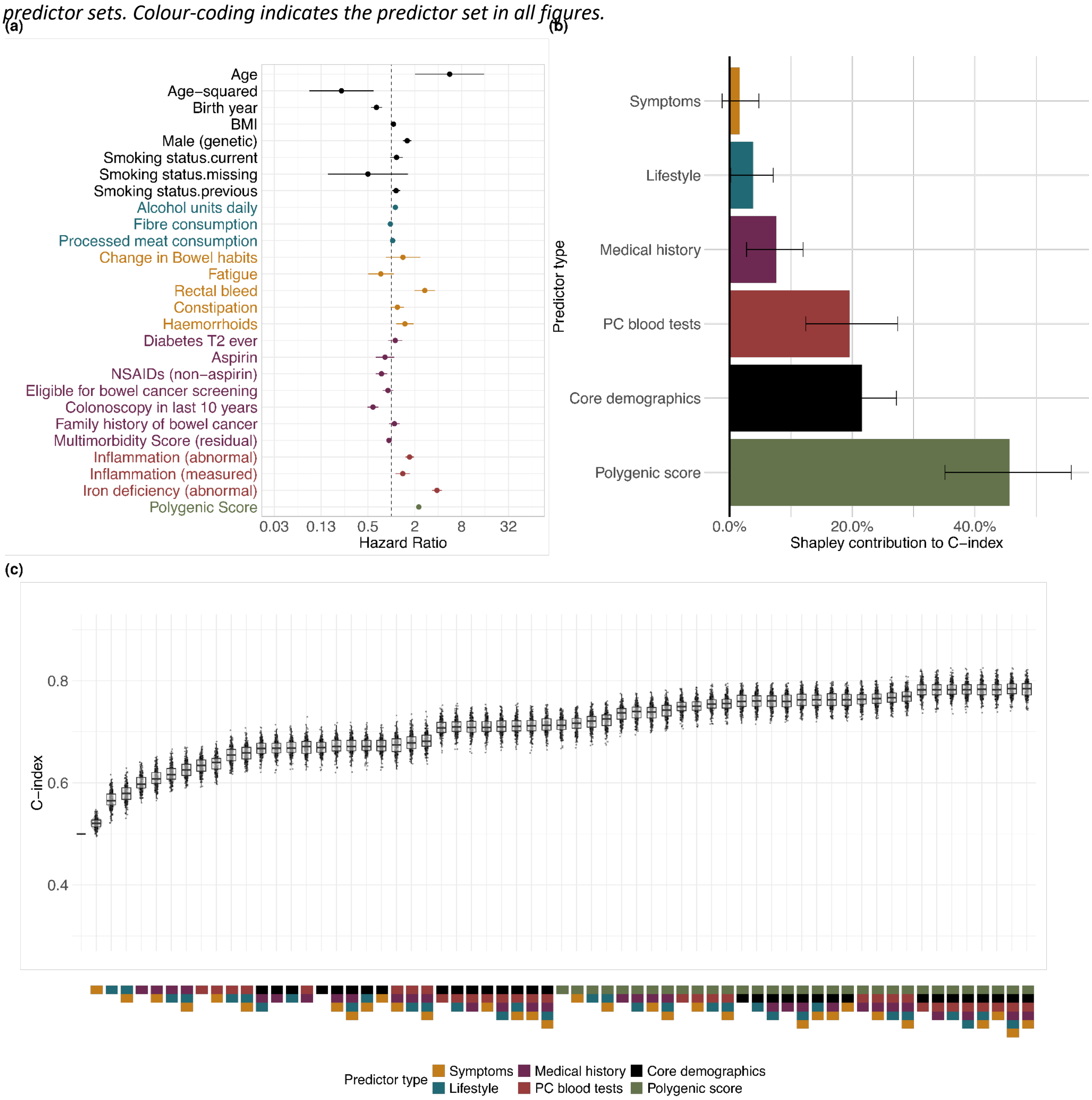

***Supplementary Figure 6b: Figure 6b-a*** *Hazard ratios from bidirectional stepwise Cox regression for the* ***symptomatic subcohort****, using* ***LDPred-generated polygenic scores****;* ***Figure 6b-b*** *The inclusion-order-agnostic discriminative contribution (C-index > 0.5) of each predictor set evaluated using Shapley values;* ***Figure 6b-c*** *C-indices from 200 bootstrap samples for each coalition of predictor sets. Colour-coding indicates the predictor set in all figures.*

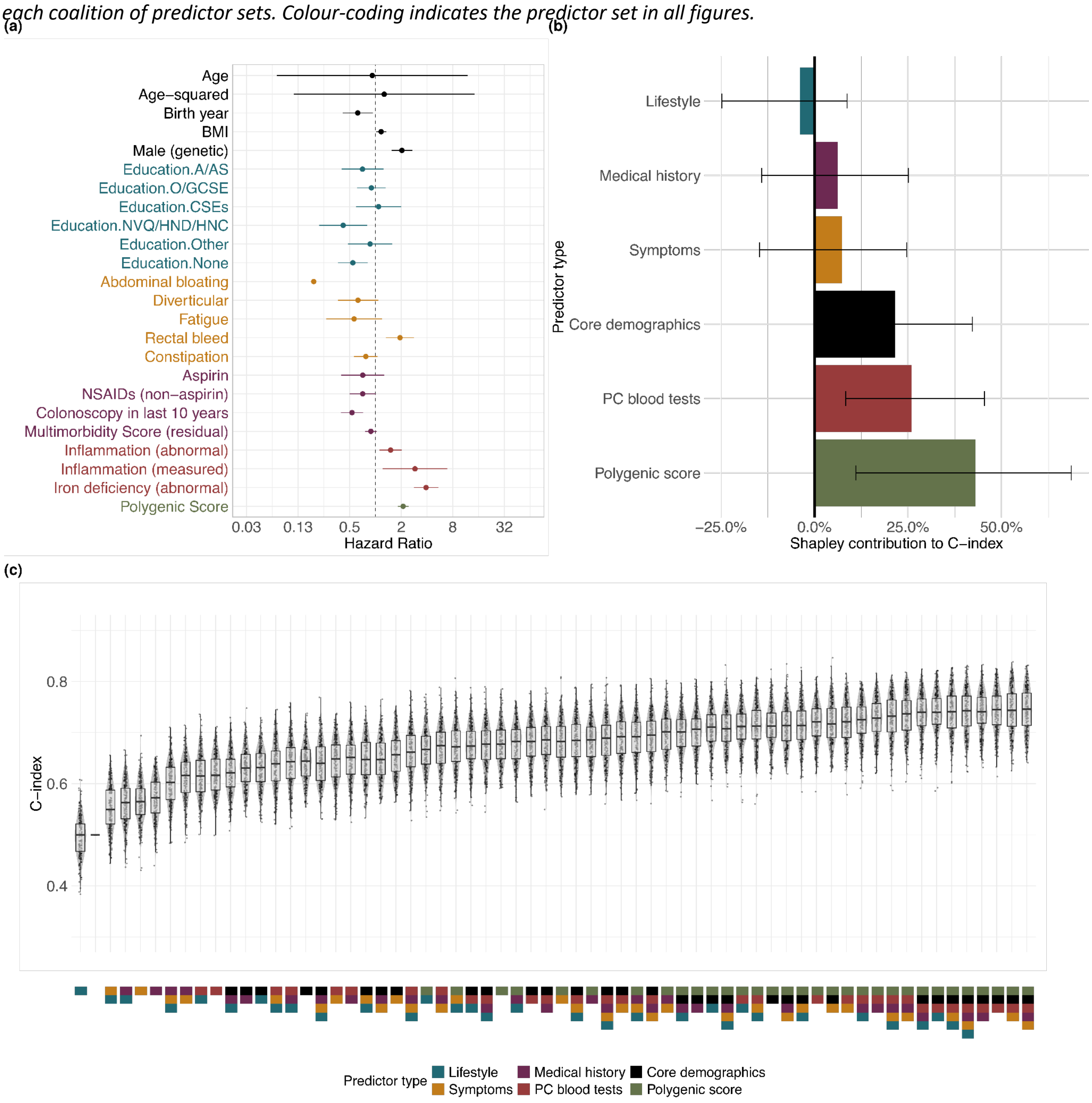

***Supplementary Figure 7:*** **Figure 7a** Hazard ratios from bidirectional stepwise Cox regression for the **"symptomatic" subcohort (N=70,241) defined as having any symptom in the predictor type "symptoms" sans fatigue**[21]; **Figure 7b** The inclusion-order-agnostic discriminative contribution (C-index > 0.5) of each predictor set evaluated using Shapley values; **Figure 7c** C-indices from 200 bootstrap samples for each coalition of predictor sets. Colour-coding indicates the predictor set in all figures.

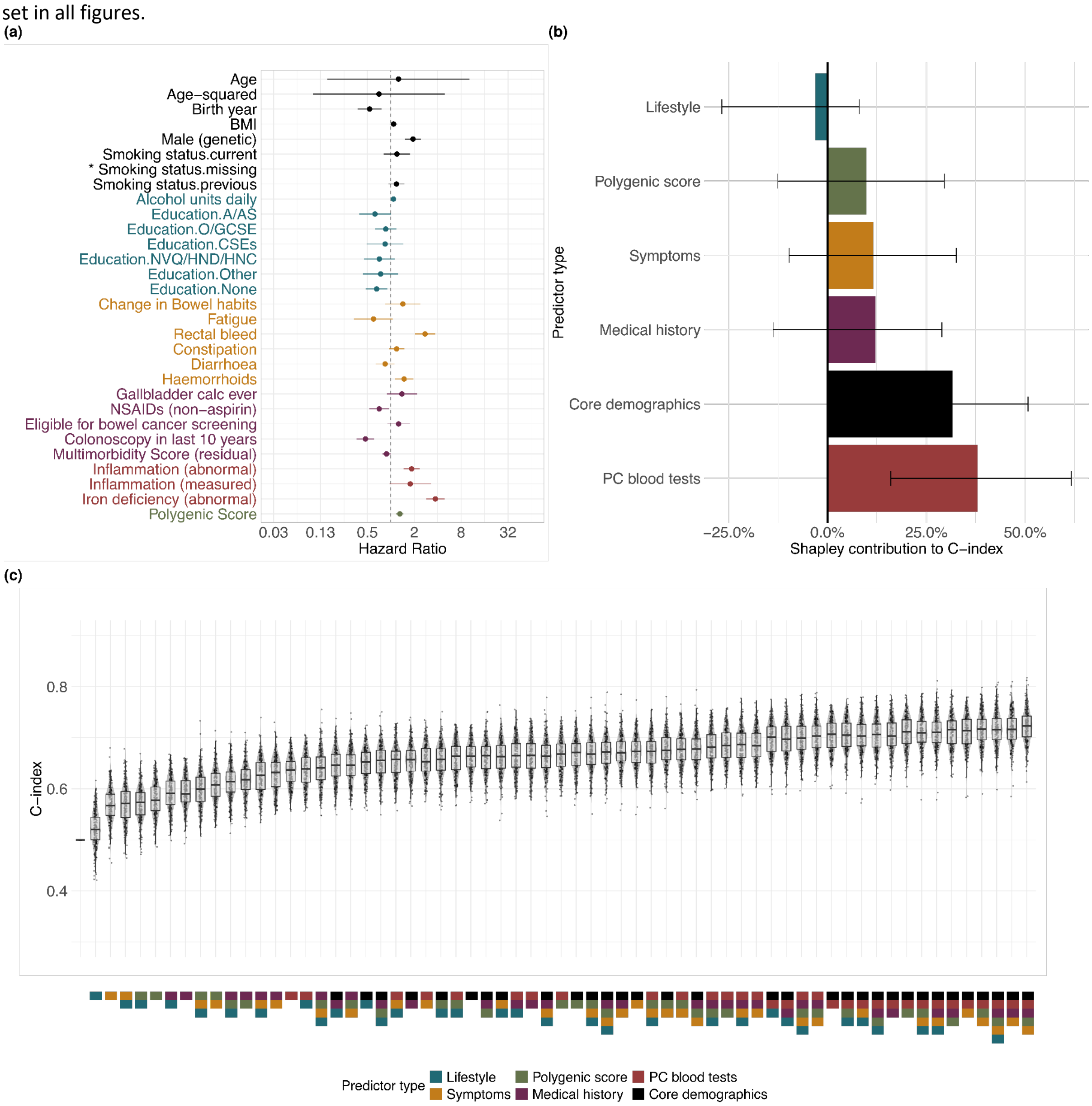

---

21 In this sensitivity analysis, we used a broader definition of the symptomatic subcohort, including any colorectal cancer (CRC) symptom listed in Table S1, excluding fatigue due to its lack of specificity. This is motivated by the fact that the symptomatic subcohort, as defined by bidirectional selection, comprises only 14% of the total case count in our study cohort, lower than what is typically reported in the literature (15).

***Supplementary Figure 8a:*** **Figure 8a-a** Hazard ratios from bidirectional stepwise Cox regression for the **study cohort but excluding participants with Vision as their GP data provider**; **Figure 8a-b** Discriminative contribution of predictors using Shapley values (C-index > 0.5); **Figure 8a-c** C-indices from 200 bootstrap samples for each coalition of predictor sets. Colour-coding indicates the predictor set in all figures.

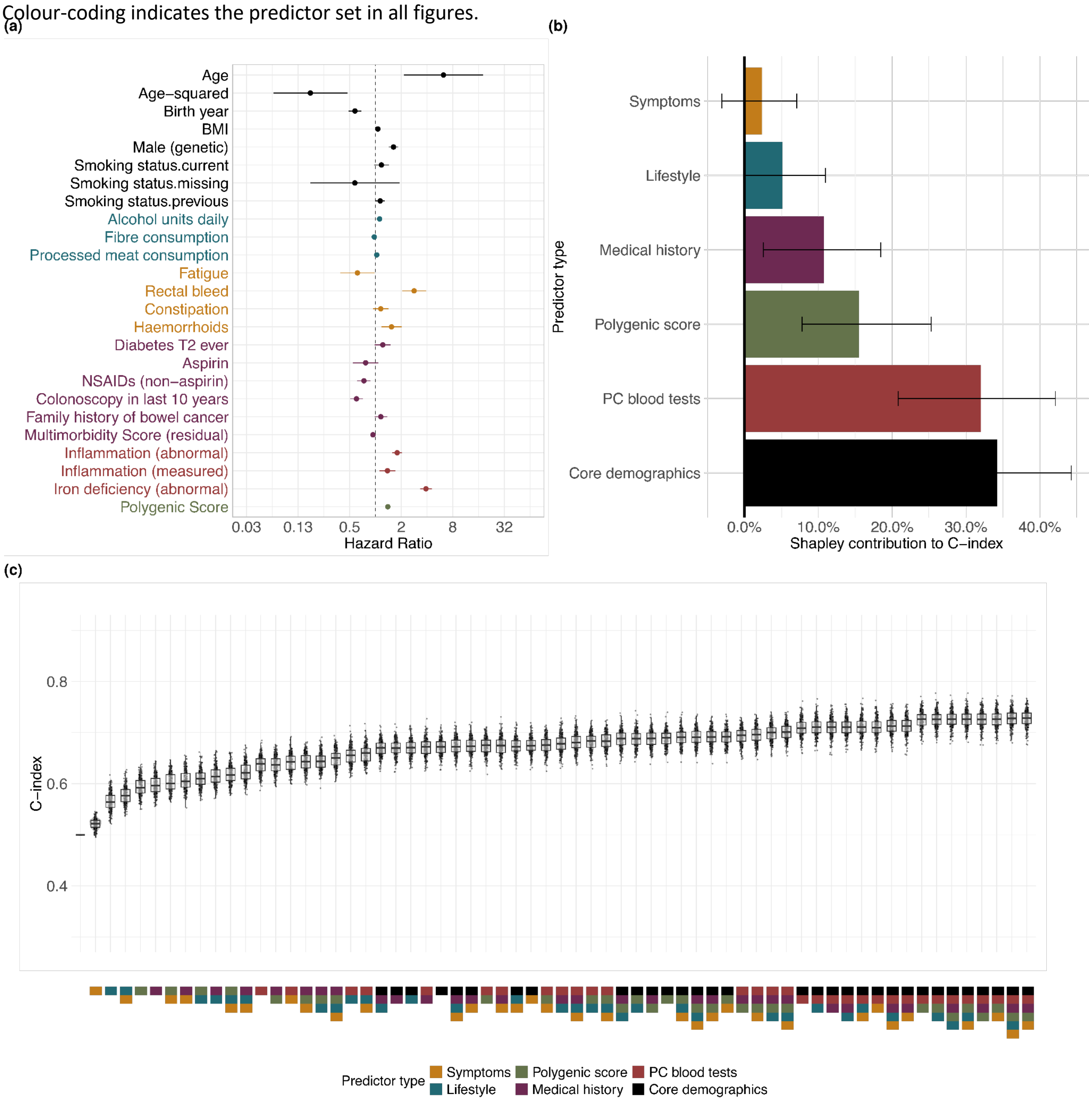

***Supplementary Figure 8b:*** **Figure 8b-a** Hazard ratios from bidirectional stepwise Cox regression for the **symptomatic subcohort but excluding participants with Vision as their GP data provider**; **Figure 8b-b** Discriminative contribution of predictors using Shapley values (C-index > 0.5); **Figure 8b-c** C-indices from 200 bootstrap samples for each coalition of predictor sets.

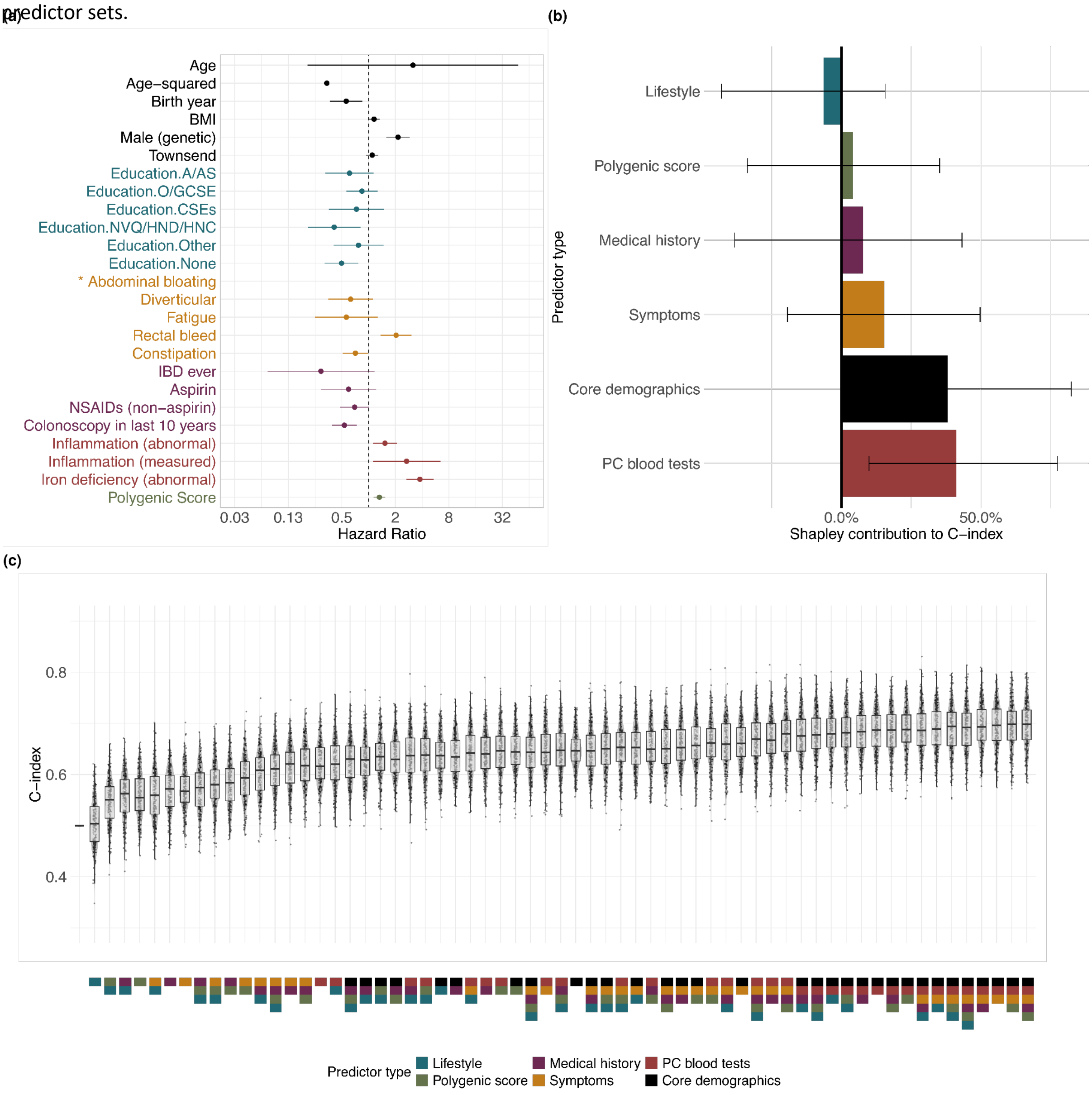

***Supplementary Figure 9:*** Calibration decile plots at 2 years for 10 random bootstrap validation samples of the (a) study cohort and (b) symptomatic subcohort, respectively.

(a) Study cohort

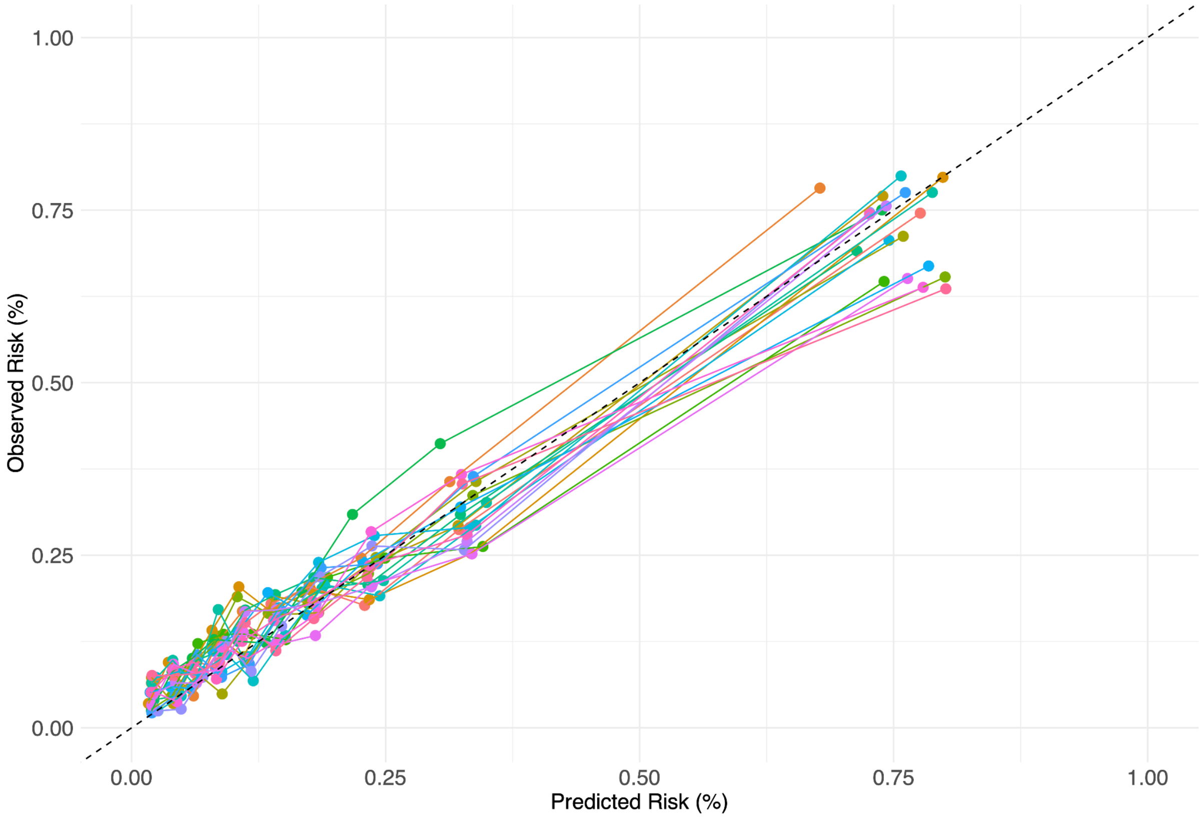

(b) Symptomatic subcohort

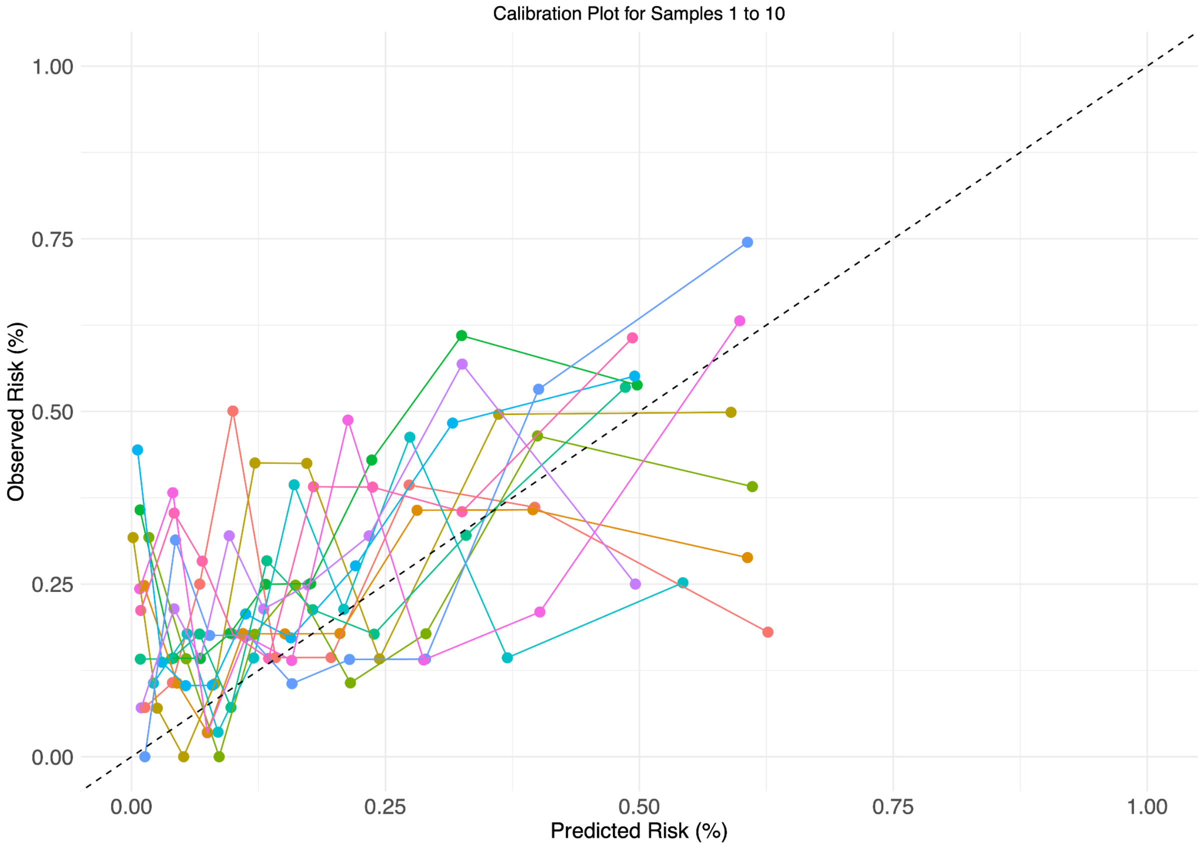

*References*

1. Paige E, Barrett J, Stevens D, Keogh RH, Sweeting MJ, Nazareth I, et al. Landmark Models for Optimizing the Use of Repeated Measurements of Risk Factors in Electronic Health Records to Predict Future Disease Risk. Am J Epidemiol. 2018 Jul 1;187(7):1530–8.

1. Paige E, Barrett J, Stevens D, Keogh RH, Sweeting MJ, Nazareth I, et al. Landmark Models for Optimizing the Use of Repeated Measurements of Risk Factors in Electronic Health Records to Predict Future Disease Risk. Am J Epidemiol. 2018 Jul 1;187(7):1530–8.

2. Houwelingen H van, Putter H. Dynamic Prediction in Clinical Survival Analysis. CRC Press; 2011. 250 p.
3. Chambers JM, Hastie T. Statistical Models in S [Internet]. Wadsworth & Brooks/Cole Advanced Books & Software; 1992. (Wadsworth & Brooks/Cole computer science series). Available from: https://books.google.co.uk/books?id=uyfvAAAAMAAJ
4. Venables WN, Ripley BD. Modern Applied Statistics with S [Internet]. New York, NY: Springer; 2002. (Chambers J, Eddy W, Härdle W, Sheather S, Tierney L, editors. Statistics and Computing). Available from: http://link.springer.com/10.1007/978-0-387-21706-2
5. step function - RDocumentation [Internet]. [cited 2023 Nov 10]. Available from: https://www.rdocumentation.org/packages/stats/versions/3.6.2/topics/step
6. Yuan M, Lin Y. Model Selection and Estimation in Regression with Grouped Variables. J R Stat Soc Ser B Stat Methodol. 2006 Feb 1;68(1):49–67.
7. Tibshirani R. The lasso method for variable selection in the Cox model. Stat Med. 1997 Feb 28;16(4):385–95.
8. Breheny P, Huang J. Group descent algorithms for nonconvex penalized linear and logistic regression models with grouped predictors. Stat Comput. 2015;25:173–87.
9. Breheny P, Huang J. Penalized methods for bi-level variable selection. Stat Interface. 2009;2:369–80.
10. Breheny P. The group exponential lasso for bi-level variable selection. Biometrics. 2015;71:731–40.
11. Shapley LS. A Value for N-Person Games [Internet]. RAND Corporation; 1952 [cited 2023 Jun 30]. Available from: https://www.rand.org/pubs/papers/P295.html
12. Lundberg SM, Lee SI. A Unified Approach to Interpreting Model Predictions. In: Guyon I, Luxburg UV, Bengio S, Wallach H, Fergus R, Vishwanathan S, et al., editors. Advances in Neural Information Processing Systems [Internet]. Curran Associates, Inc.; 2017. Available from: https://proceedings.neurips.cc/paper_files/paper/2017/file/8a20a8621978632d76c43dfd28b67767-Paper.pdf
13. Thomas M, Sakoda LC, Hoffmeister M, Rosenthal EA, Lee JK, van Duijnhoven FJB, et al. Genome-wide Modeling of Polygenic Risk Score in Colorectal Cancer Risk. Am J Hum Genet. 2020 Sep 3;107(3):432–44.
14. UK Biobank. UK Biobank: Primary Care Linked Data [Internet]. 2019 [cited 2022 Feb 24]. Available from: https://biobank.ndph.ox.ac.uk/ukb/ukb/docs/primary_care_data.pdf
15. Zakkak N, Barclay M, Swann R, McPhail S, Rubin G, Abel G, et al. The Presenting Symptom Signatures of Incident Cancer: Evidence from the English 2018 National Cancer Diagnosis Audit. British Journal of Cancer.
