## Appendix 2 for "Genetics, primary care records and lifestyle factors for short-term dynamic risk prediction of colorectal cancer: prospective study of asymptomatic and symptomatic UK Biobank participants"

**Appendix 2**
**CRC Model Development: Variables**

**Contents**

Introduction to the Appendix
Section A: UKB Baseline Data
Part I: Phenotypic Data
Part II: Genetic Data
Section B: Primary Care Records
Part I: Background
Part II: Generation of Codelists
Part II: Methods for Variables
Method 1: Ever Event
Method 2: Recent Event
Method 3: New Onset Event
Method 4: Rate of (Recent) Event
Method 5: Regular Event
Method 6: Test Results – Gold Standard and Proxy Measures
Implementation of a Multimorbidity Score
Section C: Analysis Groupings

**Introduction to the Appendix**

The variables used in the development of these models are developed using data provided by UK Biobank (UKB) for the individuals in the cohort. We use both data collected during the baseline assessment (phenotypic and genetic) and data available from the linked primary care records (includes both GP consultation records and prescription records).

Within this document, we provide:

- a description of the data used
- the methods used to extract clinically relevant variables
- a description of the selected set of variables

For each variable used in the models, we give the justification for inclusion in a model predicting risk of colorectal cancer. In order to take advantage of existing knowledge of colorectal cancer risk, and in line with guidance for reducing risk of bias when selecting variables for prognostic models [1], we have based our initial variable selection on previously published literature and have been guided by expert opinion.

**Section A: UKB Baseline Data**

*Part I: Phenotypic Data*

All participants in UKB attended a baseline assessment that included completion of questionnaires about demographics, lifestyle and their medical history, as well as the measurement of a range of physical characteristics [2]. Data on cancer incidence are available for UKB participants through linkage to national cancer registries. Although follow-up questionnaires were circulated to the participants after baseline (for example, gathering information on participant occupation) these were not completed for the whole cohort and have not been used in this analysis.

We note that there are two types of data available from the UKB baseline assessment. These can be split into the following types:

- Externally non-modifiable data, such as sex, age and ethnicity. This data can be used with confidence in predictions made at any point in the life of the participant.
- Modifiable or changeable data, including information on lifestyle (such as smoking, BMI, dietary variables) and medical history (such as self-reported current medications or family history of bowel cancer). These variables may change during the follow-up period used in the analysis (for example, if an individual quits smoking after baseline) and should be used and interpreted with caution.

Some of the risk factors drawn from the UKB baseline assessment (see the UKB data showcase for details [3]) can be used directly in the analysis. In other cases, however, it was necessary to create composite variables to describe a risk factor of interest. For example, our variable for total alcohol consumption is calculated by combining self-reported consumption of a range of different alcoholic beverages, for which participants were asked for either a weekly or monthly consumption estimate (see Fig. 1).

Similar approaches were taken to create summary variables for red and processed meat consumption. In other cases, approaches previously described in literature were used. For example, the partial fibre score developed by Bradbury et al. [4] was calculated by combining self-reported data from eight dietary questions (relating to consumption of fruit, vegetables, bread and cereals).

Details of the derivation of all the variables that use data from the UKB baseline assessment, and justification for their inclusion in the analysis, can be found in Table 1 and the accompanying spreadsheet.

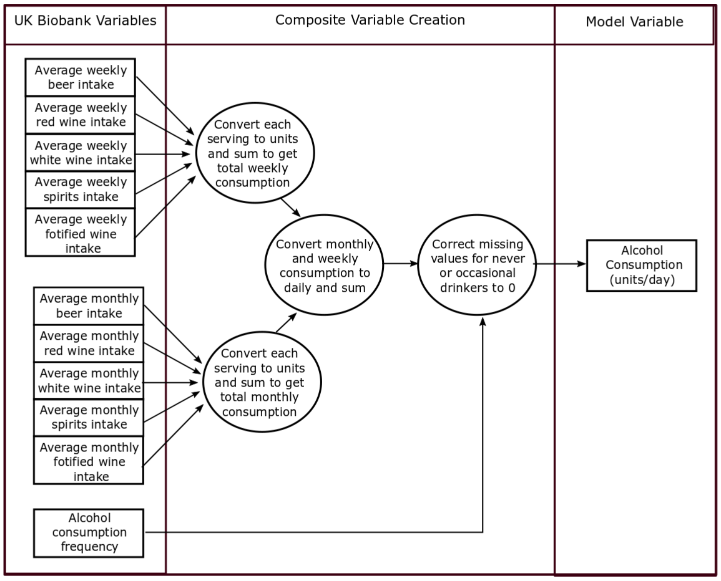

UK Biobank Variables
Composite Variable Creation
Model Variable
Average weekly beer intake
Average weekly red wine intake
Average weekly white wine intake
Average weekly spirits intake
Average weekly fotified wine intake
Convert each serving to units and sum to get total weekly consumption
Convert monthly and weekly consumption to daily and sum
Correct missing values for never or occasional drinkers to 0
Alcohol Consumption (units/day)
Average monthly beer intake
Average monthly red wine intake
Average monthly white wine intake
Average monthly spirits intake
Average monthly fotified wine intake
Convert each serving to units and sum to get total monthly consumption
Alcohol consumption frequency


[illegible]

**Table 1: UKB Phenotypic Variables using Baseline Data**

| Model Variable | UKB variables | Method | Units | Categories | Missing Data (%) | Reference for association with CRC (or other justification for inclusion) |
|---|---|---|---|---|---|---|
| Age | 34-0.0 Year of birth<br>52-0.0 Month of birth<br>53-0.0 Date at baseline | For each index date (which must be post-baseline) at which an individual is entered into the model, their age is calculated. | Years | - | 0 | https://www.nhs.uk/conditions/bowel-cancer/causes/ |
| Birth Year | 34-0.0 Year of birth | Direct mapping | - | - | 0 | Accounts of changes to the healthcare system and lifestyle over time. |
| Sex | 21022-0.0 Genetic sex | Direct mapping | - | 1. Female<br>2. Male | 0 | Men are known to be at higher risk of colorectal cancer |
| Ethnicity | 21000-0.0 Ethnic background | Grouped participants using the six broad ethnic groupings given by UK Biobnk – ignoring any sub-groupings within this variable. Given the small numbers in both, individuals who self-reported Chinese background were grouped with those who self-reported other ethnic backgrounds. | - | 1. White<br>2. Mixed background<br>3. SE Asian<br>4. Black<br>5. Other (including Chinese) | 0.3* | Incidence of bowel cancer is known to be higher in white individauls than in several other ethnic groups. https://www.cancerresearchuk.org/health-professional/cancer-statistics/statistics-by-cancer-type/bowel-cancer |
| Education Level | 6138-0.0 Qualifications (highest obtained) | Direct mapping – see https://biobank.ndph.ox.ac.uk/showcase/coding.cgi?id=100305 for more details.<br><br>Qualifications listed in category column (or equivalent) | - | 1. Degree<br>2. A-levels<br>3. GCSes<br>4. CSEs<br>5. Vocational<br>6. Professional<br>7. None | 2.02 | |
| Deprivation | 189-0.0: Townsend deprivation index at recruitment | Direct mapping | - | - | 0 | |
| BMI | 21000-0.0 body mass index | Direct mapping | $kg/m^2$ | - | 0 | https://www.nhs.uk/conditions/bowel-cancer/causes/ |

| | | | | | | |
|---|---|---|---|---|---|---|
| Smoking Status | 20116-0.0 smoking status | Direct mapping | - | 1. Current<br>2. Former<br>3. Never | 0.3* | https://www.nhs.uk/conditions/bowel-cancer/causes/ |
| Alcohol Consumption | 4407-0.0 Average monthly red wine intake<br>4418-0.0 Average monthly champagne plus white wine intake<br>4429-0.0 Average monthly beer plus cider intake<br>4440-0.0 Average monthly spirits intake<br>4451-0.0 Average monthly fortified wine intake<br>4462-0.0 Average monthly intake of other alcoholic drinks<br>1568-0.0 Average weekly red wine intake<br>1578-0.0 Average weekly champagne plus white wine intake<br>1588-0.0 Average weekly beer plus cider intake<br>1598-0.0 Average weekly spirits intake<br>1608-0.0 Average weekly fortified wine intake<br>5364-0.0 Average weekly intake of other alcoholic drinks | UKB participants were asked about their alcohol consumption either weekly or monthly depending on their answer to a previous question (e.g. "In an average month, how many glasses of red wine to you drink?").<br><br>First, total weekly and monthly unit consumption are calculated. The conversions used to calculate units from self-reported servings is given in Table 1(b).<br><br>Weekly and monthly totals are then converted to units/day and combined into a single variable. | Units (UK, 8g)/day | - | 0.33 | https://www.nhs.uk/conditions/bowel-cancer/causes/ |
| Processed Meat Consumption | 1349-0.0 Processed meat intake | UKB participants were asked about their weekly processed meat consumption. Their responses were converted for analysis (see Table 1(a)). | Servings/ week | - | 0.45 | https://www.nhs.uk/conditions/bowel-cancer/causes/ |
| Red Meat Consumption | 1369-0.0 Beef intake<br>1379-0.0 Lamb/mutton intake<br>1389-0.0 Pork intake | UKB participants were asked about their weekly beef, lamb and pork consumption. | Servings/ week | - | 0.37 | https://www.nhs.uk/conditions/bowel-cancer/causes/ |

| | | | | | | |
|---|---|---|---|---|---|---|
| | | Their responses were converted to portions using the same methods as for processed meat (see Table 1(a)).<br><br>Then all three variables were summed to get total red meat consumption per week. | | | | |
| Fibre Consumption | 1289-0.0 Cooked vegetable intake (1.51%)<br>1299-0.0 Salad / raw vegetable intake (1.56%)<br>1309-0.0 Fresh fruit intake (0.65%)<br>1319-0.0 Dried fruit intake (1.38%)<br>1438-0.0 Bread intake (2.06%)<br>1448-0.0 Bread type (4.2%)<br>1458-0.0 Cereal intake (0.64%)<br>1468-0.0 Cereal type (17.4%) | An estimate of fibre consumption is made by summing together the contributions from a range of different dietary sources, using the "partial fibre score" developed by Bradbury et al. [4] for use in UK Biobank data (which has previously been shown to reliably rank participants according to fibre intake). More detail on diet is not available for the whole UKB cohort, which limits the types of fibre that are included in this score.<br><br>Each type of fibre is converted into a daily average before they are summed together. | Servings of fibre/day | - | 0.21 | A higher fibre diet is known to be protective for colorectal cancer. |
| Family history of bowel cancer | 20107-0.0 to 20107-0.9 father's disease history<br>20110-0.0 to 20110-0.10 mother's disease history<br>20111-0.0 to 20111-0.11 sibilings' disease history | Set variable to 1 if any family member has a history of bowel cancer (otherwise 0). | - | 0 - no<br>1 - yes | n/a | https://www.nhs.uk/conditions/bowel-cancer/causes/ |
| Family history of breast cancer | 20107-0.0 to 20107-0.9 father's disease history<br>20110-0.0 to 20110-0.10 mother's disease history<br>20111-0.0 to 20111-0.11 sibilings' disease history | Set variable to 1 if any family member has a history of breast cancer (otherwise 0). | - | 0 - no<br>1 - yes | n/a | |

| | | | | | | |
|---|---|---|---|---|---|---|
| Family history of lung cancer | 20107-0.0 to 20107-0.9 father's disease history<br>20110-0.0 to 20110-0.10 mother's disease history<br>20111-0.0 to 20111-0.11 sibilings' disease history | Set variable to 1 if any family member has a history of lung cancer (otherwise 0). | - | 0 - no<br>1 - yes | n/a | |
| Eligible for bowel screening | 21003-0.0 Age at assessment<br>53-0.0 Date at assessment | At each index date, determine if a person was eligible for bowel cancer screening based on the date (after 01-01-2008) and their age (60-74 years) at the start of the 2-year lookback period. | - | 0 - no<br>1 - yes | | People who have undergone bowel cancer screening have a lower risk of colorectal cancer. |

**Table 1(a)**

| UKB coding for meat variables | Description (how often do you eat …?) | Analysis portions |
|---|---|---|
| 0 | Never | 0 |
| 1 | Less than once a week | 0.5 |
| 2 | Once a week | 1 |
| 3 | 2-4 times a week | 3 |
| 4 | 5-6 times a week | 5.5 |
| 5 | Once or more daily | 7 |
| -1 | Do not know | NA |
| -3 | Prefer not to answer | NA |

**Table 1(b)**

| Alcohol type | Units/serving |
|---|---|
| Red wine | 2.1 |
| White wine | 2.1 |
| Fortified wine | 2.4 |
| Beer | 2.5 |
| Spirits | 1.0 |
| Other | 1.5 |

*Part II: Genetic Data*

Full genotype information is available for 488,377 members of UKB cohort. The blood samples (taken during baseline assessment) of the UKB participants were genotyped using Affymetrix UK BiLEVE Axiom Array and Affymetrix UK Biobank Axiom array and imputed to the combined 1000 Genomes Project v.3 and UK10K reference panels using SHAPEIT3 and IMPUTE3 [5]. This yielded data on approximately 96 million single nucleotide polymorphisms (SNPs).

A polygenic score (PGS) was used to quantify genetic predisposition for CRC; PRS-CSx was developed using a cohort of  European and East Asian ancestry individuals – 55,105 cases with CRC and 65,079 controls - and includes 1,145,689 SNPs [6]. A sensitivity analysis using a different PGS, LDPred, which was derived using only individuals with European ancestry  - 35145 cases with CRC and 288,934 controls - and consisting of 1,180,765 SNPs [7]. We note that some of the development population used for the LDPred PGS was drawn from the UK Biobnk cohort, so use of this score risks overfitting in this analysis.

Principal component analysis is used to characterise population structure and to identify individuals with similar ancestry. Twenty principal components have been calculated (by UK Biobank) for this cohort – and have been found to align well with self-reported ethnicity. Details of how the principal components were calculated are provided by UK Biobank [8].  In our analysis, we included the first ten principal components to adjust for distinct ancestries and ethnic backgrounds. We include theseprincipal components within the “polygenic score” set of predictors - and required their inclusion in the final model (if the PGS was selected) - but do not report these as predictors in our main figures and tables. Readers will find, however, the hazard ratios for these ten principal components included in the relevant tables in the supplementary materials.

**Section B: Primary Care Records**

*Part I: Background*

Linked primary care records are currently available for around 45% of the UKB cohort (n=197,939) [9]. These data include both records of clinical events and prescriptions. An additional file is also provided which gives the registration records (dates of joining and leaving each practise) for all the included individuals.

As described in the UKB documentation on “Primary Care Linked Data” [9], the records are provided by four different data suppliers (see Table 2). The four different providers cover different regions and use distinct coding systems. Differences in the data extracted from each data provider is expected, given the known regional variation in healthcare and health outcomes in the UK. Furthermore, different coding frameworks are used across the different regions and data providers, so some system variation is also expected.

We note, that the linkage for English Vision practises is incomplete, as people registered with English Vision who have died before the end of the linkage period have mostly not had their data provided.

Using the estimates from UKB, this is likely to have excluded about 500-600 people who died between baseline and 2017, and this could introduce selection bias into the analysis cohort. To account for this, our analysis includes a sensitivity analysis without English Vision patients. However, this sensitivity analysis uses a cohort with a different geographical distribution (a higher relative proportion of Welsh and Scottish participants) and also changes the realtivel proportions of the coding frameworks used; these differences should be considered when interpreting the results.

**Table 2: UKB Primary Care Data Providers**

| # | Country | Data Provider | Number of participants (approx.) | Clinical records coding | Prescription records coding |
|---|---|---|---|---|---|
| 1 | Scotland | EMIS & Vision | 27,000 | readv2 | readv2/BNF(6-character) |
| 2 | Wales | EMIS & Vision | 21,000 | readv2 | readv2 |
| 3 | England | TPP | 165,000 | CTV3 | BNF (10-character) |
| 4 | England | Vision | 18,000 | readv2 | readv2/dmd |

*Part II: Codelists*

The primary care data for the UKB are provided in a row-per-event format, where each individual may appear in the dataset multiple times, for different clinical events (see the dummy data in Fig. 2). For each event, the dataset provides the participant ID, the date the event was recorded, the data provider and a code that defines the event type. By using a list that includes codes that describe a certain type of clinical occurrence (e.g. abdominal pain, or haemorrhoid medication), known as a “codelist”, the datasets can be queried to identify all events of that type. Some events (e.g, Xa96v – “Haemoglobin concentration”, in Fig. 2) also include a value field (20mg/l), which may provide additional useful information.
Where possible existing codelists, from (i) previously published studies, (ii) previously developed by members of the collaboration or (iii) available online (for example through the OpenCodelist resource: https://www.opencodelists.org/), were used. References for existing codeslists are given in the accompanying spreadsheet to this appendix. Where new codelists were developed or existing codelists

[illegible]

| ID | Date | Data Provider | Read Code | CTV3 code | Value |
|---|---|---|---|---|---|
| 10002 | 01/05/1985 | 3 | 1829. | *NA* | *NA* |
| 10002 | 08/09/2001 | 3 | 25CA. | *NA* | *NA* |
| 10002 | 10/06/2013 | 3 | 9EV5. | *NA* | *NA* |
| 10002 | 22/09/2016 | 2 | *NA* | XM125 | *NA* |
| 20685 | 18/03/2015 | 4 | *NA* | Xa96v | 40 |
| 20685 | 18/03/2015 | 4 | *NA* | .42d4 | 200 |
| ….. | … | … | … | … | … |

augmented, relevant codes were identified through automated searching of UKB dictionaries for each relevant coding frameworks [10] and then checked by a member of the team with clinical expertise.

When generating (or adapting existing) codelists for a specific event of interest (e.g. abdominal mass), it is also necessary to convert between the different coding frameworks used by the four data providers (see Table 2). All identified codes from one coding framework had to be converted to all other coding frameworks to ensure full and comprehensive coverage. For the clinical data, this simply required a conversion between readv2 and ctv3 (and vice versa), using a look-up table provided by UKB [10], but the process for prescription data was more complex (see Fig. 3).

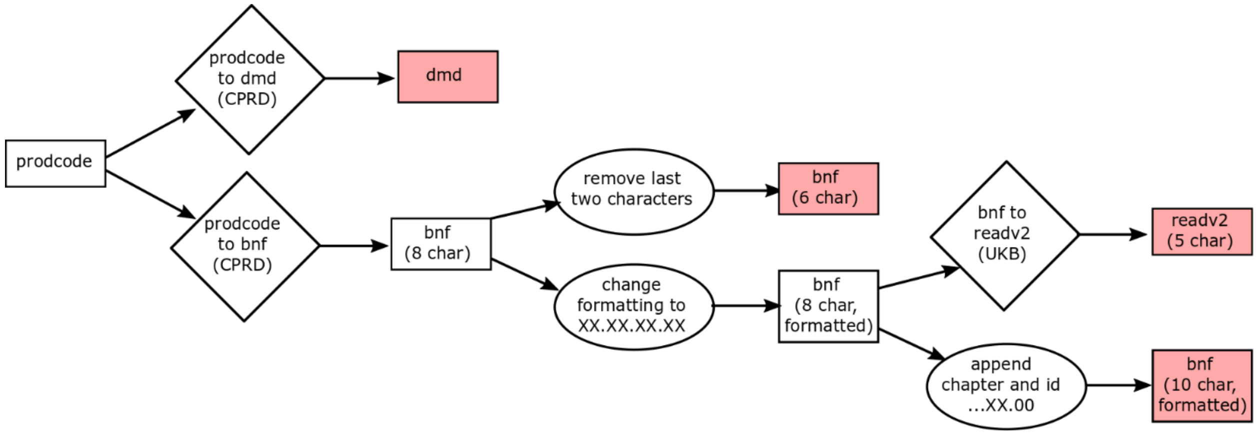


[illegible]

Given the differences in scope of codes from the different frameworks, this process can introduce some irrelevant codes into the codelists, so a final manual check was carried out on all codelists before they were used in analysis.

When using and interpreting primary care data we must consider not only that it may be incomplete, but also that the coding of clinical events may have changed over time. For example, bowel cancer screening was introduced in 2006, but initially coding these screening events GP records was sparse. However, these events have become more routinely recorded in recent years (see Fig. 4).

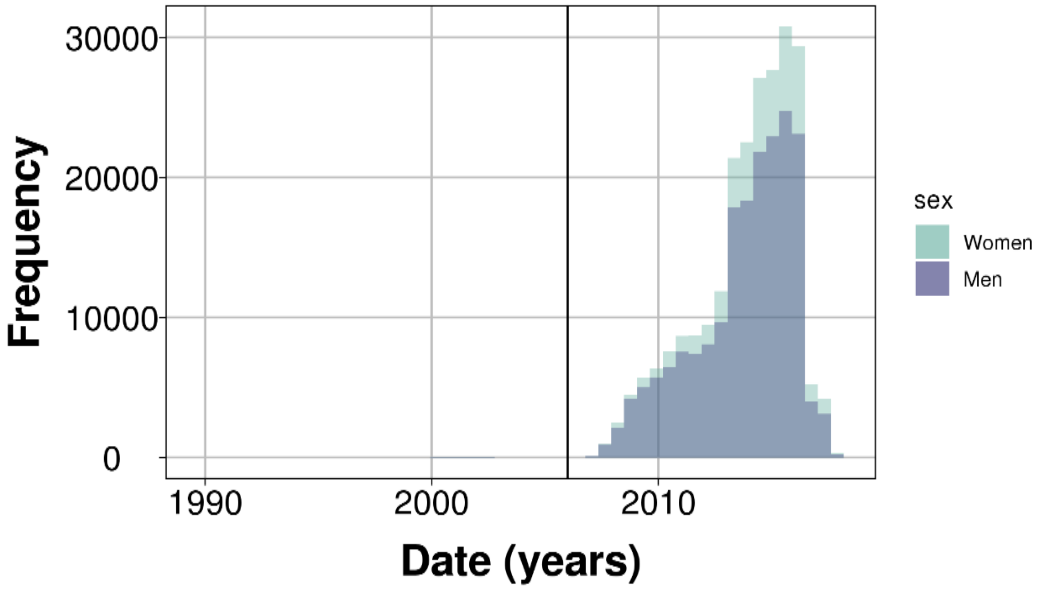


*Figure 4: Number of events related to bowel cancer screening coded in the UKB linked GP clinical records. The black line represents the introduction of bowel cancer screening in England in 2006.*

*Part III: Methods for Predicitors*

Various methods were developed to generate clinically relevant predictors. In a recent study by Carney et al. [11], it was highlighted that when using electronic health records, careful consideration regarding the time in which events are recorded is needed. In particular, events that occur shortly before a cancer diagnosis (including symptoms) are considered as “markers” of underlying disease, whereas events that occur some time in the past may instead by considered as a “modifier” of the risk of an individual developing cancer.

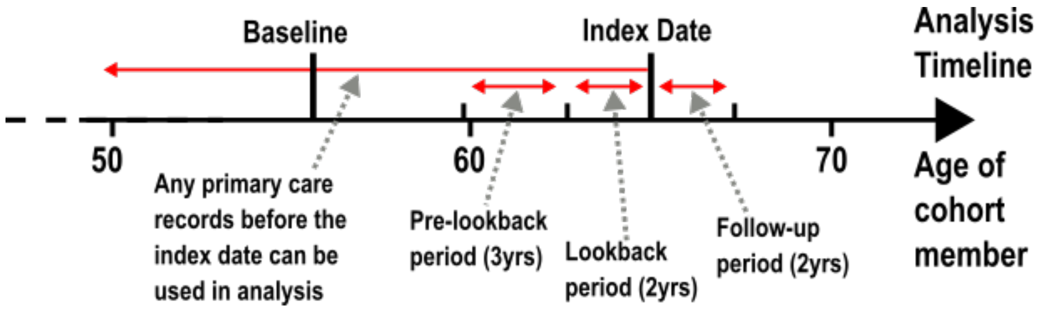


[illegible]

In our analysis, the variables were developed with reference to a point in time (the index date – equivalent to the landmark ages) at which the modelling is carried out. Description of the superlandmark framework and how index dates are defined for each cohort member can be found in the the main manuscript with additional details in Supplementary Methods 1. Primary care records dated before the index date can be used in the analysis, with the period of interest - the “lookback period” - being determined based on the clinical rationale for including each variable in the model. In all cases, index dates are after the date of baseline assessment for that individual and before the end of the period covered by their available GP records.

***Method 1: Ever events***

For some variables, we are interested if an event of this type has *ever been recorded* for each individual. This was typically used in this analysis to identify long-term conditions, which may modify the risk of an individual developing colorectal cancer (e.g. type 2 diabetes). A binary variable is created that distinguishes between individuals with at least one record of an event (1) and no records of an event (0) at any point in time before the index date.

$$\text{Event} = \begin{cases} 1, & \text{if there is} \geq 1 \text{ instance of the event before the index date} \\ 0, & \text{if there are no recorded instances of the event before the index date} \end{cases}$$

**Table 3: List of predictors defined as ever events extracted from UKB primary care records**

| *Predictor* | *Shortname* | *Reference for association (or other justification of inclusion)* |
|---|---|---|
| Diagnosis of gallstones or cholelithiasis | Gallbladder Calc | Benign gastrointestinal conditions (including gallstones) [12] and gall bladder disease specifically [13], have been shown to be associated with increased risk of colorectal cancer. |
| Diagnosis of inflammatory bowel disease | IBD | Inflammatory bowel disease is a well-established risk factor for colorectal cancer [14]. |
| Diagnosis of type 1 diabetes | Type 1 diabetes | There is some evidence that type 1 diabetes is associated with colorectal cancer [15]. |
| Diagnosis of type 2 diabetes | Type 2 diabetes | Type 2 diabetes is a well-established risk factor for many cancer types, including colorectal cancer [16, 17]. |

***Method 2: Recent events***

For other predictors, we are only interested in events that have occurred recently. This methods was used when the presence of a single instance event in the recent past may be a marker of underlying disease (for example, rectal bleeding is an alarm symptom of colorectal cancer) or where a particular event may be protective a few years after it has occured (for example, a colonoscopy). For each variable, a "lookback window" from the index date is used, with the length of the window depending on how the event is associated with colorectal cancer. For example, symptoms that may indicate underlying (but not yet diagnosed) colorectal cancer are defined over a short (2-year) lookback window. A binary variable is created, distinguishing between individuals with at least one record (1) and no records (0) of the event in the lookback window.

$$\text{Recent event} = \begin{cases} 1, & \text{if there is} \geq 1 \text{ instance of event in the lookback period for an index date} \\ 0, & \text{if there are no instances of event in the lookback window for an index date} \end{cases}$$

**Table 4: List of predictors defined as recent events extracted from UKB primary care records**

| **Variable** | **Lookback Period** | **Shortname** | **Reference for association (or other justification of inclusion)** |
|---|---|---|---|
| Recent abdominal lump | 2 years | Abdominal Lump | Abdominal lumps (or masses) are a well-established alarm symptom of colorectal cancer [18, 19]. |
| Recent rectal mass | 2 years | Rectal Mass | Rectal lumps (or masses) are a well-established alarm symptom of bowel cancer [18, 19]. |
| Recent change in bowel habit | 2 years | Change in Bowel Habit | A recent change in bowel habit is a well-established alarm symptom of colorectal cancer [18, 19]. |
| Recent rectal bleeding | 2 years | Rectal Bleeding | A recent change in bowel habit is a well-established alarm symptom of colorectal cancer [18, 19]. |
| Recent colonoscopy or sigmiodoscopy | 10 years | Colonoscopy | Individuals who have a colonoscopy or sigmoidoscopy (either as part of bowel cancer screening or otherwise) are more likely to have preventative interventions (such as the removal of polyps) that can reduce the risk of colorectal cancer [20]. |

***Method 3: New onset events***

For other events, they are only of interest if they have occurred in the recent past *and* indicate a change in the health status of an individual. This method was used for symptoms that may be present for many people in the population (for example due to chronic conditions) but may indicate underlying prevalent colorectal cancer if they have recently been reported for the first time. This method was also used to identify recent diagnoses which could be potentially be misdiagnoses of symptoms caused by underlying colorectal cancer. To determine an instance of a new onset event , we must define both a lookback window in which the event occurs (as in the previous "recent event" method) and a prior period in which no events of this type are recorded. A binary variable is created, distinguishing between individuals with a record of the event in the lookback period *and* no record of the event in the preceding period (0) and all other individuals (1).

$$\text{New onset event} = \begin{cases} 1, & \text{if} \geq 1 \text{ events in the lookback and no instances in the preceding period} \\ 0, & \text{for everyone else} \end{cases}$$

**Table 5: List of predictors defined as new onset events extracted from UKB primary care records**
**combines GP records and prescription records (see box 1)*

| **Variable** | **Lookback Period** | **Preceding Period** | **Shortname** | **Reference for association (or other justification of inclusion)** |
|---|---|---|---|---|
| New onset constipation* | 2 years | 3 years | Constipation | New onset constipation may indicate a change in bowel habit [19]. |
| New onset diarrhoea* | 2 years | 3 years | Diarrhoea | New onset diarrhoea may indicate a change in bowel habit [19]. |
| New onset haemorrhoids* | 2 years | 3 years | Haemorroids | A recent diagnosis of haemorroids may indicate a change in bowel habit, and symptoms of colorectal cancer could have be misattributed [19]. |
| New onset weight loss | 2 years | 3 years | Weight Loss | Weight loss is a non-specific symptoms of late stage cancer [18]. |
| New onset jaundice | 2 years | 3 years | Jaundice | Jaundice can be a symptom of metastatic colorectal cancer [21, 22]. |
| New onset fatigue | 2 years | 3 years | Fatigue | Fatigue is a non-specific symptoms of cancer and can be caused by anaemia (which may be caused by underlying cancer) [18, 22]. |
| Recent diagnosis of diverticular disease | 2 years | 3 years | Diverticular Disease | A recent diagnosis of diverticular disease may indicate underlying colorectal cancer; this may be |

| | | | | |
|---|---|---|---|---|
| | | | | caused by misattribution of cancer symptoms [12, 23]. |
| Recent diagnosis of irritable bowel syndrome | 2 years | 3 years | IBS | A recent diagnosis of IBS may indicate underlying colorectal cancer; this may be caused by misattribution of cancer symptoms [12]. |

***Method 4: Incidence of (recent) event***

For other variables, the number of times a recent event is recorded may be of interest. For example, for mild or vague symptoms (such as abdominal pain) repeated events may indicate persistent or severe symptoms. This method was used for symptoms that may indicate bowel cancer, but will in the vast majority of cases have benign explanation. For example, there are many possible causes of stomach disorders, however, in a person with underlying colorectal cancer this could be caused by a bowel obstruction.

As we are again looking at symptoms, as described previously, a lookback window was used (2 years) so that only recent events contributed to the analysis. As large numbers of events of the same type are relatively rare within this cohort (and could distort any association) we truncated the maximum number of events to four for each individual at each index date. A categorical variable is created, with possible values of 0 (no events), 1, 2, 3 or 4 (four or more events).

$$\text{Number of times event recorded} = \begin{cases} 4, & \text{if 4 or more events in the lookback period} \\ 3, & \text{if 3 events in the lookback period} \\ 2, & \text{if 2 events in the lookback period} \\ 1, & \text{if 1 event in the lookback period} \\ 0, & \text{if no events in the lookback period} \end{cases}$$

**Table 6: List of predictors defined as the incidence of events extracted from UKB primary care records**

| **Variable** | **Lookback Period** | **Truncation Number** | **Shortname** | **Reference for association (or other justification of inclusion)** |
|---|---|---|---|---|
| Number of times abdominal bloating reported | 2 years | 4 | Abdominal Bloating | Abdominal bloating can indicate bowel obstruction [19, 22]. |
| Number of times abdominal pain reported | 2 years | 4 | Abdominal Pain | Abdominal pain can indicate bowel obstruction [19, 22]. |
| Number of times pelvic pain reported | 2 years | 4 | Pelvic Pain | Pelvic pain can indicate bowel obstruction [19, 22]. |
| Number of times stomach disorders reported | 2 years | 4 | Stomach Disorders | Stomach disorders can indicate bowel obstruction [19, 22]. |

In practise, all of these variables had low case counts for the >1 event categories, and were simplified to binary variables using method 3 (new-onset events).

***Method 5: Regular events***

Within this study, an event was defined to be regular if it occurred more than 4 times in a 12-month period, *at any point* in the primary care records of an individual before the index date. This method was used to identify regular *prescriptions* of a certain types of medicine - which are known to modify long-term risk of colorectal cancer (e.g. aspirin) - prior to the index date.

**Table 7: List of predictors defined as the regular events extracted from UKB primary care records**

| Variable | Shortname | Reference for association (or other justification of inclusion) |
|---|---|---|
| Regular use of aspirin | Aspirin use | Aspirin has a well-established preventative association with colorectal cancer [24], however, we note that the NHS does not recommend regular use of aspirin to prevent colorectal cancer [25], except in the case of individuals with Lynch syndrome [26].<br>In a systematic review of colorectal cancer models, eight (out of 52) included aspirin as a protective risk factor [27]. |
| Regular use of NSAIDs | NSAID use | NSAIDs have a well-established preventative association with colorectal cancer [28], however, given the side effects of long-term use are not recommended as a prevention measure by healthcare providers. In a systematic review of colorectal cancer models, 13 (out of 52) included aspirin as a risk factor [27]. |

$$\text{Regular event} = \begin{cases} 1, & \text{if there are} \geq 4 \text{ records of an event in any 12 month period before the index date} \\ 0, & \text{for everyone else} \end{cases}$$

***Method 6: Test results – direct and proxy measures***

When considering blood tests (or other biomarkers) we want to know if the test was carried out, and the recorded result of that test (or the "value"). A test being carried out may be a proxy for the presence of relevant symptoms or may indicate that the healthcare provider suspected underlying cancer, while an "abnormal result" is a potentially a clinical marker of disease. As these variables are markers (rather than modifers) of disease, we are only interested in events of this type close to diagnosis, and therefore again use a short lookback period (2 years).

$$\text{Test carried out} = \begin{cases} 1, & \text{if there is} \geq 1 \text{ record of the measure of interest in the lookback period} \\ 0, & \text{if there is no record of the relevant test in the lookback period} \end{cases}$$

$$\text{Abnormal result} = \begin{cases} 1, & \text{if there are} \geq 1 \text{ abnormal results for the measure of interest in the lookback period} \\ 0, & \text{if there are no abnormal results recorded in the lookback period} \end{cases}$$

However, blood tests – especially direct measures of a clinical marker – may be relatively sparse in the dataset. To maximise our use of the available data, we prioritise direct measurements of a clinical marker, but in the absence of a direct measure also consider suitable proxy measures. For example, iron deficiency anaemia (a symptom of colorectal cancer) is defined as low haemoglobin and low ferritin. However, the test for ferritin is not included in the standard blood test, so there are fewer results for this test than for haemoglobin. Therefore, in individuals without a test for both haemoglobin *and* ferritin, we also look for low levels of mean cell volume in the presence of low haemoglobin and low haemoglobin alone if no other relevant tests were carried out.

**Table 8: List of blood test variables extracted from UKB primary care records**

| **Variable** | **Lookback Period** | **Shortname** | **Reference for association (or other justification of inclusion)** |
|---|---|---|---|
| Test for inflammation carried out | 2 years | Inflammation (measured) | A record of a test for inflammation, which suggests that the primary care provider may suspect underlying cancer or another condition. |
| Inflammation | 2 years | Inflammation (abnormal) | C-reactive protein (CRP), erythrocyte sedimentation rate (ESR) and plasma viscosity (PV) are all inflammatory markers that can indicate the presence of cancer (amongst other causes) [29]. Inflammation may also be indicated indirectly by measurements of thrombocytosis (high levels of platelets) [30], or hyperferritinemia (abnormally high levels of ferritin) [31]. Additionally, hypoalbuminemia (abnormally low levels of serum albumin) may indicate colorectal cancer specific inflammation [32]. |
| Test of iron deficiency anaemia carried out | 2 years | Iron deficiency anaemia (measured) | A record of a test for iron deficiency, which suggests that the primary care provider may suspect underlying cancer or another condition. |
| Iron deficiency anaemia | 2 years | Iron deficiency anaemia (abnormal) | Iron deficiency anaemia is a well-established feature of colorectal cancer, and it often present at diagnosis (PPV is around 10%) [33].<br><br>Iron deficiency anaemia is defined as abnormally low levels of both haemoglobin and ferritin. Additionally, microcytosis (abnormally low mean cell volume (MCV)) accompanied by low haemoglobin is a strong indicator of iron deficiency and independently associated with colorectal cancer [34]. Iron deficiency anaemia may also be indicated by low levels of haemoglobin alone in the absence of a test of ferritin or MCV levels. |

For each blood test considered, we need to define the normal and abnormal ranges in order to categorise the results. We drew the thresholds used from NICE guidelines where available, and in other cases, thresholds used elsewhere in the literature.  The normal range of several variables is defined differently depending on other characteristics (e.g. sex and age) of the cohort member.

**Table 8: Consituent measures and their respective thresholds for the predictor iron deficiency anaemia**

| Measurement Type | Description | Thresholds |
|---|---|---|
| **Direct** | Low haemoglobin *AND* low ferritin | Haemoglobin: women < 120g/L, men <130g/L [35]<br>Ferritin < 30µg/L [35] |
| **Proxy 1** | Low haemoglobin *AND* low MCV | Haemoglobin: women < 120g/L, men <130g/L [35]<br>MCV < 85fL [34] |
| **Proxy 2** | Low haemoglobin (in the absence of ferritin and MCV measurements) | Haemoglobin: women < 120g/L, men <130g/L [35] |
| **Proxy 3** | Low MCV (in the absence of haemoglobin measurements) | MCV < 85fL [34] |

**Table 9: Consituent measures and their respective thresholds for the predictor inflammation**

| Measurement Type | Description | Thresholds |
|---|---|---|
| **Direct** | High CRP, ESR or PV | CRP > 6.8 mg/L [36-38]<br>PV > 1.72 mPa.s [36-38]<br>ESR (stratified by age and sex, see Table 9a) [36-38] |
| **Proxy 1** | High platelets (in absence of CRP/ESR/PV measurements) | Platelets > 450 $10^9$L [30] |
| **Proxy 2** | Low albumin (in absence of CRP/ESR/PV measurements) | Albumin < 35 g/L [32] |
| **Proxy 3** | High ferritin (in the absence of haemoglobin measurements) | Ferritin: men> 300 µ g/L, women > 200 µ g/L [31] |

**Table 9a: ESR thresholds**

| ESR thresholds (mm/hr) | Men | Women |
|---|---|---|
| **<40** | >11 | >14 |
| **40-49** | >12 | >15 |
| **50-59** | >14 | >17 |

| | | |
|---|---|---|
| **60-69** | >14 | >18 |
| **70-79** | >20 | >22 |
| **>80** | >20 | >23 |

***Implementation of a Multi-morbidity Score***

In previous studies, pre-existing conditions or co-morbidities have been shown to affect the diagnostic process and management of symptoms of as-yet-undiagnosed prevalent cancers - including colorectal cancer [23, 39-41]. However, adding many different conditions with small event numbers to the model as separate predictors is not desirable. Therefore, it was decided to use the Cambridge multi-morbidity score, previously developed in a UK primary care population, to summarise these characteristics for members of the cohort in this analysis.

The Cambridge multi-morbidity score combines 37 measures of morbidity to give an overall measure of health [42]. The score was developed for use with electronic health records, and makes use of several of the methods described in the previous sections of this document and uses a variety of lookback period lengths depending on the clinical properties of each morbidity. Codelists for the conditions are provided online by the research group that developed the score [43], these were then converted to the coding frameworks used in UK Biobank as described previously.

The original paper developed a range of scores for different outcomes (GP consultation rate, mortality, hospital admission, general outcome), each with a long version (containing all 37 conditions) and a short version (the 20 most “important” conditions). We used the long “general outcome”version of the Cambridge multimorbidity score, as this does not include variables for age and sex (these predictors are included separately in our model).

**Section C - Analysis Groupings**

In Table 10, the full set of candidate predictors that we plan to use in the colorectal cancer model development are given, grouped by both data type and analysis grouping. In the analysis, we looked at the impact of each group of variables (when added to the core group) separately and in combination.

The justification for the analysis groupings (or sets) is based on both data availability and information type:

1. Core

*Age, age group, birth year, sex, smoking status, body mass index (BMI) and Townsend deprivation index (TDI),  Ethnicity*

This set of predictors are all extracted from the UKB baseline assessment and use information easily available to GPs currently.

2. Polygenic Score

*Polygenic Score (PRS-CSx) and ten principal components**

Genetic data for UKB participants was collected at baseline. Although genetic information is not currently available in primary care setting, it is likely that genetic information will become routinely available in the near future [44].

3. Other Lifestyle

*Education (type), alcohol consumption, processed meat consumption, red meat consumption, fibre consumption,*

This set of predictors are all extracted from the UKB baseline assessment, and cover information we do not typically expect to be available in primary care records. If models of this type were implemented within routine primary care, collection of this type of data may present an addtional burden for both patients and the healthcare system.  Additionally, this type of predictor typically relies on self-reporting, hence, accurate measurement is challenging (e.g. dietary variables, alcohol consumption) [45, 46].

4. Symptoms

*Alarm symptoms: abdominal lump, rectal mass, change in bowel habit, rectal bleeding,*
*New onset symptoms: constipation, diarrhoea, fatigue, jaundice, weight loss*
*Incidence of Symptoms: Abdominal bloating, abdominal pain, pelvic pain, stomach disorders*
*Symptom proxy/misdiagnoses: recent diagnosis of diverticular disease, recent diagnosis of irritable bowel syndrome (IBS), recent diagnosis of haemorrhoids*

These variables are created using information primarily from primary care clinical records, with some information from the prescription records also being extracted.  All of the information needed to derive these variables is expected to be routinely collected and present in primary care records.

5. Medical History

*Family history of bowel cancer, recent colonoscopy, eligible for bowel cancer screening, inflammatory bowel disease (IBD), Type 2 Diabetes, Type 1 Diabetes, Gallstones (or cholelithiasis), regular use of aspirin, regular use of NSAIDs, Multimorbidity Score.*

These predictors are primarily extracted from primary care records (note family history data is taken from from UKB baseline) and include indicators of overall health and family history. The risk factors in this grouping are expected to be present over a long period (modifiers of risk) and this is reflected in longer lookback periods (6-10 years) or use of the whole clinical history avlaible from the primary care reocrds. Most of the information needed to determine these variables is routinely collected and present in GP records.

6. Common blood tests (and results)

*Iron deficiency anaemia and inflammation*

These variables are created using information from the clinical primary care records only. All of the information needed to derive these variables is expected to be routinely collected and present in primary care records. For each included indicator of colorectal cancer, we created two variables (one identified if the cohort member has had the test in the lookback period, the second if the results were abnormal).

**Table 10**: Summary of Variables by data type and analysis grouping (n=57)

| | | Baseline Assessment | | Electronic Health Records | |
|---|---|---|---|---|---|
| | | **Phenotypic** | **Genetic** | **Clinical Record (GP)** | **Prescription Records** |
| **Analysis Grouping** | **Core (n=8)** | Age: Age, Age Group, Birth Year<br>Sex<br>BMI<br>Smoking<br>Townsend Deprivation Index<br>Ethnicity | | | |
| | **Medical History (n=12)** | Family History of Bowel Cancer<br>Family History of Breast Cancer<br>Family History of Lung Cancer<br>Eligible for Bowel Cancer Screening | | Multimorbidity score*<br>Colonoscopy in last 10 years<br>Comorbidities: Type 1 diabetes, type 2 diabetes, inflammatory bowel disease, gallbladder calculus | Multimorbidity score*<br>Regular use: NSAIDs, aspirin |
| | **Symptoms (n=16)** | | | Ever: Abdominal lump, Rectal Mass, Change in bowel habit, Rectal bleeding<br>Rate: Abdominal bloating, Abdominal pain, Pelvic pain, Stomach disorders<br>New Onset: Constipation*, Diarrhoea*, Weight loss, Jaundice, Fatigue, diverticular disease, irritable bowel syndrome, haemorrhoids | New Onset: Constipation*, Diarrhoea*, |
| | **Biomarkers (n=4)** | | | Iron deficiency anaemia: test carried out, abnormal result<br>Inflammation: test carried out, abnormal result | |
| | **Genetic (n=11)** | | Polygenic Score (PRS-CSx)<br>Principal components (1-10) | | |
| | **Additional Lifestyle (n=6)** | Dietary Variables: processed meat, red meat, fibre score<br>Education: highest qualification<br>Alcohol consumption | | | |

*Variables that use both clinical and prescription EHR
